# Early Life Experiences and Adult Orientation to Promote Good in 22 Countries

**DOI:** 10.1101/2025.03.14.25323975

**Authors:** Ying Chen, Eric S. Kim, Julia S. Nakamura, Dorota Weziak-Bialowolska, Renae Wilkinson, R. Noah Padgett, Byron R. Johnson, Tyler J. VanderWeele

**Author notes:** **Corresponding author:** Dr. Ying Chen, Department of Epidemiology, Harvard T. H. Chan School of Public Health, Kresge Building, 677 Huntington Avenue, Boston, MA 02115.

## Abstract

Prior research suggests associations between character involving an orientation to promote good (i.e., a disposition to take actions that contribute to the good of oneself and others) and improved well-being outcomes. However, less is known about childhood factors that may lead to a greater disposition to promote good. This study used data from 202,898 adults in 22 countries to evaluate childhood antecedents of an orientation to promote good. We examined the associations between retrospectively reported childhood experiences and adult disposition to promote good in each country individually, and cross-nationally by meta-analytically pooling results across countries. The pooled results suggest that childhood experiences including having positive relationships with parents, higher subjective financial status, better childhood self-rated health, frequent religious service attendance, an earlier year of birth, and being female were associated with a greater orientation to promote good in adulthood. Conversely, the childhood experiences of abuse and feeling like an outsider in the family were associated with lower levels of promoting good. In country-specific analyses, the direction and strength of these associations differed by country, indicating diverse societal influences. This study provides a valuable foundation for future investigations into the influence of childhood experiences on character across cultures and national contexts.

## INTRODUCTION

In addition to reducing risk factors and illness, there has been growing interest in a strength-based approach that focuses on fostering capabilities to enhance well-being^1^. One inherent human strength that has been considered as essential for a flourishing life is expressing character, for example, living in accord with one’s ethical values^2,3^. For many people this will involve an orientation to promote what is good for oneself and for others.

In this context, researchers, particularly in the field of positive psychology, have become interested in understanding character strengths—positive attributes with moral significance that are crucial for a person’s identity, sense of self, and fulfillment^4^. Prior researchers have identified a total of 24 character strengths (e.g., kindness, perseverance, fairness, curiosity, gratitude, etc.), which reflect central ethical beliefs that are nearly ubiquitously valued across cultures^5–7^. Each of these character strengths can be assessed on a continuum with validated measures^5^. Beyond character strengths, other measures have been proposed to assess character that involves a global disposition to promote the good of both oneself and others^1^. An example item for measuring disposition to promote good is “I always act to promote good in all circumstances, even in difficult and challenging situations”^1,8^. This measure does not take a position on what “goodness” is, but rather evaluates the extent to which people report acting in line with their own perceptions of what is good. The understanding of “goodness” may vary across individuals, cultures, and societies, and this measure provides insights into how people perceive their capacity to act for good in general. People with a greater disposition to promote good may be more likely to express specific character strengths across contexts, thus disposition to promote good may be understood as a higher-order measure of character and an upstream factor that influences behaviors to express character (e.g., service to others, volunteering), what one might refer to as “moral perseverance”^9^. Prior empirical studies suggest that disposition to promote good is generally positively associated with health and well-being outcomes, such as greater life satisfaction^10^, happiness^11^, mental health^12^, self-rated physical health^8^, social connectedness^8^, and flourishing^13^. There is also evidence suggesting that character may be modified via strength-based interventions that aim to help people fulfill their potential^14–16^.

While living in accord with one’s ethical values is a potential inherent to all humans^17,18^, socioeconomic and cultural factors may shape the expression of character^19^. For example, contextual factors such as greater economic development, reduced income inequality, and higher levels of institutional trust are all correlated with higher rates of prosocial behavior in a society^6,20,21^. As another example, individualistic societies emphasize autonomy and personal goals, whereas collectivist cultures more greatly value interpersonal harmony and group goals^22^. Therefore, the understanding of perseverance in individualistic countries may focus on personal achievement, whereas in collectivist cultures perseverance may also demonstrate one’s commitment to the group’s harmony and success^23^. Cultural values may shape an individual’s character starting from early life by influencing the environment in which they grow and live. Some countries have incorporated character education into school curriculum to cultivate culturally-valued strengths in children and adolescents. For instance, some Asian countries have included teachings about filial piety and humility in their character education programs^24,25^.

The Psychosocial Development Theory indicates that character formation is a process that unfolds over the life course, but early life may be a critical period for fostering character^26,27^. Prior empirical studies found that population patterning of prosociality begins to diverge around middle childhood across societies, adding to the evidence that childhood may be a period when individuals are particularly sensitive to cultural influences^26^. Other studies also found that the link between character strengths and well-being is established rather early in life, implying that strength-based interventions may be most effective when introduced at a young age^28^. Although childhood predictors of orientation to promote good have seldom been studied specifically, prior research has suggested that multiple dimensions of childhood experiences may shape one’s character broadly. First, *family environment* may play an important role in character formation^29^. For instance, parenting practices such as parental warmth, role modelling, communication styles, and teaching about life may all contribute to the child’s development of ethical reasoning, emotional intelligence, responsibility, and empathy^30^. According to the Attachment Theory^31^, secure attachment with caregivers is crucial for developing social competence and emotional regulation, which in turn fosters the growth of trust and empathy. Character strengths of parents and their children also tend to converge, with both maternal and paternal influences shaping the child’s character^32^. This may be understood with the Social Learning Theory^33^, which posits that children tend to model their parents’ behaviors and incorporate those traits into their character. Some other aspects of family environment that are directly or vicariously related to parenting, such as parents’ marital status, have also been linked with the child’s character^34^. Second, experiences of *childhood adversities* may also impact character. For instance, a meta-analysis found that adults who experienced early-life adversities (e.g., childhood financial difficulties, childhood abuse) on average showed lower tendencies to share resources, possibly because they have had limited opportunities to develop capacities for expressing character^35^. To our knowledge, whether adverse childhood experiences influence one’s general orientation to promote good, or how different types of childhood adversities may differentially affect character formation, have seldom been studied specifically. It is possible that childhood financial difficulties play a greater role in prosocial acts that involve sharing material resources, whereas other types of childhood adversities (e.g., child abuse) may affect non-material acts of kindness. Third, many *faith traditions* worldwide emphasize character as an important avenue for fostering meaning and purpose in life^36^. Although the association between religion and one’s general disposition to promote good has seldom been studied directly, empirical evidence based on data from the United States suggests that a religious upbringing is positively associated with engagement in prosocial activities (e.g., volunteering, civic engagement) and the levels of specific character strengths (e.g., forgiveness) in adulthood^37^.

Past studies have substantially advanced our understanding of character, but knowledge gaps remain. First, most prior research studied specific character strengths or prosocial behaviors, but few studies have examined one’s general orientation to promote good or its antecedents. Examining character involving the promotion of good is essential because it provides insights into how a person’s overarching ethical orientation might consistently drive impactful behaviors across various aspects of life (e.g., perseverance in achieving goals, contributing to community initiatives, promoting fairness in decision-making). The measurement tools for assessing disposition to promote good are easy to implement, which have great potential to be used in large-scale surveys^1,28^. Understanding approaches to cultivate character involving an orientation to promote good will also inform strengths-based programs that may empower individuals and communities to fulfill their potential for growth and fulfillment, which may also promote social inclusion and reduce disparities in population well-being^14–16^. Second, many prior studies on character have been conducted among participants from Western, Educated, Industrialized, Rich, and Democratic (WEIRD) countries and/or used non-representative samples^38^. A more comprehensive understanding of character across diverse cultures and national contexts is needed for developing culturally appropriate strength-based programs that help individuals cultivate character and realize their potential.

To begin addressing these knowledge gaps, using data from the Global Flourishing Study (which enrolled 202,898 adults from 22 diverse countries, with the sample weighted to be nationally representative within each country), this study examined a range of childhood candidate antecedents of disposition to promote good in adulthood. We hypothesized that these childhood experiences, personal attributes, and familial or social circumstances would have varied associations with adult orientation to promote good. We also expected that the direction and strength of these associations would differ by country, reflecting diverse societal influences.

## METHODS

This study used Wave 1 data from the Global Flourishing Study (GFS). The description of the methods below has been adapted from VanderWeele et al.^39^ (the paper by VanderWeele et al. is scheduled to be published before the present study, and the reference will be updated when it becomes available). Further methodological detail regarding the GFS is available elsewhere^40–45^. The present study is a secondary analysis of the existing GFS data.

### Study Population

The Global Flourishing Study (GFS) is a longitudinal study that enrolled 202,898 adults (age range: 18 to 99 years) from 22 culturally and geographically diverse countries, with the samples weighted to be nationally representative within each country. Survey items queried aspects of well-being including happiness, health status, sense of meaning and purpose, character, social relationships, and financial stability, along with a range of demographic, socioeconomic, political, religious, personality, childhood, community, and behavior factors. Data collection was carried out by Gallup via a combination of modes (e.g., in-person, phone, web) that varied across countries^45^. Data for Wave 1 were collected principally during 2023, while some countries began data collection in 2022^40^. The following countries were included in Wave 1 data collection: Argentina, Australia, Brazil, Egypt, Germany, Hong Kong (Special Administrative Region of China), India, Indonesia, Israel, Japan, Kenya, Mexico, Nigeria, the Philippines, Poland, South Africa, Spain, Sweden, Tanzania, Turkey, United Kingdom, and the United States. Four additional waves of panel data on the participants will be collected annually from 2024-2027.

The precise sampling design to ensure nationally representative samples varied by country^40^. During the survey translation process, Gallup adhered to TRAPD model (translation, review, adjudication, pretesting, and documentation) for cross-cultural survey research (ccsg.isr.umich.edu/chapters/translation/overview)^44^. The data that supports this research are publicly available via the Center for Open Science (COS, https://www.cos.io/gfs). Details about the GFS study methodology and survey development are reported elsewhere^40^ (see also https://osf.io/y3t6m).

The present study used data from all participants in Wave 1 of GFS (N=202,898). Poststratification and nonresponse adjustments were performed to ensure the sample was representative of the adult population in each country^40,45^. Ethical approval was granted by the institutional review boards at Baylor University and Gallup, and all participants provided informed consent. All methods were carried out in accordance with relevant ethical guidelines and regulations.

### Assessment of Orientation to Promote Good

An item from VanderWeele’s Flourishing Index^1^ was used to assess one’s general orientation to promote good: “I always act to promote good in all circumstances, even in difficult and challenging situations”. The response was rated on a Likert scale ranging from 0 (not true of me at all) to 10 (completely true of me). The response was analyzed as a continuous score, with a higher score indicating a greater orientation to promote good.

### Assessment of Childhood Factors

We investigated a range of individual-, interpersonal-, and familial-level childhood experiences, as described below. We also examined several childhood demographic factors (e.g., year of birth, gender, race/ethnicity), which are important demographic correlates of health.

#### Relationship with parents

Relationship with one’s parents in childhood was assessed with the question: “Please think about your relationship with your mother/father when you were growing up. In general, would you say that relationship was very good, somewhat good, somewhat bad, or very bad?”. Relationships with mother and father were queried separately. To reduce collinearity, in regression models the variables of relationship with mother and father were both dichotomized as “very/somewhat good” and “very /somewhat bad”. Responding “Does not apply” to either parental relationship indicator was treated as a dichotomous control variable for respondents who did not have a mother or father due to death or absence.

#### Parents’ marital status

Parents’ marital status was assessed with the question: “Were your parents married to each other when you were around 12 years old?”. Response options included “married”, “divorced”, “never married”, and “one or both of them had died”.

#### Subjective financial status

Participants reported the financial status of their family as it was when they were growing up, in response to the question: “which of these phrases comes closest to your own feelings about your family’s household income when you were growing up, such as when you were around 12 years old?”. Response options included “lived comfortably”, “got by”, “found it difficult”, and “found it very difficult”.

#### Childhood abuse

Childhood abuse was assessed with the question: “Were you ever physically or sexually abused when you were growing up?”. Responses included “yes” and “no”. This information was not obtained in Israel due to restrictions on asking such questions.

#### Felt like an outsider in family

Participants’ sense of family belonging during childhood was assessed with the question: “When you were growing up, did you feel like an outsider in your family”. Response options included “yes” and “no”.

#### Self-rated health

Participants reported their health status during childhood in response to the question: “In general, how was your health when you were growing up? Was it excellent, very good, good, fair, or poor?”

#### Immigration status

Immigration status was assessed with the question “Were you born in this country?” The responses include “yes” and “no”.

#### Religious service attendance

Participants’ frequency of religious service attendance during childhood was assessed with a question: “How often did you attend religious services or worship at a temple, mosque, shrine, church or other religious building when you were around 12 years old”. The response options range from 1 (at least once a week) to 4 (never).

#### Birth Year

Participants reported their current age (in years), and their birth years were calculated as the year of data collection minus their current age.

#### Gender

Participants reported their gender, and the response options include “male”, “female”, and “other gender identities”.

#### Religious affiliation

Religious affiliation in childhood was measured with the question: “What was your religion when you were 12 years old”. Response options include 15 major religions (e.g., Christianity, Islam, Hinduism, Buddhism, Judaism etc.), “some other religion”, and “No religion/Atheist/Agnostic”. The response categories varied as appropriate for each country, and this variable was included in the country-specific analyses only but not in the meta-analysis across countries^46^. To reduce data sparsity, in regression models the response categories of religious affiliation with a prevalence <3% were collapsed. “No religion/Atheist/Agnostic” was used as the reference group when at least 3% of the observed sample within the country endorsed this category; otherwise, the most prominent religious group was used as the reference category.

#### Race and ethnicity

Race and ethnicity were assessed in most but not all countries (Germany, Japan, Spain, and Sweden had restriction on collecting such data). Response options varied as appropriate for each country, so this variable was included in the country-specific analyses only but not in the meta-analysis across countries. To reduce data sparsity, in regression models race and ethnicity was dichotomized as racial and ethnic plurality (the category with the largest proportion) and minority (collapsing other categories) in each country.

### Statistical Analyses

Descriptive analyses examined the distribution of childhood factors in individual countries and in the total combined sample. The results were weighted to be nationally representative within each country.

In the country-specific analyses, a weighted linear regression model with complex survey adjusted standard errors was used to regress orientation to promote good on *all childhood factors simultaneously* to evaluate their associations with promoting good independently from each other. A Wald-type test was conducted to obtain a global (joint) test of the effect of all categories within a childhood factor, resulting in a single global p-value for each childhood factor.

Random effects meta-analysis^47,48^ was conducted to pool regression coefficients from the country-specific analyses, along with confidence intervals, lower and upper limits 95% prediction intervals, heterogeneity (τ), and I^2^ for estimating variation within a given childhood factor category across countries^49^. Forest plots of estimates are presented in the online supplement. Religious affiliation and race and ethnicity (when available) were used as control variables within country (their coefficients are reported in the online supplement), but these coefficients themselves were not included in the meta-analyses because their response options varied by country. A pooled global p-value^50^ was reported across countries to evaluate if the association between each childhood factor and adult orientation to promote good holds within at least one country. Bonferroni corrected p-value thresholds are provided based on the number of childhood factors that we examined^51,52^.

Meta-analyses were conducted in R^53^ using the metafor package^54^, and country-specific analyses were conducted using SAS 9.4 (SAS Institute, Inc). All statistical tests were 2-sided. For each childhood predictor, in both the meta-analyses and country-specific analyses we calculated E-values to evaluate sensitivity of the observed associations to potential unmeasured confounding. An E-value is the minimum strength of the association an unmeasured confounder must have with both the outcome and the predictor, above and beyond all measured covariates, for an unmeasured confounder to explain away the observed predictor-outcome association^55^. As a supplementary analysis, we performed a population weighted meta-analysis to pool the country-specific estimates. All analyses were pre-registered with COS prior to data access, with only slight subsequent modification in covariate adjustments due to multicollinearity (https://doi.org/10.17605/OSF.IO/Y4U6W); all code to reproduce analyses are openly available in an online repository^42^.

### Missing Data

In the total combined sample, 0.33% of participants had missing data on orientation to promote good, and missing data on childhood factors ranged from 0.01% to 9.78% (except race/ethnicity, which was not measured in some countries). We conducted multivariate imputation by chained equations (with 5 imputed datasets created) to impute missing data on all variables in each country separately^56–58^. We included sampling weights in the imputation models to account for specific-variable missingness that may have been related to probability of inclusion in the study^45^.

### Accounting for Complex Sampling Design

The GFS used different sampling schemes across countries based on availability of existing panels and recruitment needs^40^. All analyses accounted for the complex survey design components by including weights, primary sampling units, and strata. Additional methodological details were reported elsewhere^40^.

## RESULTS

### Descriptive Analyses

In the total sample combined across countries (Table 1), a majority of the participants reported on their childhood experiences as follows: had very/somewhat good relationships with mother (89%) and father (80%), had married parents (75%), lived comfortably financially or got by in childhood (76%), did not experience child abuse (82%), did not feel like an outsider in the family (84%), had excellent or very good health (64%), and were native-born (94%). Additionally, a large proportion of participants attended religious services at least once per week during childhood (41%). Participants’ current age ranged from 18 to 99 years, and there was a largely similar proportion of participants from different age groups (except that fewer participants are from earlier birth cohorts with current age above 70 years). The sample has a balanced ratio of female (51%) and male (49%) participants, and a small proportion (0.3%) who identified as other gender. The sample size for each country ranged from 1,473 (Turkey) to 38,312 individuals (United States). See Supplementary Table S1A to S22A for participant characteristics in each individual country.

**Table 1.** Distribution of Childhood Factors in the Overall Sample Combined Across 22 Countries (N=202,898, With the Sample Weighted to Be Nationally Representative within Each Country)

| **Childhood factors** | **Category** | **N (%)** |
| --- | --- | --- |
| Relationship with mother | Very good | 127,836 (63%) |
|  | Somewhat good | 52,439 (26%) |
|  | Somewhat bad | 11,060 (5%) |
|  | Very bad | 4,642 (2%) |
|  | Does not apply | 5,965 (3%) |
|  | Missing | 956 (0%) |
| Relationships with father | Very good | 107,742 (53%) |
|  | Somewhat good | 55,714 (27%) |
|  | Somewhat bad | 15,807 (8%) |
|  | Very bad | 8,278 (4%) |
|  | Does not apply | 13,985 (7%) |
|  | Missing | 1,372 (1%) |
| Parent marital status | Parents married | 152,001 (75%) |
|  | Divorced | 17,726 (9%) |
|  | Single, never married | 15,534 (8%) |
|  | One or both parents had died | 7,794 (4%) |
|  | Missing | 9,843 (5%) |
| Financial status of family when growing up | Lived comfortably | 70,861 (35%) |
|  | Got by | 82,905 (41%) |
|  | Found it difficult | 35,852 (18%) |
|  | Found it very difficult | 12,606 (6%) |
|  | Missing | 674 (0%) |
| Experienced abuse | Yes | 29,139 (14%) |
|  | No | 167,279 (82%) |
|  | Missing | 6,479 (3%) |
| Felt like an outsider in the family | Yes | 28,732 (14%) |
|  | No | 170,577 (84%) |
|  | Missing | 3,589 (2%) |
| Self-rated health when growing up | Excellent | 67,121 (33%) |
|  | Very good | 63,086 (31%) |
|  | Good | 47,378 (23%) |
|  | Fair | 19,877 (10%) |
|  | Poor | 4,906 (2%) |
|  | Missing | 530 (0%) |
| Immigration status | Born in this country | 190,998 (94%) |
|  | Born in another country | 9,791 (5%) |
|  | Missing | 2,110 (1%) |
| Frequency of religious service attendance (age 12) | At least 1/week | 83,237 (41%) |
|  | 1-3/month | 33,308 (16%) |
|  | <1/ month | 36,928 (18%) |
|  | Never | 47,445 (23%) |
|  | Missing | 1,980 (1%) |

| Childhood factors | Category | N (%) |
| --- | --- | --- |
| Year of birth | 1998-2005; current age: 18-24 | 27,007 (13%) |
|  | 1993-1998; age 25-29 | 20,700 (10%) |
|  | 1983-1993; age 30-39 | 40,256 (20%) |
|  | 1973-1983; age 40-49 | 34,464 (17%) |
|  | 1963-1973; age 50-59 | 31,793 (16%) |
|  | 1953-1963; age 60-69 | 27,763 (14%) |
|  | 1943-1953; age 70-79 | 16,776 (8%) |
|  | 1943 or earlier; age 80+ | 4,119 (2%) |
|  | Missing | 20 (0%) |
| Gender | Male | 98,411 (49%) |
|  | Female | 103,488 (51%) |
|  | Other | 602 (0%) |
|  | Missing | 397 (0%) |
| Country | Argentina | 6,724 (3%) |
|  | Australia | 3,844 (2%) |
|  | Brazil | 13,204 (7%) |
|  | Egypt | 4,729 (2%) |
|  | Germany | 9,506 (5%) |
|  | Hong Kong (S.A.R of China) | 3,012 (1%) |
|  | India | 12,765 (6%) |
|  | Indonesia | 6,992 (3%) |
|  | Israel | 3,669 (2%) |
|  | Japan | 20,543 (10%) |
|  | Kenya | 11,389 (6%) |
|  | Mexico | 5,776 (3%) |
|  | Nigeria | 6,827 (3%) |
|  | Philippines | 5,292 (3%) |
|  | Poland | 10,389 (5%) |
|  | South Africa | 2,651 (1%) |
|  | Spain | 6,290 (3%) |
|  | Sweden | 15,068 (7%) |
|  | Tanzania | 9,075 (4%) |
|  | Türkiye | 1,473 (1%) |
|  | United Kingdom | 5,368 (3%) |
|  | United States | 38,312 (19%) |

Note. S.A.R. = Special Administrative Region.

### Childhood Factors Associated with Promoting Good in Adulthood: Pooled Estimates Across Countries

The random effects meta-analysis, which pooled country-specific estimates, suggested that, on average across countries, most of the childhood factors that we examined were associated with orientation to promote good in adulthood (Table 2). The global p-values for most childhood factors (except for relationship with mother and feeling like an outsider in the family) met the Bonferroni corrected threshold for statistical significance (*p*<0.0045), indicating that each of these childhood factors was associated with promoting good in at least one of the 22 countries. In the overall sample, having very/somewhat good (vs. very/somewhat bad) relationships with one’s mother (β=0.13, 95% confidence interval [CI]=0.04, 0.21) and with one’s father (β=0.09, 95% CI=0.03, 0.15) were each associated with higher scores of promoting good on average across countries. However, the global *p*-value for relationship with one’s mother passed *p*<.05 only before but not after the Bonferroni correction (*p*=0.01). On average across countries, living comfortably financially (vs. getting by) during childhood was associated with greater promoting good (β=0.13, 95% CI=0.06, 0.21). In addition, the childhood experiences of having excellent/very good vs. good self-rated health (e.g., β _excellent vs. good_=0.44, 95% CI: 0.27, 0.61) and frequent religious service attendance (e.g., β_at least once/week vs. never_=0.30, 95% CI: 0.21, 0.39) were each associated with a greater adult disposition to promote good. Conversely, the experiences of childhood abuse (β=-0.10, 95% CI: −0.17, −0.03) and feeling like an outsider in family when growing up (β=-0.11, 95% CI: −0.15, −0.06) were each inversely associated with promoting good. There was also evidence suggesting that earlier (vs. the most recent) birth cohorts reported a greater orientation to promote good (e.g., β _birth cohort 1943-1953 vs. 1998-2005_=0.31, 95% CI: 0.07, 0.56). Female participants on average reported higher levels of promoting good than male participants (β=0.07, 95% CI=0.03, 0.11). Although the confidence intervals for parent marital status and immigration status included the null value, the global *p* values were<.001, indicating evidence that these two childhood factors were associated with promoting good in at least one country but not necessarily on average across countries. The heterogeneity estimate τ evaluates the extent to which the estimates vary across countries. The calculated τ suggested that the variation across countries was particularly evident for certain categories of the childhood factors of self-rated health, year of birth, and gender, with the τ estimates above 0.40. The I^2^ estimates for certain categories of parent marital status, subjective financial status in childhood, self-rated health when growing up, immigration status, year of birth, and gender were relatively large (above 80), suggesting that variability in the estimates across countries for these childhood factor categories are mainly due to heterogeneity rather than sampling variability.

**Table 2.**
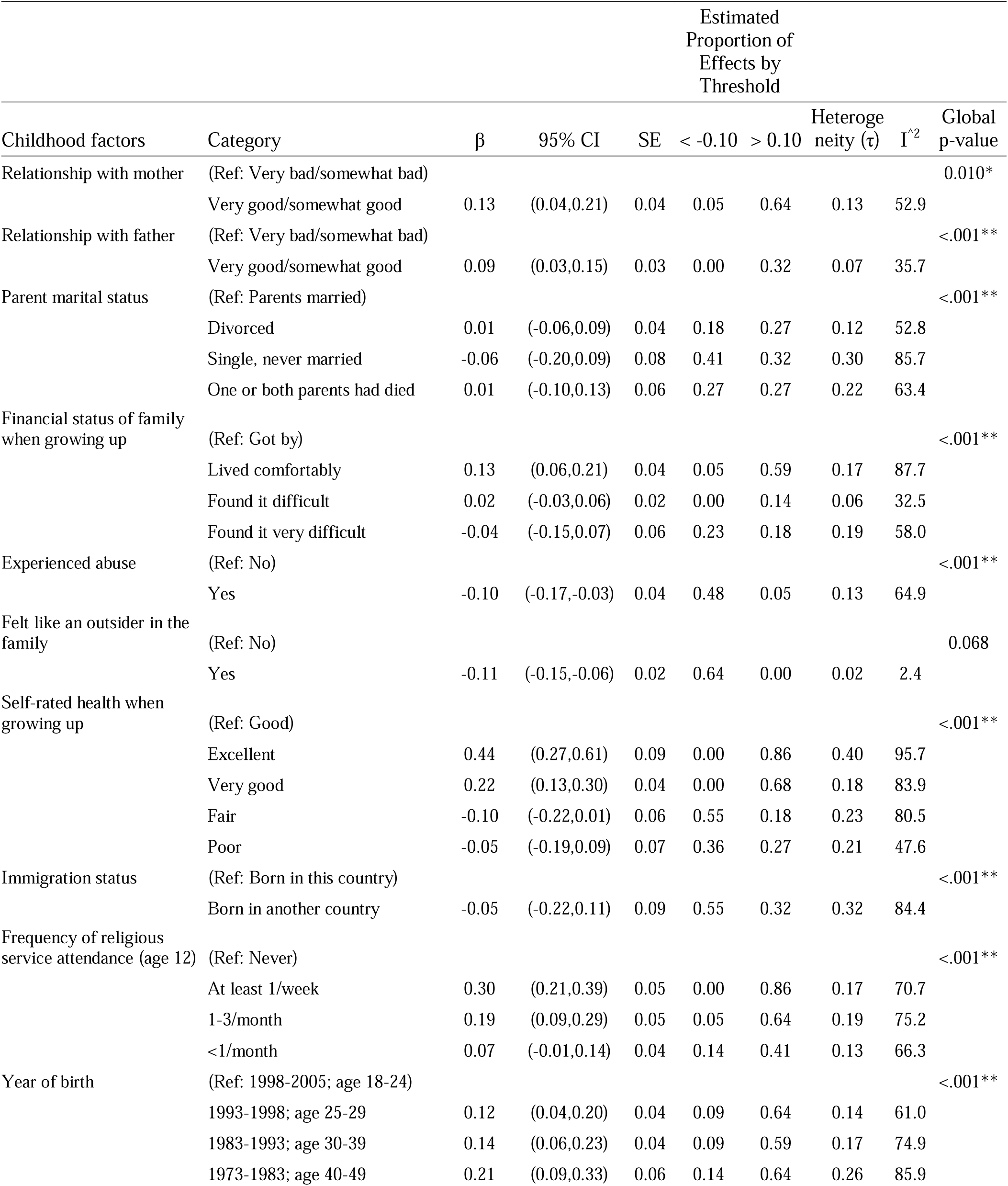
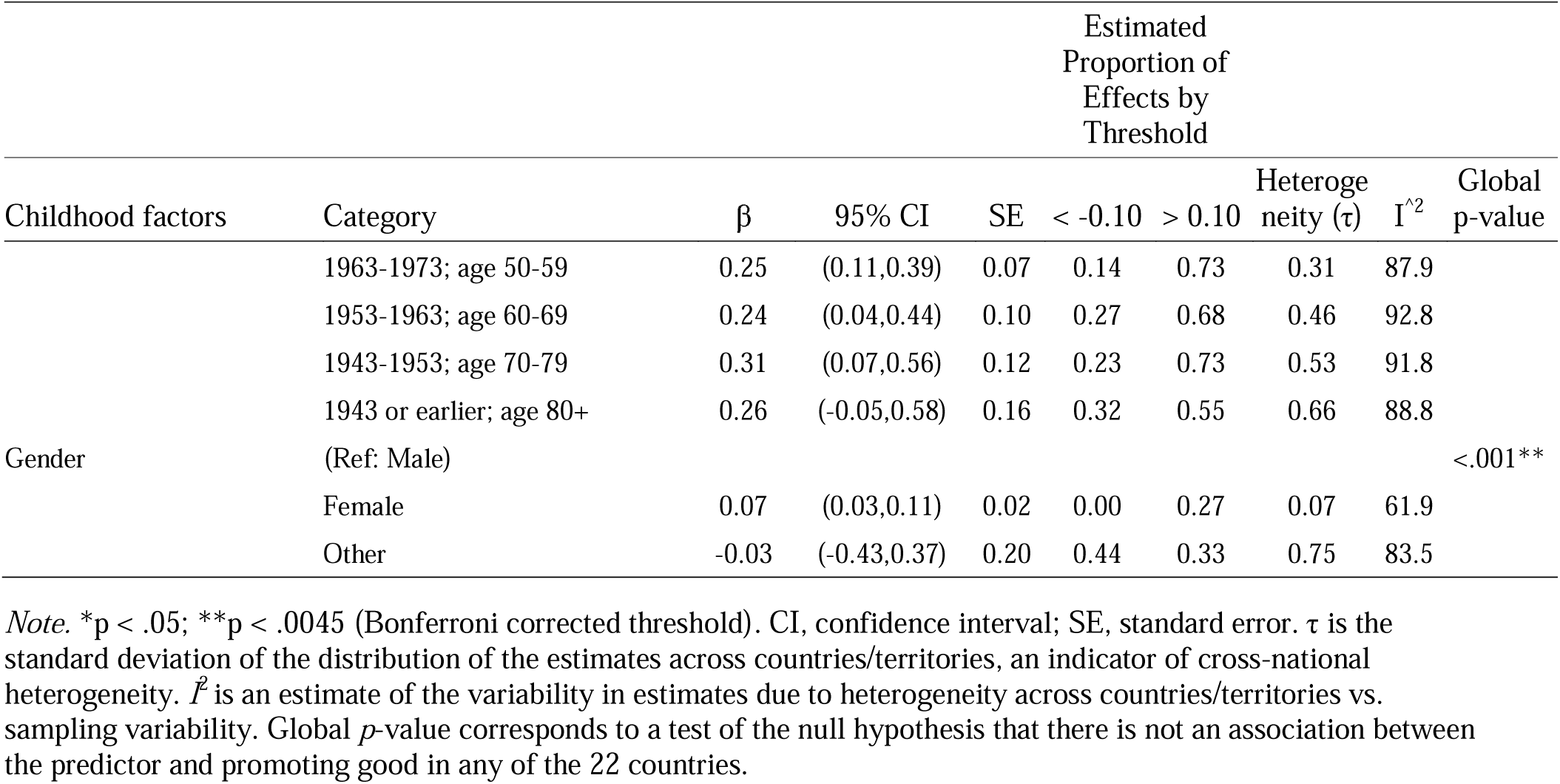
Random Effects Meta-Analysis of Association between Childhood Factors and Orientation to Promote Good in Adulthood (N=202,898).

The calculated “E-values” suggest that some of the observed associations between childhood experiences and promoting good in adulthood are moderately robust to potential unmeasured confounding (Table 3). For example, to explain away the association between excellent (vs. good) childhood self-rated health and adult levels of promoting good (β=0.44), an unmeasured confounder associated with both increased likelihood of excellent childhood health and higher levels of promoting good by risk ratios of 1.75 each, above and beyond all measured covariates, could suffice but weaker joint confounder associations could not. Likewise, to shift the confidence interval to include the null value, an unmeasured confounder associated with both increased likelihood of excellent health and higher levels of promoting good by risk ratios of 1.52-fold each could suffice, but weaker joint confounder association could not. The E-values for some other childhood factors such as weekly childhood religious service attendance are likewise of considerable magnitude.

**Table 3.** Sensitivity of Meta-Analyzed Associations between Childhood Factors and Orientation to Promote Good in Adulthood to Potential Unmeasured Confounding.

| Variable | Predictor (level) | E-value for Estimate | E-value for 95% CI |
| --- | --- | --- | --- |
| Relationship with mother | (Ref: Very bad/somewhat bad) |  |  |
|  | Very good/somewhat good | 1.31 | 1.16 |
| Relationship with father | (Ref: Very bad/somewhat bad) |  |  |
|  | Very good/somewhat good | 1.25 | 1.14 |
| Parent marital status | (Ref: Parents married) |  |  |
|  | Divorced | 1.07 | 1.00 |
|  | Single, never married | 1.19 | 1.00 |
|  | One or both parents had died | 1.09 | 1.00 |
| Financial status of family when growing up | (Ref: Got by) |  |  |
|  | Lived comfortably | 1.32 | 1.19 |
|  | Found it difficult | 1.10 | 1.00 |
|  | Found it very difficult | 1.16 | 1.00 |
| Experienced Abuse | (Ref: No) |  |  |
|  | Yes | 1.27 | 1.13 |
| Felt like an outsider in the family | (Ref: No) |  |  |
|  | Yes | 1.28 | 1.20 |
| Self-rated health when growing up | (Ref: Good) |  |  |
|  | Excellent | 1.75 | 1.52 |
|  | Very good | 1.45 | 1.32 |
|  | Fair | 1.28 | 1.00 |
|  | Poor | 1.18 | 1.00 |
| Immigration status | (Ref: Born in this country) |  |  |
|  | Born in another country | 1.19 | 1.00 |
| Frequency of religious service attendance (age 12) | (Ref: Never) |  |  |
|  | At least 1/week | 1.57 | 1.44 |
|  | 1-3/month | 1.40 | 1.25 |
|  | <1/month | 1.21 | 1.00 |
| Year of birth | (Ref: 1998-2005; age 18-24) |  |  |
|  | 1993-1998; age 25-29 | 1.30 | 1.15 |
|  | 1983-1993; age 30-39 | 1.34 | 1.19 |
|  | 1973-1983; age 40-49 | 1.43 | 1.25 |
|  | 1963-1973; age 50-59 | 1.49 | 1.28 |
|  | 1953-1963; age 60-69 | 1.48 | 1.15 |
|  | 1943-1953; age 70-79 | 1.58 | 1.22 |
|  | 1943 or earlier; age 80+ | 1.51 | 1.00 |
| Gender | (Ref: Male) |  |  |
|  | Female | 1.22 | 1.14 |
|  | Other | 1.13 | 1.00 |

Notes: The *E*-value for the effect estimate is the minimum strength of association (on the risk ratio scale) that an unmeasured confounder would need to have with both the predictor and the outcome to entirely explain away the observed association between them, conditional on the measured covariates. The *E*-value for the limit of the 95% confidence interval closest to the null denotes the minimum strength of association (on the risk ratio scale) that an unmeasured confounder would need to have with both the predictor and the outcome to shift the confidence interval to include the null value, conditional on the measured covariates.

### Childhood Factors Associated with Promoting Good in Adulthood: Country-Specific Estimates

The country-specific analyses (Supplementary Tables S1B to S22B; Supplementary Figures S1 to S27) found that in most countries the childhood experiences of family financial security, optimal self-rated health, and frequent religious service attendance were each associated with a greater disposition to promote good in adulthood (global *p*-values <.05). Specifically, living comfortably (vs. getting by) financially when growing up was associated with higher scores of promoting good across several culturally diverse but predominantly high-income or upper middle-income countries/territories (e.g., Hong Kong, Turkey, Japan, Philippines, Argentina, Mexico, United States, Spain, Sweden). Interestingly, in some of these same countries as mentioned above (including Argentina, Mexico, Spain, United States), lower levels of subjective financial status in childhood (i.e., found it very difficult/found it difficult vs. got by) were also positively associated with adult promoting good, representing a curvilinear relationship between childhood financial status and adult promoting good in these countries. Similarly, having excellent vs. good self-rated health in childhood was associated with considerably greater adult disposition to promote good in most countries, with the largest effect sizes in predominantly high-income countries/territories (e.g., Hong Kong, Japan, Sweden, United States). In addition, attending religious services at least once/week vs. never in childhood was associated with greater adult promoting good in a wide array of countries, even including some of the most secular countries in the world (e.g., Japan, Germany, Spain, Sweden)^59^.

The participants’ disposition to promote good was also strongly patterned by their birth year/current age in 19 out of the 22 countries that we examined, although the specific patterning varied widely across countries. For instance, participants who were older/from an earlier birth cohort generally reported a greater disposition of promoting good in most countries, and the evidence of such patterning was strongest in several high-income countries/territories (e.g., Hong Kong, Australia, United States, Sweden, Germany, Japan, Spain). In comparison, in some lower-middle countries (e.g., India, Tanzania), the association of birth cohorts/current age with promoting good was reversed in direction, whereby younger vs. older people reported a greater orientation to promote good.

There was also evidence suggesting that childhood experiences of parental marital status, immigration status, and gender were associated with promoting good in several countries, but the patterning varied across countries. For instance, being an immigrant vs. native-born was positively associated with promoting good in high-income countries (e.g., Spain, United States, United Kingdom, Germany, Sweden), whereas the direction of the association was reversed in several lower-middle income countries (e.g., Nigeria, India, Mexico, Brazil). While the meta-analyzed pooled estimates suggested that the childhood factors of having good relationship with one’s parents, child abuse, and feeling like an outsider in the family were each associated with promoting good on average across countries, such associations were present in only a handful of individual countries in the country-specific analyses (see Figure S1, S2, S9, and S10). For example, having very good/somewhat good (vs. very bad/somewhat bad) relationship with father was associated with greater adult promoting good in Japan (β=0.27, 95% CI: 0.19, 0.35) and Brazil (β=0.16, 95% CI: 0.04, 0.29), but the associations were null in all other individual countries. This might be due to the small number of participants who reported “very/somewhat bad relationships” with their parents in individual countries (in over half of the countries, only <5% and <10% of the participants reported having very/somewhat bad relationship with mother and with father). Likewise, the proportion of participants reporting experiences of child abuse or feeling like an outsider in the family was only around 10% in many cases, which may have led to limited statistical power.

The calculated E-values that evaluate sensitivity of the results from country-specific analyses to potential unmeasured confounding are reported in Supplementary Tables S1C to S22C.

The population weighted meta-analysis that pooled estimates across countries considering population sizes in each country yielded largely similar results as the random effects meta-analysis, except that the associations for relationships with one’s parents became weaker (Supplementary Tables S23 and S24). This may be attributed to the large weighting given to India in the population weighted meta-analysis. In our sample from India, only 2% of the participants reported having very/somewhat bad relationships (the reference group) with their parents, which may have resulted in limited statistical power.

## DISCUSSION

Character is considered to be an important domain that constitutes a flourishing life^1^. However, the antecedents of character, especially the childhood experiences that shape character in later life, remain understudied^60^. Using data from 22 countries with the samples weighted to be nationally representative in each country, this study expands the literature by considering a wide range of individual-, interpersonal-, and familial-level childhood factors as candidate antecedents of character that involves a disposition to promote the good of oneself and others.

This study provides novel evidence suggesting that having greater material resources (e.g., financial stability) and favorable conditions in non-material resources (e.g., optimal health status) during childhood may contribute to a greater disposition to promote good, particularly in several high-income and upper middle-income countries/territories. In the pooled estimates, the childhood experiences of living comfortably financially (vs. getting by) and having excellent (vs. good) health were each associated with a substantially higher disposition to promote good in adulthood on average across countries. The Reserve Capacity Framework^61^ suggests that individuals with greater material resources may encounter fewer stressors and have more opportunities to develop reserve capacity (e.g., sense of control, self-esteem, social support) for coping with stress, leading to greater health and well-being. We posit that this theory may be extended to understand childhood conditions and character formation: in some cases, those with more material resources in childhood may encounter fewer stressful life events when growing up (e.g., less likely to encounter food insecurity or lack of safe spaces for learning how to socially interact), and have more opportunities to develop capacities that allow them to express character and, for example, give to others (e.g., time and resources for engaging in prosocial activities).

Beyond material resources, favorable conditions in other dimensions such as optimal health status may likewise provide the physical and mental stamina that allow individuals to fully engage in activities that foster personal growth and meaningful social interactions. Interestingly, the positive associations of optimal childhood financial status (lived comfortably vs. got by) and childhood health (excellent vs. good) with promoting good are both particularly strong in high-income and upper-middle income countries/territories. We hypothesize that the overall wealth and stability in these nations may amplify the positive impact of individuals’ childhood resources on character expression. In these higher-income countries, where abundant resources are readily available—such as top-tier education (e.g., educational programs that focus on social-emotional learning), cultural programs (e.g., community arts centers, religious youth groups, museums that offer educational outreach), and community-focused initiatives (e.g., Habitat for Humanity youth programs, local food banks)—children with advantageous financial conditions and optimal health are particularly well-positioned to engage deeply in these enrichment or prosocial activities. This in turn can help them nurture: empathy, moral reasoning, community engagement, cultural awareness, and a strong sense of responsibility toward oneself and others—all factors that can contribute to an orientation of promoting good. Notably, it is also worth mentioning that in some of these same countries (e.g., Argentina, Spain, United States), childhood financial difficulties (found it difficult/very difficult financially vs. got by) were also associated with greater orientation to promote good in adulthood, indicating a curvilinear relationship between childhood financial status and adult promoting good in these countries. This finding is aligned with some prior evidence suggesting that experiencing resource scarcity may sometimes lead to growth in interdependent self-views, empathy, and prosociality, which is sometimes referred to as “altruism-born-of-suffering”^62,63^. People who faced financial difficulties in childhood may develop a heightened sense of empathy and a strong desire to help others who are experiencing similar challenges, as a way to contribute to society while achieving a sense of meaning and personal growth^64^. This finding also indicates that social policies and practices that promote collaborative use of resources may enhance individuals’ effective coping with resource scarcity and foster an orientation to promote good.

Prior evidence has suggested associations of a religious upbringing with greater engagement in prosocial activities (e.g., volunteering, civic engagement) and higher levels of specific character strengths (e.g., forgiveness) in adulthood^37^. In line with this evidence, our study found that frequent religious service attendance during childhood was associated with a greater general disposition to promote good in adulthood, which followed a monotonic pattern on average across countries. This reflects that many faith traditions, through teachings like the Golden Rule, prioritize character building and the importance of acting for the benefit of others, fostering a lifelong commitment to promoting good^65^. A religious upbringing may reinforce the beliefs that character provides an important avenue leading to meaning, purpose, and fulfillment^36,37^. Amongst other potential explanations, attending structured rituals may also help foster certain character strengths such as perseverance, while also connecting individuals to a community with shared beliefs that in turn fortifies one’s personal values^66^. Notably, the positive association between childhood religious service attendance and promoting good in adulthood was observed even in some of the most secular societies in the world^59^ (e.g., Japan, Sweden, Spain, Germany). We speculate that individuals raised in religious families within these secular societies might hold particularly strong religious beliefs, which may lead to a deepened appreciation of character as a vital part of a fulfilled life.

Consistent with prior literature suggesting that character is likely shaped by both nature and nurture^67^, the pooled estimates across countries in this study found that positive relationships with both one’s mother and father were each positively associated with a disposition to promote good in adulthood. In contrast, experiences of child abuse and feeling like an outsider in the family were each inversely associated with promoting good. These dynamics may be partly explained by the Attachment Theory^31^. Specifically, love from caregivers fosters secure attachment, which helps children develop trust and self-worth. Children with secure attachment are likely to develop an internal working model that positively evaluates one’s own ability to care for oneself and others; meanwhile, they may also have more opportunities and capacity to address their own emotions and care for others^68^. Conversely, early-life experiences of neglect, abuse, and lack of belonging in the family may lead to insecure attachment^69^. This may result in low self-esteem, distrust, disrupted social relationships, and long-term health and well-being issues, which create barriers to exercising one’s strengths and character.

This study also adds novel evidence that, on average across countries, individuals who were older/from earlier birth cohorts reported a greater orientation to promote good. This finding is congruent with most prior literature on age differences in specific character strengths. For instance, a recent meta-analysis of data from 1,098,748 participants in English-speaking countries found that participants who were older/from earlier birth cohorts reported higher scores on most of the 24 character strengths (e.g., perseverance, fairness, kindness, gratitude, etc.)^66^. The Socioemotional Selectivity Theory suggests that as people age, they are increasingly aware of their limited time and tend to prioritize activities and goals that confer purpose and fulfillment^70^; the intention to promote good is one such activity that may generate meaning, accomplishment, and gratification. Additionally, researcher Erik Erikson hypothesized that people pass through eight stages of development across the lifespan, each marked by a specific conflict^27^. In later life, the stage of generativity versus stagnation emerges, where people strive to create or nurture things that will outlast them, such as contributing to society. This drive for generativity might also help explain the increased focus on promoting good as people age. It is, however, worth noting that the current analyses cannot distinguish between cohort and age effects, leaving it unclear whether the variations in orientation to promote good are mainly due to differences between birth cohorts or age groups. Country-specific analyses further revealed important variations across countries, suggesting that in high-income countries disposition to promote good is generally higher in participants from earlier birth cohorts/with older age. In contrast, the pattern was reversed in several lower-middle income countries. We hypothesize that such variations with birth cohorts/age groups may be attributed to different social trends and structural factors in these societies. For example, in some wealthy nations, such as the United States, young people are facing a mental health crisis and a declined sense of purpose in life^71^, which may diminish their social engagement, emotional resources, empathy, and sense of hope, in turn influencing their perception of the importance of promoting good and limiting their capacity to act for the greater good. In comparison, in some less economically developed countries older adults have limited support from the social protection system (e.g. limited access to public pensions, inadequate healthcare services, and insufficient long-term care options)^72^, which may result in structural barriers for their engagement in generative activities.

This study has several limitations that need to be considered. First, the adult participants recalled their childhood experiences simultaneously as when they reported on disposition to promote good, which may have led to recall bias. Nevertheless, for recall bias to completely explain away the observed associations will require that the effect of adult promoting good on biasing the recall of childhood experiences would have to be at least as strong as the observed associations themselves^73^, and many of these are quite substantial. Second, most of the constructs in this study were measured with single-item questions (due to the breadth of the topics covered in the Global Flourishing Study, the survey used single-item questions for many constructs^41^).

Therefore, potential measurement error needs to be considered when interpreting the results. Third, while disposition to promote good was assessed with an item from a previously validated measure^1^ and the survey translation closely followed relevant guidelines in cross-cultural survey research, some participants reported difficulties in understanding this question/translation in the GFS cognitive interviews^44^. This also suggests that the results may be subject to measurement error. Fourth, methodological complexities in cross-cultural research need to be considered. The cross-national variations in the findings may reflect true differences across countries, but they may also be attributed to the challenges in survey translations, cultural differences in response styles, and the seasonal variation in data collection. Fifth, because we included all childhood factors in the regression models simultaneously to examine their independent associations with promoting good, there may be collinearity between some of the childhood predictors (e.g., relationship with mother, relationship with father, marital status of parents, child abuse, feeling like an outsider in the family). While we have taken several approaches to reducing concerns about collinearity (e.g., removing some variables from the analyses; see our pre-registration), we cannot entirely rule out the possibility of collinearity. Sixth, due to lack of data this study cannot examine other childhood factors^32^ (e.g., temperament, peer relationships, school experience, neighborhood conditions) that may likewise shape one’s disposition to promote good or confound the associations between childhood antecedents and adult promoting good. Further studies that examine a comprehensive set of childhood antecedents of character and studies that focus on specific childhood predictors are much needed for developing a more nuanced understanding of character formation.

Character is often considered as an essential component of a flourishing life in and of itself, and expressing character by promoting good may also lead to favorable health and well-being outcomes^8,12^. Some strength-based interventions that aim to cultivate character strengths starting from early life have been developed, although the effect sizes of these interventions on enhancing character strengths and well-being outcomes are generally small to moderate^15,74,75^. Findings from the current study suggest that a wide array of individual, familial, societal, and cultural factors during childhood may be relevant to character formation. The findings also provide initial evidence that identifying and targeting the antecedents of character at multiple levels, including societal structural factors and cultural conditions, may inform more effective approaches to reducing disparities in access to resources and opportunities for character expression. To our knowledge, this is the first study that uses cross-national representative data to examine childhood factors that may predict adult character involving a disposition to promote good. Further studies are needed to deepen our understanding of how the disposition to promote good relates to actual engagement in acts of goodness, how childhood experiences shape character across social and cultural contexts, and of both universal and culturally specific approaches that can empower individuals and communities to cultivate strengths and fulfill their potential. Such knowledge may ultimately contribute to enhancing a more equitable distribution of population well-being and to promoting communal flourishing.

## Acknowledgements

The Global Flourishing Study was supported by funding from the John Templeton Foundation (grant #61665), Templeton Religion Trust (#1308), Templeton World Charity Foundation (#0605), Well-Being for Planet Earth Foundation, Fetzer Institute (#4354), Well Being Trust, Paul L. Foster Family Foundation, and the David and Carol Myers Foundation. The funding source had no impact on the study design; on the collection, analysis and interpretation of data; on the writing of the report; or on the decision to submit the article for publication.

## Author Contributions

B.R. J., and T.J.V. developed the study concept. Y.C., E.S.K., J.S.N., D.W.B., R.W., R.N.P., B.R. J., and T.J.V. contributed to the study design. Y.C. and R.N.P. had full access to the data, conducted data analyses, and take responsibility for the integrity of the data and accuracy of the data analysis. Y.C. drafted the manuscript. E.S.K., J.S.N., D.W.B., R.W., R.N.P., B.R. J., and T.J.V. provided critical revisions, and approved the final submitted version of the manuscript.

## Ethics approval and consent to participate

Ethical approval was granted by the institutional review boards at Baylor University (IRB Reference #: 6431841317) and Gallup (IRB Reference #: 2021-11-02), and all participants provided informed consent.

## Data Availability

Data for Wave 1 of the Global Flourishing Study is available through the Center for Open Science upon submission of a pre-registration (https://doi.org/10.17605/OSF.IO/3JTZ8), and will be openly available without pre-registration beginning February 2025. Please see https://www.cos.io/gfs-access-data for more information about data access.

## Code Availability

All analyses were pre-registered with COS prior to data access (https://doi.org/10.17605/OSF.IO/Y4U6W); All code to reproduce analyses are openly available in the online OSF repository (https://doi.org/10.17605/osf.io/vbype).

## Completing Interests

Tyler VanderWeele reports consulting fees from Gloo Inc., along with shared revenue received by Harvard University in its license agreement with Gloo according to the University IP policy. Other authors have no conflicts of interest to declare.

