## Supplementary file for "Early Life Experiences and Adult Orientation to Promote Good in 22 Countries"

- Table S1a – S22c: Supplementary tables for country-specific analyses.
- Table S23: Supplementary table for population weighted meta-analysis.
- Table S24: Supplementary table for population weighted meta-analysis of E-values.
- Figure S1– S27: Supplementary figures for estimates of childhood predictor categories.

***Supplementary Table S1A. Nationally Representative Childhood Descriptive Statistics for Argentina***

| **Variable** | **Category** | **N (%)** |
| --- | --- | --- |
| Relationship with mother | Very good | 4463 (66%) |
|  | Somewhat good | 1436 (21%) |
|  | Somewhat bad | 299 (4%) |
|  | Very bad | 216 (3%) |
|  | Does not apply | 273 (4%) |
|  | Missing | 36 (1%) |
| Relationships with father | Very good | 3612 (54%) |
|  | Somewhat good | 1537 (23%) |
|  | Somewhat bad | 440 (7%) |
|  | Very bad | 401 (6%) |
|  | Does not apply | 694 (10%) |
|  | Missing | 39 (1%) |
| Parent marital status | Parents married | 4110 (61%) |
|  | Divorced | 637 (9%) |
|  | Single, never married | 1368 (20%) |
|  | One or both parents had died | 199 (3%) |
|  | Missing | 410 (6%) |
| Subjective financial status of family growing up | Lived comfortably | 2042 (30%) |
|  | Got by | 2305 (34%) |
|  | Found it difficult | 1789 (27%) |
|  | Found it very difficult | 569 (8%) |
|  | Missing | 19 (0%) |
| Abuse | Yes | 1302 (19%) |
|  | No | 5271 (78%) |
|  | Missing | 151 (2%) |
| Felt like an outsider in the family | Yes | 1165 (17%) |
|  | No | 5458 (81%) |
|  | Missing | 101 (2%) |
| Self-rated health growing up | Excellent | 2402 (36%) |
|  | Very good | 1819 (27%) |
|  | Good | 1830 (27%) |
|  | Fair | 505 (8%) |
|  | Poor | 156 (2%) |
|  | Missing | 12 (0%) |
| Immigration status | Born in this country | 6346 (94%) |
|  | Born in another country | 348 (5%) |
|  | Missing | 29 (0%) |
| Age 12 religious service attendance | At least 1/week | 2601 (39%) |
|  | 1-3/month | 1204 (18%) |
|  | <1/ month | 1059 (16%) |
|  | Never | 1808 (27%) |
|  | Missing | 53 (1%) |
| Year of birth | 1998-2005; current age: 18-24 | 1108 (16%) |
|  | 1993-1998; age 25-29 | 719 (11%) |
|  | 1983-1993; age 30-39 | 1432 (21%) |
|  | 1973-1983; age 40-49 | 1254 (19%) |
|  | 1963-1973; age 50-59 | 1014 (15%) |
|  | 1953-1963; age 60-69 | 730 (11%) |
|  | 1943-1953; age 70-79 | 356 (5%) |
|  | 1943 or earlier; age 80+ | 112 (2%) |
| Gender | Male | 3143 (47%) |
|  | Female | 3542 (53%) |
|  | Other | 21 (0%) |
|  | Missing | 18 (0%) |
| Religious affiliation at age 12 | Christianity | 5805 (86%) |
|  | Islam | 11 (0%) |
|  | Hinduism | 2 (0%) |
|  | Buddhism | 3 (0%) |
|  | Judaism | 51 (1%) |
|  | Sikhism | 5 (0%) |
|  | Taoism | 1 (0%) |
|  | Primal, Animist, or Folk religion | 17 (0%) |
|  | Some other religion | 10 (0%) |
|  | No religion/Atheist/Agnostic | 697 (10%) |
|  | Missing | 122 (2%) |
| Race/ethnicity | Asian | 43 (1%) |
|  | Black | 95 (1%) |
|  | Indigenous | 129 (2%) |
|  | Mestizo(a) | 1801 (27%) |
|  | Mullato(a) | 75 (1%) |
|  | White | 3406 (51%) |
|  | Other | 104 (2%) |
|  | Missing | 1070 (16%) |

***Supplementary Table S1B. Regression of Promoting Good on Childhood Predictors for Argentina***

| **Variable** | **Category** | **Estimate** | **SE** | **95% CI** | **Global P-value** |
| --- | --- | --- | --- | --- | --- |
| Relationship with mother (ref: Very bad/Somewhat bad) | Very good/Somewhat good | 0.31 | 0.12 | (0.07,0.54) | 0.010 |
| Relationship with father (ref: Very bad/Somewhat bad) | Very good/Somewhat good | 0.09 | 0.10 | (-0.09,0.28) | 0.318 |
| Parent marital status (ref: Married) | Divorced | 0.09 | 0.10 | (-0.12,0.29) | 0.078 |
|  | Single, never married | 0.09 | 0.09 | (-0.08,0.27) |  |
|  | One or both parents had died | 0.33 | 0.15 | (0.03,0.63) |  |
| Subjective financial status of family growing up (ref: Got by) | Lived comfortably | 0.20 | 0.07 | (0.07,0.33) | <.001 |
|  | Found it difficult | 0.01 | 0.08 | (-0.14,0.16) |  |
|  | Found it very difficult | 0.42 | 0.12 | (0.18,0.66) |  |
| Abuse (ref: No) | Yes | -0.07 | 0.08 | (-0.22,0.08) | 0.338 |
| Felt like an outsider in the family (ref: No) | Yes | -0.05 | 0.09 | (-0.23,0.13) | 0.550 |
| Self-rated health growing up (ref: Good) | Excellent | 0.45 | 0.07 | (0.31,0.60) | <.001 |
|  | Very good | 0.08 | 0.08 | (-0.07,0.24) |  |
|  | Fair | 0.25 | 0.12 | (0.01,0.50) |  |
|  | Poor | -0.35 | 0.29 | (-0.92,0.22) |  |
| Immigration status (ref: Born in this country) | Born in another country | 0.03 | 0.13 | (-0.23,0.29) | 0.776 |
| Age 12 religious service attendance (ref: Never) | At least 1/week | 0.08 | 0.08 | (-0.06,0.23) | 0.008 |
|  | 1-3/month | -0.08 | 0.09 | (-0.25,0.09) |  |
|  | <1/month | -0.19 | 0.10 | (-0.38,-0.00) |  |
| Birth year (ref: 1998-2005; current age: 18-24) | 1993-1998; age 25-29 | 0.16 | 0.12 | (-0.08,0.40) | <.001 |
|  | 1983-1993; age 30-39 | 0.20 | 0.10 | (-0.00,0.40) |  |
|  | 1973-1983; age 40-49 | 0.42 | 0.10 | (0.22,0.61) |  |
|  | 1963-1973; age 50-59 | 0.63 | 0.10 | (0.43,0.83) |  |
|  | 1953-1963; age 60-69 | 0.41 | 0.12 | (0.18,0.65) |  |
|  | 1943-1953; age 70-79 | 0.39 | 0.16 | (0.08,0.70) |  |
|  | 1943 or earlier; age 80+ | 0.21 | 0.18 | (-0.15,0.57) |  |
| Gender (ref:Male) | Female | 0.24 | 0.06 | (0.13,0.35) | <.001 |
|  | Other | 0.22 | 0.39 | (-0.55,1.00) |  |
| Religious affiliation at age 12 (ref: No religion/Atheist/Agnostic) | Christianity | 0.08 | 0.12 | (-0.15,0.32) | 0.655 |
|  | Collapsed affiliations with prevalence<3% | -0.04 | 0.25 | (-0.53,0.44) |  |
| Race/ethnicity (ref: plurality) | Race/ethnicity minority | -0.06 | 0.06 | (-0.18,0.07) | 0.306 |

***Supplementary Table S1C. E-Values for Estimates and CI for Argentina***

| **Variable** | **Category** | **E-Value for Estimate** | **E-Value for 95% CI** |
| --- | --- | --- | --- |
| Relationship with mother (ref: Very bad/Somewhat bad) | Very good/Somewhat good | 1.65 | 1.25 |
| Relationship with father (ref: Very bad/Somewhat bad) | Very good/Somewhat good | 1.29 | 1.00 |
| Parent marital status (ref: Married) | Divorced | 1.27 | 1.00 |
|  | Single, never married | 1.29 | 1.00 |
|  | One or both parents had died | 1.69 | 1.15 |
| Subjective financial status of family growing up (ref: Got by) | Lived comfortably | 1.48 | 1.23 |
|  | Found it difficult | 1.07 | 1.00 |
|  | Found it very difficult | 1.83 | 1.44 |
| Abuse (ref: No) | Yes | 1.25 | 1.00 |
| Felt like an outsider in the family (ref: No) | Yes | 1.19 | 1.00 |
| Self-rated health growing up (ref: Good) | Excellent | 1.89 | 1.66 |
|  | Very good | 1.27 | 1.00 |
|  | Fair | 1.57 | 1.08 |
|  | Poor | 1.72 | 1.00 |
| Immigration status (ref: Born in this country) | Born in another country | 1.15 | 1.00 |
| Age 12 religious service attendance (ref: Never) | At least 1/week | 1.27 | 1.00 |
|  | 1-3/month | 1.27 | 1.00 |
|  | <1/month | 1.47 | 1.03 |
| Birth year (ref: 1998-2005; current age: 18-24) | 1993-1998; age 25-29 | 1.41 | 1.00 |
|  | 1983-1993; age 30-39 | 1.48 | 1.00 |
|  | 1973-1983; age 40-49 | 1.83 | 1.52 |
|  | 1963-1973; age 50-59 | 2.18 | 1.85 |
|  | 1953-1963; age 60-69 | 1.83 | 1.44 |
|  | 1943-1953; age 70-79 | 1.78 | 1.26 |
|  | 1943 or earlier; age 80+ | 1.50 | 1.00 |
| Gender (ref:Male) | Female | 1.55 | 1.35 |
|  | Other | 1.52 | 1.00 |
| Religious affiliation at age 12 (ref: No religion/Atheist/Agnostic) | Christianity | 1.27 | 1.00 |
|  | Collapsed affiliations with prevalence<3% | 1.19 | 1.00 |
| Race/ethnicity (ref: plurality) | Race/ethnicity minority | 1.21 | 1.00 |

***Supplementary Table S2A. Nationally Representative Childhood Descriptive Statistics for Australia***

| **Variable** | **Category** | **N (%)** |
| --- | --- | --- |
| Relationship with mother | Very good | 2554 (66%) |
|  | Somewhat good | 925 (24%) |
|  | Somewhat bad | 218 (6%) |
|  | Very bad | 107 (3%) |
|  | Does not apply | 32 (1%) |
|  | Missing | 7 (0%) |
| Relationships with father | Very good | 2032 (53%) |
|  | Somewhat good | 1144 (30%) |
|  | Somewhat bad | 315 (8%) |
|  | Very bad | 196 (5%) |
|  | Does not apply | 148 (4%) |
|  | Missing | 9 (0%) |
| Parent marital status | Parents married | 3048 (79%) |
|  | Divorced | 462 (12%) |
|  | Single, never married | 187 (5%) |
|  | One or both parents had died | 96 (2%) |
|  | Missing | 52 (1%) |
| Subjective financial status of family growing up | Lived comfortably | 1756 (46%) |
|  | Got by | 1496 (39%) |
|  | Found it difficult | 422 (11%) |
|  | Found it very difficult | 154 (4%) |
|  | Missing | 16 (0%) |
| Abuse | Yes | 995 (26%) |
|  | No | 2790 (73%) |
|  | Missing | 59 (2%) |
| Felt like an outsider in the family | Yes | 756 (20%) |
|  | No | 3062 (80%) |
|  | Missing | 26 (1%) |
| Self-rated health growing up | Excellent | 1736 (45%) |
|  | Very good | 1087 (28%) |
|  | Good | 603 (16%) |
|  | Fair | 308 (8%) |
|  | Poor | 106 (3%) |
|  | Missing | 4 (0%) |
| Immigration status | Born in this country | 2953 (77%) |
|  | Born in another country | 885 (23%) |
|  | Missing | 6 (0%) |
| Age 12 religious service attendance | At least 1/week | 1362 (35%) |
|  | 1-3/month | 486 (13%) |
|  | <1/ month | 600 (16%) |
|  | Never | 1307 (34%) |
|  | Missing | 90 (2%) |
| Year of birth | 1998-2005; current age: 18-24 | 345 (9%) |
|  | 1993-1998; age 25-29 | 282 (7%) |
|  | 1983-1993; age 30-39 | 641 (17%) |
|  | 1973-1983; age 40-49 | 618 (16%) |
|  | 1963-1973; age 50-59 | 691 (18%) |
|  | 1953-1963; age 60-69 | 589 (15%) |
|  | 1943-1953; age 70-79 | 498 (13%) |
|  | 1943 or earlier; age 80+ | 178 (5%) |
|  | Missing | 2 (0%) |
| Gender | Male | 1861 (48%) |
|  | Female | 1941 (50%) |
|  | Other | 36 (1%) |
|  | Missing | 6 (0%) |
| Religious affiliation at age 12 | Christianity | 2678 (70%) |
|  | Islam | 48 (1%) |
|  | Hinduism | 39 (1%) |
|  | Buddhism | 16 (0%) |
|  | Judaism | 29 (1%) |
|  | Sikhism | 6 (0%) |
|  | Baha’i | 5 (0%) |
|  | Taoism | 1 (0%) |
|  | Primal, Animist, or Folk religion | 4 (0%) |
|  | Some other religion | 8 (0%) |
|  | No religion/Atheist/Agnostic | 990 (26%) |
|  | Missing | 21 (1%) |
| Race/ethnicity | Aboriginal | 53 (1%) |
|  | Australian | 1946 (51%) |
|  | Australian British/European | 1047 (27%) |
|  | Chinese | 75 (2%) |
|  | Indian | 58 (2%) |
|  | Japanese | 1 (0%) |
|  | Malay | 11 (0%) |
|  | Sinhalese | 1 (0%) |
|  | Spanish | 2 (0%) |
|  | Sri Lankan Moor | 1 (0%) |
|  | Sri Lankan Tamil | 7 (0%) |
|  | Vietnamese | 7 (0%) |
|  | Russian | 7 (0%) |
|  | Samoan | 4 (0%) |
|  | New Zealander | 91 (2%) |
|  | Other European | 357 (9%) |
|  | Other | 163 (4%) |
|  | Missing | 14 (0%) |

***Supplementary Table S2B. Regression of Promoting Good on Childhood Predictors for Australia***

| **Variable** | **Category** | **Estimate** | **SE** | **95% CI** | **Global P-value** |
| --- | --- | --- | --- | --- | --- |
| Relationship with mother (ref: Very bad/Somewhat bad) | Very good/Somewhat good | -0.02 | 0.15 | (-0.32,0.28) | 0.895 |
| Relationship with father (ref: Very bad/Somewhat bad) | Very good/Somewhat good | 0.11 | 0.12 | (-0.12,0.34) | 0.353 |
| Parent marital status (ref: Married) | Divorced | 0.19 | 0.12 | (-0.04,0.42) | <.001 |
|  | Single, never married | 0.75 | 0.19 | (0.38,1.11) |  |
|  | One or both parents had died | 0.54 | 0.19 | (0.16,0.91) |  |
| Subjective financial status of family growing up (ref: Got by) | Lived comfortably | 0.07 | 0.07 | (-0.07,0.21) | 0.352 |
|  | Found it difficult | -0.13 | 0.12 | (-0.37,0.10) |  |
|  | Found it very difficult | -0.04 | 0.20 | (-0.44,0.35) |  |
| Abuse (ref: No) | Yes | -0.09 | 0.09 | (-0.27,0.08) | 0.287 |
| Felt like an outsider in the family (ref: No) | Yes | -0.21 | 0.11 | (-0.42,-0.00) | 0.046 |
| Self-rated health growing up (ref: Good) | Excellent | 0.43 | 0.10 | (0.24,0.62) | <.001 |
|  | Very good | 0.04 | 0.10 | (-0.16,0.25) |  |
|  | Fair | 0.06 | 0.16 | (-0.24,0.37) |  |
|  | Poor | -0.02 | 0.28 | (-0.57,0.54) |  |
| Immigration status (ref: Born in this country) | Born in another country | -0.01 | 0.09 | (-0.18,0.16) | 0.878 |
| Age 12 religious service attendance (ref: Never) | At least 1/week | 0.21 | 0.09 | (0.02,0.39) | 0.008 |
|  | 1-3/month | 0.04 | 0.11 | (-0.17,0.25) |  |
|  | <1/month | -0.14 | 0.11 | (-0.36,0.09) |  |
| Birth year (ref: 1998-2005; current age: 18-24) | 1993-1998; age 25-29 | 0.29 | 0.21 | (-0.12,0.70) | <.001 |
|  | 1983-1993; age 30-39 | 0.50 | 0.18 | (0.15,0.84) |  |
|  | 1973-1983; age 40-49 | 0.66 | 0.17 | (0.32,0.99) |  |
|  | 1963-1973; age 50-59 | 0.51 | 0.17 | (0.18,0.85) |  |
|  | 1953-1963; age 60-69 | 0.89 | 0.17 | (0.56,1.22) |  |
|  | 1943-1953; age 70-79 | 1.17 | 0.18 | (0.83,1.52) |  |
|  | 1943 or earlier; age 80+ | 1.27 | 0.18 | (0.91,1.63) |  |
| Gender (ref:Male) | Female | 0.05 | 0.07 | (-0.08,0.18) | 0.672 |
|  | Other | -0.11 | 0.39 | (-0.88,0.66) |  |
| Religious affiliation at age 12 (ref: No religion/Atheist/Agnostic) | Christianity | -0.06 | 0.10 | (-0.24,0.13) | 0.829 |
|  | Collapsed affiliations with prevalence<3% | 0.00 | 0.21 | (-0.42,0.42) |  |
| Race/ethnicity (ref: plurality) | Race/ethnicity minority | 0.07 | 0.07 | (-0.08,0.21) | 0.347 |

***Supplementary Table S2C. E-Values for Estimates and CI for Australia***

| **Variable** | **Category** | **E-Value for Estimate** | **E-Value for 95% CI** |
| --- | --- | --- | --- |
| Relationship with mother (ref: Very bad/Somewhat bad) | Very good/Somewhat good | 1.10 | 1.00 |
| Relationship with father (ref: Very bad/Somewhat bad) | Very good/Somewhat good | 1.33 | 1.00 |
| Parent marital status (ref: Married) | Divorced | 1.47 | 1.00 |
|  | Single, never married | 2.42 | 1.79 |
|  | One or both parents had died | 2.05 | 1.43 |
| Subjective financial status of family growing up (ref: Got by) | Lived comfortably | 1.24 | 1.00 |
|  | Found it difficult | 1.37 | 1.00 |
|  | Found it very difficult | 1.18 | 1.00 |
| Abuse (ref: No) | Yes | 1.29 | 1.00 |
| Felt like an outsider in the family (ref: No) | Yes | 1.50 | 1.04 |
| Self-rated health growing up (ref: Good) | Excellent | 1.87 | 1.56 |
|  | Very good | 1.18 | 1.00 |
|  | Fair | 1.23 | 1.00 |
|  | Poor | 1.12 | 1.00 |
| Immigration status (ref: Born in this country) | Born in another country | 1.09 | 1.00 |
| Age 12 religious service attendance (ref: Never) | At least 1/week | 1.50 | 1.13 |
|  | 1-3/month | 1.17 | 1.00 |
|  | <1/month | 1.38 | 1.00 |
| Birth year (ref: 1998-2005; current age: 18-24) | 1993-1998; age 25-29 | 1.63 | 1.00 |
|  | 1983-1993; age 30-39 | 1.98 | 1.40 |
|  | 1973-1983; age 40-49 | 2.25 | 1.68 |
|  | 1963-1973; age 50-59 | 2.01 | 1.45 |
|  | 1953-1963; age 60-69 | 2.69 | 2.08 |
|  | 1943-1953; age 70-79 | 3.29 | 2.57 |
|  | 1943 or earlier; age 80+ | 3.51 | 2.73 |
| Gender (ref:Male) | Female | 1.21 | 1.00 |
|  | Other | 1.32 | 1.00 |
| Religious affiliation at age 12 (ref: No religion/Atheist/Agnostic) | Christianity | 1.21 | 1.00 |
|  | Collapsed affiliations with prevalence<3% | 1.02 | 1.00 |
| Race/ethnicity (ref: plurality) | Race/ethnicity minority | 1.24 | 1.00 |

***Supplementary Table S3A. Nationally Representative Childhood Descriptive Statistics for Brazil***

| **Variable** | **Category** | **N (%)** |
| --- | --- | --- |
| Relationship with mother | Very good | 8369 (63%) |
|  | Somewhat good | 3559 (27%) |
|  | Somewhat bad | 483 (4%) |
|  | Very bad | 214 (2%) |
|  | Does not apply | 507 (4%) |
|  | Missing | 73 (1%) |
| Relationships with father | Very good | 6364 (48%) |
|  | Somewhat good | 3654 (28%) |
|  | Somewhat bad | 1035 (8%) |
|  | Very bad | 756 (6%) |
|  | Does not apply | 1303 (10%) |
|  | Missing | 93 (1%) |
| Parent marital status | Parents married | 8546 (65%) |
|  | Divorced | 1384 (10%) |
|  | Single, never married | 1985 (15%) |
|  | One or both parents had died | 508 (4%) |
|  | Missing | 781 (6%) |
| Subjective financial status of family growing up | Lived comfortably | 4998 (38%) |
|  | Got by | 4616 (35%) |
|  | Found it difficult | 2484 (19%) |
|  | Found it very difficult | 1027 (8%) |
|  | Missing | 79 (1%) |
| Abuse | Yes | 2606 (20%) |
|  | No | 10147 (77%) |
|  | Missing | 451 (3%) |
| Felt like an outsider in the family | Yes | 1659 (13%) |
|  | No | 11234 (85%) |
|  | Missing | 311 (2%) |
| Self-rated health growing up | Excellent | 5312 (40%) |
|  | Very good | 3392 (26%) |
|  | Good | 2873 (22%) |
|  | Fair | 1368 (10%) |
|  | Poor | 228 (2%) |
|  | Missing | 30 (0%) |
| Immigration status | Born in this country | 12688 (96%) |
|  | Born in another country | 153 (1%) |
|  | Missing | 363 (3%) |
| Age 12 religious service attendance | At least 1/week | 6306 (48%) |
|  | 1-3/month | 2491 (19%) |
|  | <1/ month | 2629 (20%) |
|  | Never | 1707 (13%) |
|  | Missing | 71 (1%) |
| Year of birth | 1998-2005; current age: 18-24 | 1986 (15%) |
|  | 1993-1998; age 25-29 | 1468 (11%) |
|  | 1983-1993; age 30-39 | 2908 (22%) |
|  | 1973-1983; age 40-49 | 2638 (20%) |
|  | 1963-1973; age 50-59 | 2131 (16%) |
|  | 1953-1963; age 60-69 | 1435 (11%) |
|  | 1943-1953; age 70-79 | 510 (4%) |
|  | 1943 or earlier; age 80+ | 126 (1%) |
| Gender | Male | 6320 (48%) |
|  | Female | 6820 (52%) |
|  | Other | 35 (0%) |
|  | Missing | 30 (0%) |
| Religious affiliation at age 12 | Christianity | 11403 (86%) |
|  | Islam | 15 (0%) |
|  | Hinduism | 1 (0%) |
|  | Buddhism | 27 (0%) |
|  | Judaism | 40 (0%) |
|  | Baha’i | 1 (0%) |
|  | Jainism | 4 (0%) |
|  | Shinto | 4 (0%) |
|  | Taoism | 1 (0%) |
|  | Confucianism | 7 (0%) |
|  | Primal, Animist, or Folk religion | 17 (0%) |
|  | Spiritism | 336 (3%) |
|  | Umbanda, Candomblé, and other African-derived religions | 262 (2%) |
|  | Some other religion | 87 (1%) |
|  | No religion/Atheist/Agnostic | 908 (7%) |
|  | Missing | 94 (1%) |
| Race/ethnicity | Branca | 5169 (39%) |
|  | Preta | 1615 (12%) |
|  | Parda | 5125 (39%) |
|  | Amarela | 238 (2%) |
|  | Indígena | 131 (1%) |
|  | Other | 61 (0%) |
|  | Missing | 865 (7%) |

***Supplementary Table S3B. Regression of Promoting Good on Childhood Predictors for Brazil***

| **Variable** | **Category** | **Estimate** | **SE** | **95% CI** | **Global P-value** |
| --- | --- | --- | --- | --- | --- |
| Relationship with mother (ref: Very bad/Somewhat bad) | Very good/Somewhat good | 0.15 | 0.10 | (-0.04,0.35) | 0.113 |
| Relationship with father (ref: Very bad/Somewhat bad) | Very good/Somewhat good | 0.16 | 0.06 | (0.04,0.29) | 0.008 |
| Parent marital status (ref: Married) | Divorced | -0.04 | 0.07 | (-0.17,0.09) | 0.671 |
|  | Single, never married | -0.06 | 0.07 | (-0.21,0.08) |  |
|  | One or both parents had died | -0.10 | 0.13 | (-0.35,0.15) |  |
| Subjective financial status of family growing up (ref: Got by) | Lived comfortably | 0.06 | 0.05 | (-0.03,0.15) | 0.210 |
|  | Found it difficult | 0.02 | 0.06 | (-0.10,0.15) |  |
|  | Found it very difficult | 0.19 | 0.10 | (-0.00,0.38) |  |
| Abuse (ref: No) | Yes | -0.05 | 0.06 | (-0.16,0.06) | 0.403 |
| Felt like an outsider in the family (ref: No) | Yes | -0.22 | 0.08 | (-0.37,-0.07) | 0.004 |
| Self-rated health growing up (ref: Good) | Excellent | 0.55 | 0.06 | (0.44,0.67) | <.001 |
|  | Very good | 0.15 | 0.06 | (0.03,0.27) |  |
|  | Fair | -0.13 | 0.10 | (-0.33,0.06) |  |
|  | Poor | 0.19 | 0.16 | (-0.13,0.50) |  |
| Immigration status (ref: Born in this country) | Born in another country | -0.48 | 0.24 | (-0.96,-0.01) | 0.028 |
| Age 12 religious service attendance (ref: Never) | At least 1/week | 0.31 | 0.08 | (0.16,0.46) | <.001 |
|  | 1-3/month | 0.21 | 0.09 | (0.04,0.37) |  |
|  | <1/month | 0.08 | 0.09 | (-0.09,0.25) |  |
| Birth year (ref: 1998-2005; current age: 18-24) | 1993-1998; age 25-29 | 0.23 | 0.09 | (0.06,0.40) | <.001 |
|  | 1983-1993; age 30-39 | 0.42 | 0.07 | (0.28,0.55) |  |
|  | 1973-1983; age 40-49 | 0.53 | 0.07 | (0.38,0.67) |  |
|  | 1963-1973; age 50-59 | 0.72 | 0.08 | (0.57,0.87) |  |
|  | 1953-1963; age 60-69 | 0.73 | 0.09 | (0.55,0.91) |  |
|  | 1943-1953; age 70-79 | 0.53 | 0.18 | (0.17,0.88) |  |
|  | 1943 or earlier; age 80+ | 0.88 | 0.21 | (0.47,1.30) |  |
| Gender (ref:Male) | Female | 0.04 | 0.04 | (-0.04,0.12) | 0.159 |
|  | Other | -1.01 | 0.63 | (-2.25,0.24) |  |
| Religious affiliation at age 12 (ref: No religion/Atheist/Agnostic) | Christianity | 0.08 | 0.10 | (-0.11,0.27) | 0.604 |
|  | Collapsed affiliations with prevalence<3% | 0.04 | 0.14 | (-0.23,0.31) |  |
| Race/ethnicity (ref: plurality) | Race/ethnicity minority | 0.12 | 0.04 | (0.03,0.20) | 0.007 |

***Supplementary Table S3C. E-Values for Estimates and CI for Brazil***

| **Variable** | **Category** | **E-Value for Estimate** | **E-Value for 95% CI** |
| --- | --- | --- | --- |
| Relationship with mother (ref: Very bad/Somewhat bad) | Very good/Somewhat good | 1.38 | 1.00 |
| Relationship with father (ref: Very bad/Somewhat bad) | Very good/Somewhat good | 1.39 | 1.16 |
| Parent marital status (ref: Married) | Divorced | 1.16 | 1.00 |
|  | Single, never married | 1.22 | 1.00 |
|  | One or both parents had died | 1.29 | 1.00 |
| Subjective financial status of family growing up (ref: Got by) | Lived comfortably | 1.21 | 1.00 |
|  | Found it difficult | 1.12 | 1.00 |
|  | Found it very difficult | 1.43 | 1.00 |
| Abuse (ref: No) | Yes | 1.18 | 1.00 |
| Felt like an outsider in the family (ref: No) | Yes | 1.48 | 1.22 |
| Self-rated health growing up (ref: Good) | Excellent | 1.98 | 1.81 |
|  | Very good | 1.37 | 1.13 |
|  | Fair | 1.35 | 1.00 |
|  | Poor | 1.43 | 1.00 |
| Immigration status (ref: Born in this country) | Born in another country | 1.87 | 1.10 |
| Age 12 religious service attendance (ref: Never) | At least 1/week | 1.62 | 1.39 |
|  | 1-3/month | 1.46 | 1.17 |
|  | <1/month | 1.25 | 1.00 |
| Birth year (ref: 1998-2005; current age: 18-24) | 1993-1998; age 25-29 | 1.49 | 1.21 |
|  | 1983-1993; age 30-39 | 1.78 | 1.58 |
|  | 1973-1983; age 40-49 | 1.94 | 1.73 |
|  | 1963-1973; age 50-59 | 2.24 | 2.00 |
|  | 1953-1963; age 60-69 | 2.26 | 1.98 |
|  | 1943-1953; age 70-79 | 1.94 | 1.41 |
|  | 1943 or earlier; age 80+ | 2.51 | 1.85 |
| Gender (ref:Male) | Female | 1.17 | 1.00 |
|  | Other | 2.72 | 1.00 |
| Religious affiliation at age 12 (ref: No religion/Atheist/Agnostic) | Christianity | 1.25 | 1.00 |
|  | Collapsed affiliations with prevalence<3% | 1.16 | 1.00 |
| Race/ethnicity (ref: plurality) | Race/ethnicity minority | 1.32 | 1.15 |

***Supplementary Table S4A. Nationally Representative Childhood Descriptive Statistics for Egypt***

| **Variable** | **Category** | **N (%)** |
| --- | --- | --- |
| Relationship with mother | Very good | 4110 (87%) |
|  | Somewhat good | 505 (11%) |
|  | Somewhat bad | 21 (0%) |
|  | Very bad | 10 (0%) |
|  | Does not apply | 83 (2%) |
| Relationships with father | Very good | 3713 (79%) |
|  | Somewhat good | 683 (14%) |
|  | Somewhat bad | 56 (1%) |
|  | Very bad | 30 (1%) |
|  | Does not apply | 233 (5%) |
|  | Missing | 14 (0%) |
| Parent marital status | Parents married | 4049 (86%) |
|  | Divorced | 131 (3%) |
|  | Single, never married | 9 (0%) |
|  | One or both parents had died | 485 (10%) |
|  | Missing | 55 (1%) |
| Subjective financial status of family growing up | Lived comfortably | 1251 (26%) |
|  | Got by | 2352 (50%) |
|  | Found it difficult | 857 (18%) |
|  | Found it very difficult | 268 (6%) |
|  | Missing | 1 (0%) |
| Abuse | Yes | 405 (9%) |
|  | No | 4293 (91%) |
|  | Missing | 30 (1%) |
| Felt like an outsider in the family | Yes | 260 (5%) |
|  | No | 4456 (94%) |
|  | Missing | 13 (0%) |
| Self-rated health growing up | Excellent | 2687 (57%) |
|  | Very good | 1174 (25%) |
|  | Good | 497 (11%) |
|  | Fair | 265 (6%) |
|  | Poor | 106 (2%) |
|  | Missing | 1 (0%) |
| Immigration status | Born in this country | 4713 (100%) |
|  | Born in another country | 16 (0%) |
|  | Missing | 1 (0%) |
| Age 12 religious service attendance | At least 1/week | 2307 (49%) |
|  | 1-3/month | 570 (12%) |
|  | <1/ month | 629 (13%) |
|  | Never | 1165 (25%) |
|  | Missing | 57 (1%) |
| Year of birth | 1998-2005; current age: 18-24 | 960 (20%) |
|  | 1993-1998; age 25-29 | 607 (13%) |
|  | 1983-1993; age 30-39 | 1204 (25%) |
|  | 1973-1983; age 40-49 | 897 (19%) |
|  | 1963-1973; age 50-59 | 613 (13%) |
|  | 1953-1963; age 60-69 | 387 (8%) |
|  | 1943-1953; age 70-79 | 54 (1%) |
|  | 1943 or earlier; age 80+ | 7 (0%) |
| Gender | Male | 2394 (51%) |
|  | Female | 2334 (49%) |
|  | Missing | 0 (0%) |
| Religious affiliation at age 12 | Christianity | 123 (3%) |
|  | Islam | 4602 (97%) |
|  | Jainism | 1 (0%) |
|  | Taoism | 0 (0%) |
|  | Missing | 3 (0%) |
| Race/ethnicity | Arab | 4585 (97%) |
|  | Turkish | 9 (0%) |
|  | Greek | 1 (0%) |
|  | Bedouin Arab | 4 (0%) |
|  | Nubian | 27 (1%) |
|  | Missing | 102 (2%) |

***Supplementary Table S4B. Regression of Promoting Good on Childhood Predictors for Egypt***

| **Variable** | **Category** | **Estimate** | **SE** | **95% CI** | **Global P-value** |
| --- | --- | --- | --- | --- | --- |
| Relationship with mother (ref: Very bad/Somewhat bad) | Very good/Somewhat good | -0.71 | 0.31 | (-1.33,-0.10) | 0.024 |
| Relationship with father (ref: Very bad/Somewhat bad) | Very good/Somewhat good | -0.07 | 0.24 | (-0.54,0.40) | 0.771 |
| Parent marital status (ref: Married) | Divorced | 0.13 | 0.26 | (-0.40,0.65) | 0.837 |
|  | Single, never married | 0.49 | 0.93 | (-1.36,2.34) |  |
|  | One or both parents had died | 0.02 | 0.16 | (-0.30,0.33) |  |
| Subjective financial status of family growing up (ref: Got by) | Lived comfortably | 0.12 | 0.08 | (-0.05,0.28) | 0.408 |
|  | Found it difficult | 0.11 | 0.12 | (-0.11,0.34) |  |
|  | Found it very difficult | -0.07 | 0.18 | (-0.43,0.28) |  |
| Abuse (ref: No) | Yes | 0.05 | 0.14 | (-0.23,0.32) | 0.708 |
| Felt like an outsider in the family (ref: No) | Yes | -0.21 | 0.20 | (-0.60,0.18) | 0.280 |
| Self-rated health growing up (ref: Good) | Excellent | 0.10 | 0.13 | (-0.17,0.36) | 0.059 |
|  | Very good | 0.08 | 0.15 | (-0.21,0.38) |  |
|  | Fair | -0.30 | 0.21 | (-0.71,0.11) |  |
|  | Poor | 0.39 | 0.23 | (-0.07,0.86) |  |
| Immigration status (ref: Born in this country) | Born in another country | -0.58 | 0.65 | (-1.89,0.72) | 0.298 |
| Age 12 religious service attendance (ref: Never) | At least 1/week | 0.51 | 0.11 | (0.29,0.73) | <.001 |
|  | 1-3/month | 0.27 | 0.14 | (-0.01,0.55) |  |
|  | <1/month | 0.32 | 0.12 | (0.08,0.57) |  |
| Birth year (ref: 1998-2005; current age: 18-24) | 1993-1998; age 25-29 | -0.01 | 0.13 | (-0.27,0.26) | 0.219 |
|  | 1983-1993; age 30-39 | -0.09 | 0.11 | (-0.31,0.13) |  |
|  | 1973-1983; age 40-49 | 0.07 | 0.11 | (-0.15,0.30) |  |
|  | 1963-1973; age 50-59 | -0.01 | 0.15 | (-0.30,0.27) |  |
|  | 1953-1963; age 60-69 | -0.27 | 0.20 | (-0.66,0.11) |  |
|  | 1943-1953; age 70-79 | -0.74 | 0.44 | (-1.62,0.14) |  |
|  | 1943 or earlier; age 80+ | -0.30 | 0.30 | (-0.89,0.29) |  |
| Gender (ref:Male) | Female | 0.28 | 0.10 | (0.10,0.47) | 0.003 |
| Religious affiliation at age 12 (ref: Islam) | Collapsed affiliations with prevalence<3% | -0.18 | 0.19 | (-0.56,0.20) | 0.318 |
| Race/ethnicity (ref: plurality) | Race/ethnicity minority | 0.18 | 0.35 | (-0.52,0.88) | 0.589 |

***Supplementary Table S4C. E-Values for Estimates and CI for Egypt***

| **Variable** | **Category** | **E-Value for Estimate** | **E-Value for 95% CI** |
| --- | --- | --- | --- |
| Relationship with mother (ref: Very bad/Somewhat bad) | Very good/Somewhat good | 2.07 | 1.27 |
| Relationship with father (ref: Very bad/Somewhat bad) | Very good/Somewhat good | 1.20 | 1.00 |
| Parent marital status (ref: Married) | Divorced | 1.30 | 1.00 |
|  | Single, never married | 1.78 | 1.00 |
|  | One or both parents had died | 1.09 | 1.00 |
| Subjective financial status of family growing up (ref: Got by) | Lived comfortably | 1.29 | 1.00 |
|  | Found it difficult | 1.28 | 1.00 |
|  | Found it very difficult | 1.22 | 1.00 |
| Abuse (ref: No) | Yes | 1.17 | 1.00 |
| Felt like an outsider in the family (ref: No) | Yes | 1.42 | 1.00 |
| Self-rated health growing up (ref: Good) | Excellent | 1.26 | 1.00 |
|  | Very good | 1.23 | 1.00 |
|  | Fair | 1.54 | 1.00 |
|  | Poor | 1.66 | 1.00 |
| Immigration status (ref: Born in this country) | Born in another country | 1.90 | 1.00 |
| Age 12 religious service attendance (ref: Never) | At least 1/week | 1.80 | 1.53 |
|  | 1-3/month | 1.50 | 1.00 |
|  | <1/month | 1.57 | 1.23 |
| Birth year (ref: 1998-2005; current age: 18-24) | 1993-1998; age 25-29 | 1.06 | 1.00 |
|  | 1983-1993; age 30-39 | 1.24 | 1.00 |
|  | 1973-1983; age 40-49 | 1.21 | 1.00 |
|  | 1963-1973; age 50-59 | 1.08 | 1.00 |
|  | 1953-1963; age 60-69 | 1.50 | 1.00 |
|  | 1943-1953; age 70-79 | 2.11 | 1.00 |
|  | 1943 or earlier; age 80+ | 1.54 | 1.00 |
| Gender (ref:Male) | Female | 1.52 | 1.26 |
| Religious affiliation at age 12 (ref: Islam) | Collapsed affiliations with prevalence<3% | 1.38 | 1.00 |
| Race/ethnicity (ref: plurality) | Race/ethnicity minority | 1.38 | 1.00 |

***Supplementary Table S5A. Nationally Representative Childhood Descriptive Statistics for Germany***

| **Variable** | **Category** | **N (%)** |
| --- | --- | --- |
| Relationship with mother | Very good | 5497 (58%) |
|  | Somewhat good | 3031 (32%) |
|  | Somewhat bad | 496 (5%) |
|  | Very bad | 187 (2%) |
|  | Does not apply | 241 (3%) |
|  | Missing | 54 (1%) |
| Relationships with father | Very good | 4652 (49%) |
|  | Somewhat good | 3012 (32%) |
|  | Somewhat bad | 846 (9%) |
|  | Very bad | 385 (4%) |
|  | Does not apply | 538 (6%) |
|  | Missing | 73 (1%) |
| Parent marital status | Parents married | 7620 (80%) |
|  | Divorced | 927 (10%) |
|  | Single, never married | 578 (6%) |
|  | One or both parents had died | 245 (3%) |
|  | Missing | 136 (1%) |
| Subjective financial status of family growing up | Lived comfortably | 3177 (33%) |
|  | Got by | 4508 (47%) |
|  | Found it difficult | 1481 (16%) |
|  | Found it very difficult | 314 (3%) |
|  | Missing | 26 (0%) |
| Abuse | Yes | 1086 (11%) |
|  | No | 8321 (88%) |
|  | Missing | 99 (1%) |
| Felt like an outsider in the family | Yes | 1105 (12%) |
|  | No | 8262 (87%) |
|  | Missing | 139 (1%) |
| Self-rated health growing up | Excellent | 2633 (28%) |
|  | Very good | 3518 (37%) |
|  | Good | 2582 (27%) |
|  | Fair | 612 (6%) |
|  | Poor | 134 (1%) |
|  | Missing | 26 (0%) |
| Immigration status | Born in this country | 8722 (92%) |
|  | Born in another country | 744 (8%) |
|  | Missing | 40 (0%) |
| Age 12 religious service attendance | At least 1/week | 1943 (20%) |
|  | 1-3/month | 1899 (20%) |
|  | <1/ month | 2887 (30%) |
|  | Never | 2749 (29%) |
|  | Missing | 27 (0%) |
| Year of birth | 1998-2005; current age: 18-24 | 829 (9%) |
|  | 1993-1998; age 25-29 | 774 (8%) |
|  | 1983-1993; age 30-39 | 1438 (15%) |
|  | 1973-1983; age 40-49 | 1494 (16%) |
|  | 1963-1973; age 50-59 | 1729 (18%) |
|  | 1953-1963; age 60-69 | 1915 (20%) |
|  | 1943-1953; age 70-79 | 1137 (12%) |
|  | 1943 or earlier; age 80+ | 190 (2%) |
| Gender | Male | 4641 (49%) |
|  | Female | 4843 (51%) |
|  | Other | 11 (0%) |
|  | Missing | 11 (0%) |
| Religious affiliation at age 12 | Christianity | 5751 (61%) |
|  | Islam | 350 (4%) |
|  | Hinduism | 15 (0%) |
|  | Buddhism | 25 (0%) |
|  | Judaism | 18 (0%) |
|  | Sikhism | 5 (0%) |
|  | Baha’i | 2 (0%) |
|  | Jainism | 1 (0%) |
|  | Confucianism | 4 (0%) |
|  | Primal, Animist, or Folk religion | 19 (0%) |
|  | Some other religion | 67 (1%) |
|  | No religion/Atheist/Agnostic | 3163 (33%) |
|  | Missing | 85 (1%) |

***Supplementary Table S5B. Regression of Promoting Good on Childhood Predictors for Germany***

| **Variable** | **Category** | **Estimate** | **SE** | **95% CI** | **Global P-value** |
| --- | --- | --- | --- | --- | --- |
| Relationship with mother (ref: Very bad/Somewhat bad) | Very good/Somewhat good | 0.13 | 0.09 | (-0.06,0.31) | 0.173 |
| Relationship with father (ref: Very bad/Somewhat bad) | Very good/Somewhat good | 0.04 | 0.08 | (-0.12,0.19) | 0.611 |
| Parent marital status (ref: Married) | Divorced | 0.14 | 0.08 | (-0.02,0.30) | 0.033 |
|  | Single, never married | -0.15 | 0.11 | (-0.36,0.07) |  |
|  | One or both parents had died | -0.26 | 0.15 | (-0.56,0.03) |  |
| Subjective financial status of family growing up (ref: Got by) | Lived comfortably | -0.06 | 0.06 | (-0.18,0.05) | 0.665 |
|  | Found it difficult | -0.05 | 0.08 | (-0.19,0.10) |  |
|  | Found it very difficult | 0.02 | 0.15 | (-0.28,0.31) |  |
| Abuse (ref: No) | Yes | 0.06 | 0.08 | (-0.09,0.21) | 0.435 |
| Felt like an outsider in the family (ref: No) | Yes | -0.12 | 0.08 | (-0.27,0.03) | 0.115 |
| Self-rated health growing up (ref: Good) | Excellent | 0.45 | 0.07 | (0.31,0.58) | <.001 |
|  | Very good | 0.16 | 0.06 | (0.04,0.28) |  |
|  | Fair | 0.14 | 0.10 | (-0.06,0.33) |  |
|  | Poor | 0.36 | 0.23 | (-0.08,0.81) |  |
| Immigration status (ref: Born in this country) | Born in another country | 0.19 | 0.09 | (0.01,0.37) | 0.034 |
| Age 12 religious service attendance (ref: Never) | At least 1/week | 0.46 | 0.07 | (0.31,0.60) | <.001 |
|  | 1-3/month | 0.08 | 0.07 | (-0.06,0.23) |  |
|  | <1/month | 0.13 | 0.07 | (0.00,0.26) |  |
| Birth year (ref: 1998-2005; current age: 18-24) | 1993-1998; age 25-29 | 0.42 | 0.12 | (0.19,0.65) | <.001 |
|  | 1983-1993; age 30-39 | 0.40 | 0.11 | (0.19,0.61) |  |
|  | 1973-1983; age 40-49 | 0.55 | 0.11 | (0.34,0.76) |  |
|  | 1963-1973; age 50-59 | 0.43 | 0.11 | (0.22,0.64) |  |
|  | 1953-1963; age 60-69 | 0.72 | 0.11 | (0.51,0.92) |  |
|  | 1943-1953; age 70-79 | 0.81 | 0.11 | (0.59,1.03) |  |
|  | 1943 or earlier; age 80+ | 0.70 | 0.21 | (0.29,1.11) |  |
| Gender (ref:Male) | Female | 0.10 | 0.05 | (0.01,0.20) | 0.077 |
|  | Other | 0.26 | 0.47 | (-0.65,1.18) |  |
| Religious affiliation at age 12 (ref: No religion/Atheist/Agnostic) | Christianity | 0.05 | 0.06 | (-0.06,0.16) | 0.242 |
|  | Islam | 0.27 | 0.14 | (0.00,0.53) |  |
|  | Collapsed affiliations with prevalence<3% | -0.04 | 0.25 | (-0.53,0.45) |  |

***Supplementary Table S5C. E-Values for Estimates and CI for Germany***

| **Variable** | **Category** | **E-Value for Estimate** | **E-Value for 95% CI** |
| --- | --- | --- | --- |
| Relationship with mother (ref: Very bad/Somewhat bad) | Very good/Somewhat good | 1.33 | 1.00 |
| Relationship with father (ref: Very bad/Somewhat bad) | Very good/Somewhat good | 1.16 | 1.00 |
| Parent marital status (ref: Married) | Divorced | 1.35 | 1.00 |
|  | Single, never married | 1.36 | 1.00 |
|  | One or both parents had died | 1.54 | 1.00 |
| Subjective financial status of family growing up (ref: Got by) | Lived comfortably | 1.22 | 1.00 |
|  | Found it difficult | 1.18 | 1.00 |
|  | Found it very difficult | 1.11 | 1.00 |
| Abuse (ref: No) | Yes | 1.21 | 1.00 |
| Felt like an outsider in the family (ref: No) | Yes | 1.31 | 1.00 |
| Self-rated health growing up (ref: Good) | Excellent | 1.80 | 1.61 |
|  | Very good | 1.38 | 1.17 |
|  | Fair | 1.34 | 1.00 |
|  | Poor | 1.68 | 1.00 |
| Immigration status (ref: Born in this country) | Born in another country | 1.43 | 1.09 |
| Age 12 religious service attendance (ref: Never) | At least 1/week | 1.82 | 1.61 |
|  | 1-3/month | 1.25 | 1.00 |
|  | <1/month | 1.33 | 1.04 |
| Birth year (ref: 1998-2005; current age: 18-24) | 1993-1998; age 25-29 | 1.77 | 1.43 |
|  | 1983-1993; age 30-39 | 1.74 | 1.44 |
|  | 1973-1983; age 40-49 | 1.96 | 1.66 |
|  | 1963-1973; age 50-59 | 1.78 | 1.47 |
|  | 1953-1963; age 60-69 | 2.21 | 1.90 |
|  | 1943-1953; age 70-79 | 2.35 | 2.01 |
|  | 1943 or earlier; age 80+ | 2.18 | 1.57 |
| Gender (ref:Male) | Female | 1.29 | 1.09 |
|  | Other | 1.54 | 1.00 |
| Religious affiliation at age 12 (ref: No religion/Atheist/Agnostic) | Christianity | 1.18 | 1.00 |
|  | Islam | 1.54 | 1.02 |
|  | Collapsed affiliations with prevalence<3% | 1.16 | 1.00 |

***Supplementary Table S6A. Nationally Representative Childhood Descriptive Statistics for Hong Kong***

| **Variable** | **Category** | **N (%)** |
| --- | --- | --- |
| Relationship with mother | Very good | 1077 (36%) |
|  | Somewhat good | 1164 (39%) |
|  | Somewhat bad | 293 (10%) |
|  | Very bad | 49 (2%) |
|  | Does not apply | 426 (14%) |
|  | Missing | 3 (0%) |
| Relationships with father | Very good | 868 (29%) |
|  | Somewhat good | 1089 (36%) |
|  | Somewhat bad | 393 (13%) |
|  | Very bad | 102 (3%) |
|  | Does not apply | 557 (19%) |
|  | Missing | 3 (0%) |
| Parent marital status | Parents married | 2752 (91%) |
|  | Divorced | 114 (4%) |
|  | Single, never married | 40 (1%) |
|  | One or both parents had died | 50 (2%) |
|  | Missing | 56 (2%) |
| Subjective financial status of family growing up | Lived comfortably | 906 (30%) |
|  | Got by | 1527 (51%) |
|  | Found it difficult | 473 (16%) |
|  | Found it very difficult | 84 (3%) |
|  | Missing | 22 (1%) |
| Abuse | Yes | 318 (11%) |
|  | No | 2688 (89%) |
|  | Missing | 5 (0%) |
| Felt like an outsider in the family | Yes | 664 (22%) |
|  | No | 2224 (74%) |
|  | Missing | 124 (4%) |
| Self-rated health growing up | Excellent | 545 (18%) |
|  | Very good | 1073 (36%) |
|  | Good | 863 (29%) |
|  | Fair | 426 (14%) |
|  | Poor | 91 (3%) |
|  | Missing | 13 (0%) |
| Immigration status | Born in this country | 2637 (88%) |
|  | Born in another country | 321 (11%) |
|  | Missing | 53 (2%) |
| Age 12 religious service attendance | At least 1/week | 432 (14%) |
|  | 1-3/month | 528 (18%) |
|  | <1/ month | 753 (25%) |
|  | Never | 1295 (43%) |
|  | Missing | 4 (0%) |
| Year of birth | 1998-2005; current age: 18-24 | 217 (7%) |
|  | 1993-1998; age 25-29 | 198 (7%) |
|  | 1983-1993; age 30-39 | 507 (17%) |
|  | 1973-1983; age 40-49 | 580 (19%) |
|  | 1963-1973; age 50-59 | 711 (24%) |
|  | 1953-1963; age 60-69 | 620 (21%) |
|  | 1943-1953; age 70-79 | 164 (5%) |
|  | 1943 or earlier; age 80+ | 15 (0%) |
| Gender | Male | 1390 (46%) |
|  | Female | 1620 (54%) |
|  | Other | 2 (0%) |
| Religious affiliation at age 12 | Christianity | 715 (24%) |
|  | Islam | 86 (3%) |
|  | Hinduism | 27 (1%) |
|  | Buddhism | 323 (11%) |
|  | Judaism | 16 (1%) |
|  | Sikhism | 4 (0%) |
|  | Jainism | 1 (0%) |
|  | Shinto | 18 (1%) |
|  | Taoism | 81 (3%) |
|  | Confucianism | 10 (0%) |
|  | Primal, Animist, or Folk religion | 15 (0%) |
|  | Chinese folk/traditional religion | 108 (4%) |
|  | Some other religion | 5 (0%) |
|  | No religion/Atheist/Agnostic | 1601 (53%) |
|  | Missing | 1 (0%) |
| Race/ethnicity | Chinese (Cantonese) | 1930 (64%) |
|  | Chinese (Chaoshan) | 201 (7%) |
|  | Chinese (Fujianese) | 117 (4%) |
|  | Chinese (Hakka) | 121 (4%) |
|  | Chinese (Shanghainese) | 89 (3%) |
|  | Chinese (Other ethnicity) | 264 (9%) |
|  | East Asian (Korean, Japanese) | 10 (0%) |
|  | Southeast Asian (Filipino, Indonesian, Thailand) | 46 (2%) |
|  | South Asian (Indian, Nepalese, Pakistani) | 17 (1%) |
|  | Taiwanese | 14 (0%) |
|  | White | 15 (0%) |
|  | Other | 4 (0%) |
|  | Missing | 184 (6%) |

***Supplementary Table S6B. Regression of Promoting Good on Childhood Predictors for Hong Kong***

| **Variable** | **Category** | **Estimate** | **SE** | **95% CI** | **Global P-value** |
| --- | --- | --- | --- | --- | --- |
| Relationship with mother (ref: Very bad/Somewhat bad) | Very good/Somewhat good | 0.04 | 0.12 | (-0.20,0.28) | 0.666 |
| Relationship with father (ref: Very bad/Somewhat bad) | Very good/Somewhat good | 0.21 | 0.13 | (-0.04,0.46) | 0.088 |
| Parent marital status (ref: Married) | Divorced | 0.21 | 0.27 | (-0.31,0.74) | 0.264 |
|  | Single, never married | 0.59 | 0.34 | (-0.06,1.25) |  |
|  | One or both parents had died | 0.16 | 0.31 | (-0.45,0.77) |  |
| Subjective financial status of family growing up (ref: Got by) | Lived comfortably | 0.73 | 0.10 | (0.54,0.93) | <.001 |
|  | Found it difficult | -0.18 | 0.14 | (-0.45,0.09) |  |
|  | Found it very difficult | 0.05 | 0.34 | (-0.62,0.71) |  |
| Abuse (ref: No) | Yes | 0.13 | 0.14 | (-0.15,0.41) | 0.377 |
| Felt like an outsider in the family (ref: No) | Yes | -0.02 | 0.12 | (-0.25,0.20) | 0.796 |
| Self-rated health growing up (ref: Good) | Excellent | 1.67 | 0.15 | (1.37,1.97) | <.001 |
|  | Very good | 0.80 | 0.10 | (0.60,0.99) |  |
|  | Fair | -0.69 | 0.13 | (-0.95,-0.43) |  |
|  | Poor | -1.12 | 0.54 | (-2.18,-0.07) |  |
| Immigration status (ref: Born in this country) | Born in another country | -0.24 | 0.18 | (-0.59,0.12) | 0.190 |
| Age 12 religious service attendance (ref: Never) | At least 1/week | 0.50 | 0.16 | (0.18,0.82) | <.001 |
|  | 1-3/month | 0.51 | 0.13 | (0.25,0.77) |  |
|  | <1/month | 0.12 | 0.11 | (-0.10,0.34) |  |
| Birth year (ref: 1998-2005; current age: 18-24) | 1993-1998; age 25-29 | -0.14 | 0.17 | (-0.48,0.20) | <.001 |
|  | 1983-1993; age 30-39 | 0.02 | 0.14 | (-0.25,0.29) |  |
|  | 1973-1983; age 40-49 | 0.37 | 0.12 | (0.12,0.61) |  |
|  | 1963-1973; age 50-59 | 0.61 | 0.12 | (0.37,0.85) |  |
|  | 1953-1963; age 60-69 | 0.94 | 0.15 | (0.63,1.24) |  |
|  | 1943-1953; age 70-79 | 1.05 | 0.32 | (0.42,1.68) |  |
|  | 1943 or earlier; age 80+ | -0.32 | 0.25 | (-0.81,0.18) |  |
| Gender (ref:Male) | Female | 0.10 | 0.07 | (-0.04,0.24) | 0.047 |
|  | Other | 1.00 | 0.47 | (0.08,1.92) |  |
| Religious affiliation at age 12 (ref: No religion/Atheist/Agnostic) | Christianity | -0.13 | 0.15 | (-0.41,0.16) | 0.272 |
|  | Chinese folk/traditional religion | 0.09 | 0.22 | (-0.35,0.53) |  |
|  | Buddhism | -0.02 | 0.13 | (-0.29,0.24) |  |
|  | Collapsed affiliations with prevalence<3% | 0.25 | 0.18 | (-0.10,0.61) |  |
| Race/ethnicity (ref: plurality) | Race/ethnicity minority | 0.15 | 0.10 | (-0.03,0.34) | 0.073 |

***Supplementary Table S6C. E-Values for Estimates and CI for Hong Kong***

| **Variable** | **Category** | **E-Value for Estimate** | **E-Value for 95% CI** |
| --- | --- | --- | --- |
| Relationship with mother (ref: Very bad/Somewhat bad) | Very good/Somewhat good | 1.17 | 1.00 |
| Relationship with father (ref: Very bad/Somewhat bad) | Very good/Somewhat good | 1.44 | 1.00 |
| Parent marital status (ref: Married) | Divorced | 1.45 | 1.00 |
|  | Single, never married | 1.98 | 1.00 |
|  | One or both parents had died | 1.37 | 1.00 |
| Subjective financial status of family growing up (ref: Got by) | Lived comfortably | 2.18 | 1.91 |
|  | Found it difficult | 1.40 | 1.00 |
|  | Found it very difficult | 1.17 | 1.00 |
| Abuse (ref: No) | Yes | 1.32 | 1.00 |
| Felt like an outsider in the family (ref: No) | Yes | 1.12 | 1.00 |
| Self-rated health growing up (ref: Good) | Excellent | 3.82 | 3.22 |
|  | Very good | 2.27 | 1.99 |
|  | Fair | 2.11 | 1.74 |
|  | Poor | 2.78 | 1.22 |
| Immigration status (ref: Born in this country) | Born in another country | 1.48 | 1.00 |
| Age 12 religious service attendance (ref: Never) | At least 1/week | 1.84 | 1.39 |
|  | 1-3/month | 1.86 | 1.51 |
|  | <1/month | 1.31 | 1.00 |
| Birth year (ref: 1998-2005; current age: 18-24) | 1993-1998; age 25-29 | 1.34 | 1.00 |
|  | 1983-1993; age 30-39 | 1.11 | 1.00 |
|  | 1973-1983; age 40-49 | 1.66 | 1.31 |
|  | 1963-1973; age 50-59 | 2.00 | 1.66 |
|  | 1953-1963; age 60-69 | 2.48 | 2.03 |
|  | 1943-1953; age 70-79 | 2.66 | 1.73 |
|  | 1943 or earlier; age 80+ | 1.59 | 1.00 |
| Gender (ref:Male) | Female | 1.27 | 1.00 |
|  | Other | 2.58 | 1.25 |
| Religious affiliation at age 12 (ref: No religion/Atheist/Agnostic) | Christianity | 1.32 | 1.00 |
|  | Chinese folk/traditional religion | 1.25 | 1.00 |
|  | Buddhism | 1.11 | 1.00 |
|  | Collapsed affiliations with prevalence<3% | 1.50 | 1.00 |
| Race/ethnicity (ref: plurality) | Race/ethnicity minority | 1.36 | 1.00 |

***Supplementary Table S7A. Nationally Representative Childhood Descriptive Statistics for India***

| **Variable** | **Category** | **N (%)** |
| --- | --- | --- |
| Relationship with mother | Very good | 11465 (90%) |
|  | Somewhat good | 788 (6%) |
|  | Somewhat bad | 88 (1%) |
|  | Very bad | 73 (1%) |
|  | Does not apply | 269 (2%) |
|  | Missing | 82 (1%) |
| Relationships with father | Very good | 10923 (86%) |
|  | Somewhat good | 995 (8%) |
|  | Somewhat bad | 126 (1%) |
|  | Very bad | 100 (1%) |
|  | Does not apply | 481 (4%) |
|  | Missing | 141 (1%) |
| Parent marital status | Parents married | 5578 (44%) |
|  | Divorced | 236 (2%) |
|  | Single, never married | 1055 (8%) |
|  | One or both parents had died | 940 (7%) |
|  | Missing | 4956 (39%) |
| Subjective financial status of family growing up | Lived comfortably | 4946 (39%) |
|  | Got by | 3010 (24%) |
|  | Found it difficult | 2703 (21%) |
|  | Found it very difficult | 2035 (16%) |
|  | Missing | 70 (1%) |
| Abuse | Yes | 1468 (11%) |
|  | No | 10526 (82%) |
|  | Missing | 771 (6%) |
| Felt like an outsider in the family | Yes | 1926 (15%) |
|  | No | 10780 (84%) |
|  | Missing | 59 (0%) |
| Self-rated health growing up | Excellent | 2182 (17%) |
|  | Very good | 3882 (30%) |
|  | Good | 4028 (32%) |
|  | Fair | 2202 (17%) |
|  | Poor | 424 (3%) |
|  | Missing | 47 (0%) |
| Immigration status | Born in this country | 12629 (99%) |
|  | Born in another country | 110 (1%) |
|  | Missing | 26 (0%) |
| Age 12 religious service attendance | At least 1/week | 5288 (41%) |
|  | 1-3/month | 2959 (23%) |
|  | <1/ month | 2719 (21%) |
|  | Never | 1478 (12%) |
|  | Missing | 321 (3%) |
| Year of birth | 1998-2005; current age: 18-24 | 2543 (20%) |
|  | 1993-1998; age 25-29 | 1640 (13%) |
|  | 1983-1993; age 30-39 | 3109 (24%) |
|  | 1973-1983; age 40-49 | 2275 (18%) |
|  | 1963-1973; age 50-59 | 1574 (12%) |
|  | 1953-1963; age 60-69 | 1188 (9%) |
|  | 1943-1953; age 70-79 | 370 (3%) |
|  | 1943 or earlier; age 80+ | 67 (1%) |
| Gender | Male | 6473 (51%) |
|  | Female | 6292 (49%) |
| Religious affiliation at age 12 | Christianity | 254 (2%) |
|  | Islam | 1550 (12%) |
|  | Hinduism | 10417 (82%) |
|  | Buddhism | 180 (1%) |
|  | Sikhism | 126 (1%) |
|  | Jainism | 9 (0%) |
|  | Shinto | 4 (0%) |
|  | Primal, Animist, or Folk religion | 27 (0%) |
|  | Some other religion | 59 (0%) |
|  | No religion/Atheist/Agnostic | 7 (0%) |
|  | Missing | 131 (1%) |
| Race/ethnicity | General | 3538 (28%) |
|  | Other backward caste | 4177 (33%) |
|  | Schedule caste | 3599 (28%) |
|  | Schedule tribe | 1185 (9%) |
|  | Missing | 267 (2%) |

***Supplementary Table S7B. Regression of Promoting Good on Childhood Predictors for India***

| **Variable** | **Category** | **Estimate** | **SE** | **95% CI** | **Global P-value** |
| --- | --- | --- | --- | --- | --- |
| Relationship with mother (ref: Very bad/Somewhat bad) | Very good/Somewhat good | 0.20 | 0.23 | (-0.25,0.65) | 0.374 |
| Relationship with father (ref: Very bad/Somewhat bad) | Very good/Somewhat good | -0.22 | 0.19 | (-0.60,0.16) | 0.250 |
| Parent marital status (ref: Married) | Divorced | -0.14 | 0.23 | (-0.62,0.35) | <.001 |
|  | Single, never married | 0.08 | 0.10 | (-0.11,0.27) |  |
|  | One or both parents had died | 0.48 | 0.12 | (0.24,0.73) |  |
| Subjective financial status of family growing up (ref: Got by) | Lived comfortably | -0.16 | 0.07 | (-0.30,-0.01) | <.001 |
|  | Found it difficult | -0.17 | 0.09 | (-0.34,0.00) |  |
|  | Found it very difficult | -0.65 | 0.10 | (-0.85,-0.45) |  |
| Abuse (ref: No) | Yes | -0.14 | 0.10 | (-0.33,0.05) | 0.134 |
| Felt like an outsider in the family (ref: No) | Yes | -0.02 | 0.09 | (-0.20,0.16) | 0.799 |
| Self-rated health growing up (ref: Good) | Excellent | 0.06 | 0.09 | (-0.11,0.24) | 0.014 |
|  | Very good | 0.09 | 0.08 | (-0.06,0.24) |  |
|  | Fair | -0.23 | 0.09 | (-0.41,-0.05) |  |
|  | Poor | -0.05 | 0.17 | (-0.38,0.28) |  |
| Immigration status (ref: Born in this country) | Born in another country | -0.66 | 0.26 | (-1.16,-0.16) | 0.010 |
| Age 12 religious service attendance (ref: Never) | At least 1/week | 0.17 | 0.10 | (-0.03,0.37) | 0.135 |
|  | 1-3/month | 0.22 | 0.11 | (0.01,0.43) |  |
|  | <1/month | 0.10 | 0.12 | (-0.13,0.32) |  |
| Birth year (ref: 1998-2005; current age: 18-24) | 1993-1998; age 25-29 | -0.10 | 0.10 | (-0.29,0.09) | <.001 |
|  | 1983-1993; age 30-39 | -0.23 | 0.09 | (-0.40,-0.06) |  |
|  | 1973-1983; age 40-49 | -0.37 | 0.10 | (-0.56,-0.18) |  |
|  | 1963-1973; age 50-59 | -0.38 | 0.11 | (-0.60,-0.16) |  |
|  | 1953-1963; age 60-69 | -0.40 | 0.12 | (-0.64,-0.15) |  |
|  | 1943-1953; age 70-79 | -0.51 | 0.19 | (-0.88,-0.15) |  |
|  | 1943 or earlier; age 80+ | -1.39 | 0.59 | (-2.55,-0.22) |  |
| Gender (ref:Male) | Female | 0.07 | 0.06 | (-0.05,0.18) | 0.250 |
| Religious affiliation at age 12 (ref: Hinduism) | Islam | -0.06 | 0.12 | (-0.30,0.18) | 0.022 |
|  | Collapsed affiliations with prevalence<3% | 0.32 | 0.12 | (0.08,0.57) |  |
| Race/ethnicity (ref: plurality) | Race/ethnicity minority | -0.14 | 0.07 | (-0.27,-0.00) | 0.044 |

***Supplementary Table S7C. E-Values for Estimates and CI for India***

| **Variable** | **Category** | **E-Value for Estimate** | **E-Value for 95% CI** |
| --- | --- | --- | --- |
| Relationship with mother (ref: Very bad/Somewhat bad) | Very good/Somewhat good | 1.35 | 1.00 |
| Relationship with father (ref: Very bad/Somewhat bad) | Very good/Somewhat good | 1.37 | 1.00 |
| Parent marital status (ref: Married) | Divorced | 1.27 | 1.00 |
|  | Single, never married | 1.20 | 1.00 |
|  | One or both parents had died | 1.64 | 1.40 |
| Subjective financial status of family growing up (ref: Got by) | Lived comfortably | 1.30 | 1.08 |
|  | Found it difficult | 1.31 | 1.00 |
|  | Found it very difficult | 1.81 | 1.61 |
| Abuse (ref: No) | Yes | 1.28 | 1.00 |
| Felt like an outsider in the family (ref: No) | Yes | 1.09 | 1.00 |
| Self-rated health growing up (ref: Good) | Excellent | 1.17 | 1.00 |
|  | Very good | 1.21 | 1.00 |
|  | Fair | 1.38 | 1.16 |
|  | Poor | 1.15 | 1.00 |
| Immigration status (ref: Born in this country) | Born in another country | 1.81 | 1.30 |
| Age 12 religious service attendance (ref: Never) | At least 1/week | 1.31 | 1.00 |
|  | 1-3/month | 1.37 | 1.08 |
|  | <1/month | 1.22 | 1.00 |
| Birth year (ref: 1998-2005; current age: 18-24) | 1993-1998; age 25-29 | 1.23 | 1.00 |
|  | 1983-1993; age 30-39 | 1.38 | 1.16 |
|  | 1973-1983; age 40-49 | 1.52 | 1.32 |
|  | 1963-1973; age 50-59 | 1.53 | 1.30 |
|  | 1953-1963; age 60-69 | 1.55 | 1.29 |
|  | 1943-1953; age 70-79 | 1.67 | 1.28 |
|  | 1943 or earlier; age 80+ | 2.58 | 1.37 |
| Gender (ref:Male) | Female | 1.18 | 1.00 |
| Religious affiliation at age 12 (ref: Hinduism) | Islam | 1.16 | 1.00 |
|  | Collapsed affiliations with prevalence<3% | 1.47 | 1.19 |
| Race/ethnicity (ref: plurality) | Race/ethnicity minority | 1.27 | 1.02 |

***Supplementary Table S8A. Nationally Representative Childhood Descriptive Statistics for Indonesia***

| **Variable** | **Category** | **N (%)** |
| --- | --- | --- |
| Relationship with mother | Very good | 6238 (89%) |
|  | Somewhat good | 583 (8%) |
|  | Somewhat bad | 50 (1%) |
|  | Very bad | 26 (0%) |
|  | Does not apply | 68 (1%) |
|  | Missing | 27 (0%) |
| Relationships with father | Very good | 6067 (87%) |
|  | Somewhat good | 628 (9%) |
|  | Somewhat bad | 68 (1%) |
|  | Very bad | 52 (1%) |
|  | Does not apply | 115 (2%) |
|  | Missing | 61 (1%) |
| Parent marital status | Parents married | 5557 (79%) |
|  | Divorced | 448 (6%) |
|  | Single, never married | 47 (1%) |
|  | One or both parents had died | 735 (11%) |
|  | Missing | 205 (3%) |
| Subjective financial status of family growing up | Lived comfortably | 3408 (49%) |
|  | Got by | 2955 (42%) |
|  | Found it difficult | 439 (6%) |
|  | Found it very difficult | 181 (3%) |
|  | Missing | 9 (0%) |
| Abuse | Yes | 486 (7%) |
|  | No | 6427 (92%) |
|  | Missing | 79 (1%) |
| Felt like an outsider in the family | Yes | 343 (5%) |
|  | No | 6639 (95%) |
|  | Missing | 10 (0%) |
| Self-rated health growing up | Excellent | 1246 (18%) |
|  | Very good | 1968 (28%) |
|  | Good | 2490 (36%) |
|  | Fair | 1233 (18%) |
|  | Poor | 55 (1%) |
|  | Missing | 1 (0%) |
| Immigration status | Born in this country | 6958 (100%) |
|  | Born in another country | 34 (0%) |
| Age 12 religious service attendance | At least 1/week | 5363 (77%) |
|  | 1-3/month | 973 (14%) |
|  | <1/ month | 329 (5%) |
|  | Never | 275 (4%) |
|  | Missing | 51 (1%) |
| Year of birth | 1998-2005; current age: 18-24 | 1216 (17%) |
|  | 1993-1998; age 25-29 | 849 (12%) |
|  | 1983-1993; age 30-39 | 1591 (23%) |
|  | 1973-1983; age 40-49 | 1576 (23%) |
|  | 1963-1973; age 50-59 | 1169 (17%) |
|  | 1953-1963; age 60-69 | 490 (7%) |
|  | 1943-1953; age 70-79 | 83 (1%) |
|  | 1943 or earlier; age 80+ | 17 (0%) |
| Gender | Male | 3461 (50%) |
|  | Female | 3513 (50%) |
|  | Other | 7 (0%) |
|  | Missing | 11 (0%) |
| Religious affiliation at age 12 | Christianity | 528 (8%) |
|  | Islam | 6373 (91%) |
|  | Hinduism | 75 (1%) |
|  | Buddhism | 5 (0%) |
|  | Jainism | 1 (0%) |
|  | Taoism | 0 (0%) |
|  | Confucianism | 1 (0%) |
|  | Primal, Animist, or Folk religion | 1 (0%) |
|  | No religion/Atheist/Agnostic | 2 (0%) |
|  | Missing | 8 (0%) |
| Race/ethnicity | Banjar/Melayu Banjar | 320 (5%) |
|  | Betawi | 251 (4%) |
|  | Bugis | 243 (3%) |
|  | Jawa | 2846 (41%) |
|  | Madura | 262 (4%) |
|  | Minangkabau | 273 (4%) |
|  | Sunda/Parahyangan | 1172 (17%) |
|  | Bali | 69 (1%) |
|  | Batak | 165 (2%) |
|  | Makasar | 91 (1%) |
|  | Other | 1262 (18%) |
|  | Missing | 38 (1%) |

***Supplementary Table S8B. Regression of Promoting Good on Childhood Predictors for Indonesia***

| **Variable** | **Category** | **Estimate** | **SE** | **95% CI** | **Global P-value** |
| --- | --- | --- | --- | --- | --- |
| Relationship with mother (ref: Very bad/Somewhat bad) | Very good/Somewhat good | 0.67 | 0.39 | (-0.10,1.43) | 0.085 |
| Relationship with father (ref: Very bad/Somewhat bad) | Very good/Somewhat good | -0.01 | 0.20 | (-0.41,0.38) | 0.883 |
| Parent marital status (ref: Married) | Divorced | -0.00 | 0.13 | (-0.26,0.26) | 0.506 |
|  | Single, never married | -0.42 | 0.42 | (-1.24,0.41) |  |
|  | One or both parents had died | 0.12 | 0.12 | (-0.11,0.35) |  |
| Subjective financial status of family growing up (ref: Got by) | Lived comfortably | 0.20 | 0.06 | (0.08,0.32) | 0.001 |
|  | Found it difficult | -0.13 | 0.15 | (-0.42,0.15) |  |
|  | Found it very difficult | -0.25 | 0.28 | (-0.80,0.30) |  |
| Abuse (ref: No) | Yes | -0.39 | 0.17 | (-0.72,-0.07) | 0.016 |
| Felt like an outsider in the family (ref: No) | Yes | -0.11 | 0.17 | (-0.46,0.23) | 0.508 |
| Self-rated health growing up (ref: Good) | Excellent | 0.27 | 0.09 | (0.09,0.44) | 0.008 |
|  | Very good | 0.21 | 0.07 | (0.08,0.34) |  |
|  | Fair | 0.09 | 0.08 | (-0.07,0.24) |  |
|  | Poor | 0.36 | 0.44 | (-0.50,1.23) |  |
| Immigration status (ref: Born in this country) | Born in another country | -0.17 | 0.73 | (-1.61,1.26) | 0.813 |
| Age 12 religious service attendance (ref: Never) | At least 1/week | 0.19 | 0.15 | (-0.10,0.49) | 0.187 |
|  | 1-3/month | 0.08 | 0.17 | (-0.25,0.40) |  |
|  | <1/month | 0.01 | 0.18 | (-0.33,0.36) |  |
| Birth year (ref: 1998-2005; current age: 18-24) | 1993-1998; age 25-29 | 0.19 | 0.09 | (0.01,0.38) | 0.007 |
|  | 1983-1993; age 30-39 | 0.20 | 0.09 | (0.02,0.37) |  |
|  | 1973-1983; age 40-49 | 0.28 | 0.09 | (0.11,0.45) |  |
|  | 1963-1973; age 50-59 | 0.08 | 0.12 | (-0.15,0.31) |  |
|  | 1953-1963; age 60-69 | -0.16 | 0.15 | (-0.46,0.14) |  |
|  | 1943-1953; age 70-79 | 0.08 | 0.26 | (-0.44,0.59) |  |
|  | 1943 or earlier; age 80+ | -0.39 | 0.79 | (-1.94,1.16) |  |
| Gender (ref:Male) | Female | 0.11 | 0.06 | (0.00,0.22) | 0.006 |
|  | Other | -2.19 | 0.90 | (-3.96,-0.41) |  |
| Religious affiliation at age 12 (ref: Islam) | Christianity | -0.05 | 0.10 | (-0.25,0.16) | 0.637 |
|  | Collapsed affiliations with prevalence<3% | 0.20 | 0.24 | (-0.27,0.67) |  |
| Race/ethnicity (ref: plurality) | Race/ethnicity minority | 0.03 | 0.06 | (-0.09,0.15) | 0.592 |

***Supplementary Table S8C. E-Values for Estimates and CI for Indonesia***

| **Variable** | **Category** | **E-Value for Estimate** | **E-Value for 95% CI** |
| --- | --- | --- | --- |
| Relationship with mother (ref: Very bad/Somewhat bad) | Very good/Somewhat good | 2.15 | 1.00 |
| Relationship with father (ref: Very bad/Somewhat bad) | Very good/Somewhat good | 1.09 | 1.00 |
| Parent marital status (ref: Married) | Divorced | 1.03 | 1.00 |
|  | Single, never married | 1.77 | 1.00 |
|  | One or both parents had died | 1.33 | 1.00 |
| Subjective financial status of family growing up (ref: Got by) | Lived comfortably | 1.44 | 1.24 |
|  | Found it difficult | 1.34 | 1.00 |
|  | Found it very difficult | 1.53 | 1.00 |
| Abuse (ref: No) | Yes | 1.74 | 1.23 |
| Felt like an outsider in the family (ref: No) | Yes | 1.31 | 1.00 |
| Self-rated health growing up (ref: Good) | Excellent | 1.55 | 1.28 |
|  | Very good | 1.46 | 1.24 |
|  | Fair | 1.26 | 1.00 |
|  | Poor | 1.70 | 1.00 |
| Immigration status (ref: Born in this country) | Born in another country | 1.41 | 1.00 |
| Age 12 religious service attendance (ref: Never) | At least 1/week | 1.44 | 1.00 |
|  | 1-3/month | 1.24 | 1.00 |
|  | <1/month | 1.09 | 1.00 |
| Birth year (ref: 1998-2005; current age: 18-24) | 1993-1998; age 25-29 | 1.44 | 1.08 |
|  | 1983-1993; age 30-39 | 1.45 | 1.13 |
|  | 1973-1983; age 40-49 | 1.57 | 1.31 |
|  | 1963-1973; age 50-59 | 1.25 | 1.00 |
|  | 1953-1963; age 60-69 | 1.39 | 1.00 |
|  | 1943-1953; age 70-79 | 1.25 | 1.00 |
|  | 1943 or earlier; age 80+ | 1.73 | 1.00 |
| Gender (ref:Male) | Female | 1.31 | 1.03 |
|  | Other | 5.49 | 1.78 |
| Religious affiliation at age 12 (ref: Islam) | Christianity | 1.18 | 1.00 |
|  | Collapsed affiliations with prevalence<3% | 1.45 | 1.00 |
| Race/ethnicity (ref: plurality) | Race/ethnicity minority | 1.14 | 1.00 |

***Supplementary Table S9A. Nationally Representative Childhood Descriptive Statistics for Israel***

| **Variable** | **Category** | **N (%)** |
| --- | --- | --- |
| Relationship with mother | Very good | 2686 (73%) |
|  | Somewhat good | 793 (22%) |
|  | Somewhat bad | 110 (3%) |
|  | Very bad | 18 (0%) |
|  | Does not apply | 45 (1%) |
|  | Missing | 17 (0%) |
| Relationships with father | Very good | 2290 (62%) |
|  | Somewhat good | 912 (25%) |
|  | Somewhat bad | 234 (6%) |
|  | Very bad | 37 (1%) |
|  | Does not apply | 171 (5%) |
|  | Missing | 25 (1%) |
| Parent marital status | Parents married | 3172 (86%) |
|  | Divorced | 284 (8%) |
|  | Single, never married | 36 (1%) |
|  | One or both parents had died | 130 (4%) |
|  | Missing | 47 (1%) |
| Subjective financial status of family growing up | Lived comfortably | 923 (25%) |
|  | Got by | 1822 (50%) |
|  | Found it difficult | 667 (18%) |
|  | Found it very difficult | 239 (7%) |
|  | Missing | 17 (0%) |
| Felt like an outsider in the family | Yes | 371 (10%) |
|  | No | 3228 (88%) |
|  | Missing | 70 (2%) |
| Self-rated health growing up | Excellent | 1785 (49%) |
|  | Very good | 1284 (35%) |
|  | Good | 480 (13%) |
|  | Fair | 105 (3%) |
|  | Poor | 6 (0%) |
|  | Missing | 8 (0%) |
| Immigration status | Born in this country | 2796 (76%) |
|  | Born in another country | 868 (24%) |
|  | Missing | 5 (0%) |
| Age 12 religious service attendance | At least 1/week | 867 (24%) |
|  | 1-3/month | 435 (12%) |
|  | <1/ month | 810 (22%) |
|  | Never | 1539 (42%) |
|  | Missing | 17 (0%) |
| Year of birth | 1998-2005; current age: 18-24 | 553 (15%) |
|  | 1993-1998; age 25-29 | 407 (11%) |
|  | 1983-1993; age 30-39 | 666 (18%) |
|  | 1973-1983; age 40-49 | 616 (17%) |
|  | 1963-1973; age 50-59 | 542 (15%) |
|  | 1953-1963; age 60-69 | 469 (13%) |
|  | 1943-1953; age 70-79 | 336 (9%) |
|  | 1943 or earlier; age 80+ | 79 (2%) |
| Gender | Male | 1791 (49%) |
|  | Female | 1872 (51%) |
|  | Other | 0 (0%) |
|  | Missing | 6 (0%) |
| Religious affiliation at age 12 | Christianity | 60 (2%) |
|  | Islam | 647 (18%) |
|  | Judaism | 2873 (78%) |
|  | Sikhism | 1 (0%) |
|  | Baha’i | 1 (0%) |
|  | Primal, Animist, or Folk religion | 3 (0%) |
|  | Some other religion | 5 (0%) |
|  | No religion/Atheist/Agnostic | 69 (2%) |
|  | Missing | 10 (0%) |
| Race/ethnicity | Jewish | 2926 (80%) |
|  | Arab | 674 (18%) |
|  | Other | 39 (1%) |
|  | Missing | 30 (1%) |

***Supplementary Table S9B. Regression of Promoting Good on Childhood Predictors for Israel***

| **Variable** | **Category** | **Estimate** | **SE** | **95% CI** | **Global P-value** |
| --- | --- | --- | --- | --- | --- |
| Relationship with mother (ref: Very bad/Somewhat bad) | Very good/Somewhat good | -0.25 | 0.17 | (-0.59,0.09) | 0.144 |
| Relationship with father (ref: Very bad/Somewhat bad) | Very good/Somewhat good | -0.06 | 0.15 | (-0.36,0.25) | 0.698 |
| Parent marital status (ref: Married) | Divorced | -0.25 | 0.13 | (-0.51,0.00) | 0.007 |
|  | Single, never married | -0.95 | 0.33 | (-1.61,-0.29) |  |
|  | One or both parents had died | -0.29 | 0.19 | (-0.66,0.09) |  |
| Subjective financial status of family growing up (ref: Got by) | Lived comfortably | 0.08 | 0.08 | (-0.07,0.23) | 0.242 |
|  | Found it difficult | 0.19 | 0.09 | (0.00,0.37) |  |
|  | Found it very difficult | 0.07 | 0.17 | (-0.26,0.40) |  |
| Felt like an outsider in the family (ref: No) | Yes | -0.27 | 0.14 | (-0.55,0.01) | 0.055 |
| Self-rated health growing up (ref: Good) | Excellent | 0.58 | 0.15 | (0.28,0.87) | <.001 |
|  | Very good | 0.56 | 0.13 | (0.31,0.80) |  |
|  | Fair | 0.01 | 0.21 | (-0.41,0.43) |  |
|  | Poor | 0.34 | 0.91 | (-1.46,2.15) |  |
| Immigration status (ref: Born in this country) | Born in another country | -0.37 | 0.10 | (-0.57,-0.18) | <.001 |
| Age 12 religious service attendance (ref: Never) | At least 1/week | 0.27 | 0.10 | (0.08,0.46) | 0.022 |
|  | 1-3/month | 0.31 | 0.12 | (0.06,0.55) |  |
|  | <1/month | 0.19 | 0.09 | (0.01,0.37) |  |
| Birth year (ref: 1998-2005; current age: 18-24) | 1993-1998; age 25-29 | 0.19 | 0.12 | (-0.06,0.43) | 0.858 |
|  | 1983-1993; age 30-39 | 0.19 | 0.13 | (-0.08,0.45) |  |
|  | 1973-1983; age 40-49 | 0.12 | 0.13 | (-0.15,0.38) |  |
|  | 1963-1973; age 50-59 | 0.17 | 0.12 | (-0.07,0.41) |  |
|  | 1953-1963; age 60-69 | 0.12 | 0.13 | (-0.14,0.38) |  |
|  | 1943-1953; age 70-79 | 0.07 | 0.15 | (-0.22,0.36) |  |
|  | 1943 or earlier; age 80+ | 0.02 | 0.24 | (-0.45,0.50) |  |
| Gender (ref:Male) | Female | 0.04 | 0.08 | (-0.12,0.20) | 0.708 |
|  | Other | -0.13 | 0.23 | (-0.59,0.33) |  |
| Religious affiliation at age 12 (ref: Judaism) | Islam | -1.04 | 0.29 | (-1.62,-0.47) | 0.002 |
|  | Collapsed affiliations with prevalence<3% | -0.40 | 0.23 | (-0.87,0.06) |  |
| Race/ethnicity (ref: plurality) | Race/ethnicity minority | 0.77 | 0.23 | (0.31,1.22) | <.001 |

***Supplementary Table S9C. E-Values for Estimates and CI for Israel***

| **Variable** | **Category** | **E-Value for Estimate** | **E-Value for 95% CI** |
| --- | --- | --- | --- |
| Relationship with mother (ref: Very bad/Somewhat bad) | Very good/Somewhat good | 1.57 | 1.00 |
| Relationship with father (ref: Very bad/Somewhat bad) | Very good/Somewhat good | 1.22 | 1.00 |
| Parent marital status (ref: Married) | Divorced | 1.58 | 1.00 |
|  | Single, never married | 2.83 | 1.66 |
|  | One or both parents had died | 1.63 | 1.00 |
| Subjective financial status of family growing up (ref: Got by) | Lived comfortably | 1.27 | 1.00 |
|  | Found it difficult | 1.46 | 1.06 |
|  | Found it very difficult | 1.25 | 1.00 |
| Felt like an outsider in the family (ref: No) | Yes | 1.61 | 1.00 |
| Self-rated health growing up (ref: Good) | Excellent | 2.12 | 1.63 |
|  | Very good | 2.09 | 1.67 |
|  | Fair | 1.06 | 1.00 |
|  | Poor | 1.73 | 1.00 |
| Immigration status (ref: Born in this country) | Born in another country | 1.78 | 1.45 |
| Age 12 religious service attendance (ref: Never) | At least 1/week | 1.60 | 1.27 |
|  | 1-3/month | 1.66 | 1.23 |
|  | <1/month | 1.47 | 1.08 |
| Birth year (ref: 1998-2005; current age: 18-24) | 1993-1998; age 25-29 | 1.47 | 1.00 |
|  | 1983-1993; age 30-39 | 1.47 | 1.00 |
|  | 1973-1983; age 40-49 | 1.34 | 1.00 |
|  | 1963-1973; age 50-59 | 1.44 | 1.00 |
|  | 1953-1963; age 60-69 | 1.34 | 1.00 |
|  | 1943-1953; age 70-79 | 1.25 | 1.00 |
|  | 1943 or earlier; age 80+ | 1.13 | 1.00 |
| Gender (ref:Male) | Female | 1.17 | 1.00 |
|  | Other | 1.36 | 1.00 |
| Religious affiliation at age 12 (ref: Judaism) | Islam | 3.03 | 1.94 |
|  | Collapsed affiliations with prevalence<3% | 1.83 | 1.00 |
| Race/ethnicity (ref: plurality) | Race/ethnicity minority | 2.46 | 1.69 |

***Supplementary Table S10A. Nationally Representative Childhood Descriptive Statistics for Japan***

| **Variable** | **Category** | **N (%)** |
| --- | --- | --- |
| Relationship with mother | Very good | 5630 (27%) |
|  | Somewhat good | 9461 (46%) |
|  | Somewhat bad | 2750 (13%) |
|  | Very bad | 799 (4%) |
|  | Does not apply | 1838 (9%) |
|  | Missing | 66 (0%) |
| Relationships with father | Very good | 4156 (20%) |
|  | Somewhat good | 9081 (44%) |
|  | Somewhat bad | 3446 (17%) |
|  | Very bad | 1223 (6%) |
|  | Does not apply | 2580 (13%) |
|  | Missing | 57 (0%) |
| Parent marital status | Parents married | 17713 (86%) |
|  | Divorced | 1127 (5%) |
|  | Single, never married | 591 (3%) |
|  | One or both parents had died | 754 (4%) |
|  | Missing | 359 (2%) |
| Subjective financial status of family growing up | Lived comfortably | 8320 (41%) |
|  | Got by | 8799 (43%) |
|  | Found it difficult | 2398 (12%) |
|  | Found it very difficult | 973 (5%) |
|  | Missing | 52 (0%) |
| Abuse | Yes | 1482 (7%) |
|  | No | 18964 (92%) |
|  | Missing | 96 (0%) |
| Felt like an outsider in the family | Yes | 1963 (10%) |
|  | No | 17136 (83%) |
|  | Missing | 1444 (7%) |
| Self-rated health growing up | Excellent | 2711 (13%) |
|  | Very good | 7106 (35%) |
|  | Good | 6689 (33%) |
|  | Fair | 3199 (16%) |
|  | Poor | 758 (4%) |
|  | Missing | 80 (0%) |
| Immigration status | Born in this country | 19548 (95%) |
|  | Born in another country | 158 (1%) |
|  | Missing | 837 (4%) |
| Age 12 religious service attendance | At least 1/week | 398 (2%) |
|  | 1-3/month | 883 (4%) |
|  | <1/ month | 5023 (24%) |
|  | Never | 14117 (69%) |
|  | Missing | 123 (1%) |
| Year of birth | 1998-2005; current age: 18-24 | 1589 (8%) |
|  | 1993-1998; age 25-29 | 806 (4%) |
|  | 1983-1993; age 30-39 | 2851 (14%) |
|  | 1973-1983; age 40-49 | 3363 (16%) |
|  | 1963-1973; age 50-59 | 3770 (18%) |
|  | 1953-1963; age 60-69 | 4118 (20%) |
|  | 1943-1953; age 70-79 | 3554 (17%) |
|  | 1943 or earlier; age 80+ | 493 (2%) |
| Gender | Male | 9847 (48%) |
|  | Female | 10602 (52%) |
|  | Other | 28 (0%) |
|  | Missing | 66 (0%) |
| Religious affiliation at age 12 | Christianity | 343 (2%) |
|  | Islam | 7 (0%) |
|  | Hinduism | 4 (0%) |
|  | Buddhism | 6536 (32%) |
|  | Baha’i | 7 (0%) |
|  | Jainism | 1 (0%) |
|  | Shinto | 382 (2%) |
|  | Taoism | 14 (0%) |
|  | Confucianism | 25 (0%) |
|  | Primal, Animist, or Folk religion | 13 (0%) |
|  | Some other religion | 46 (0%) |
|  | No religion/Atheist/Agnostic | 12950 (63%) |
|  | Missing | 215 (1%) |

***Supplementary Table S10B. Regression of Promoting Good on Childhood Predictors for Japan***

| **Variable** | **Category** | **Estimate** | **SE** | **95% CI** | **Global P-value** |
| --- | --- | --- | --- | --- | --- |
| Relationship with mother (ref: Very bad/Somewhat bad) | Very good/Somewhat good | 0.13 | 0.05 | (0.04,0.22) | 0.004 |
| Relationship with father (ref: Very bad/Somewhat bad) | Very good/Somewhat good | 0.27 | 0.04 | (0.19,0.35) | <.001 |
| Parent marital status (ref: Married) | Divorced | 0.17 | 0.08 | (0.01,0.32) | 0.094 |
|  | Single, never married | 0.05 | 0.09 | (-0.12,0.23) |  |
|  | One or both parents had died | 0.12 | 0.09 | (-0.05,0.30) |  |
| Subjective financial status of family growing up (ref: Got by) | Lived comfortably | 0.32 | 0.03 | (0.25,0.39) | <.001 |
|  | Found it difficult | -0.07 | 0.05 | (-0.17,0.03) |  |
|  | Found it very difficult | -0.35 | 0.10 | (-0.54,-0.16) |  |
| Abuse (ref: No) | Yes | -0.00 | 0.08 | (-0.15,0.15) | 0.892 |
| Felt like an outsider in the family (ref: No) | Yes | -0.07 | 0.06 | (-0.20,0.06) | 0.260 |
| Self-rated health growing up (ref: Good) | Excellent | 1.34 | 0.05 | (1.23,1.45) | <.001 |
|  | Very good | 0.54 | 0.04 | (0.47,0.61) |  |
|  | Fair | -0.46 | 0.05 | (-0.56,-0.37) |  |
|  | Poor | -0.54 | 0.11 | (-0.76,-0.31) |  |
| Immigration status (ref: Born in this country) | Born in another country | 0.42 | 0.15 | (0.12,0.72) | 0.006 |
| Age 12 religious service attendance (ref: Never) | At least 1/week | 0.51 | 0.12 | (0.28,0.74) | <.001 |
|  | 1-3/month | 0.57 | 0.08 | (0.42,0.73) |  |
|  | <1/month | 0.18 | 0.04 | (0.10,0.25) |  |
| Birth year (ref: 1998-2005; current age: 18-24) | 1993-1998; age 25-29 | -0.17 | 0.10 | (-0.36,0.02) | <.001 |
|  | 1983-1993; age 30-39 | -0.07 | 0.07 | (-0.21,0.08) |  |
|  | 1973-1983; age 40-49 | -0.03 | 0.07 | (-0.17,0.11) |  |
|  | 1963-1973; age 50-59 | 0.20 | 0.07 | (0.06,0.33) |  |
|  | 1953-1963; age 60-69 | 0.57 | 0.07 | (0.44,0.70) |  |
|  | 1943-1953; age 70-79 | 1.06 | 0.07 | (0.93,1.19) |  |
|  | 1943 or earlier; age 80+ | 1.29 | 0.11 | (1.07,1.51) |  |
| Gender (ref:Male) | Female | 0.15 | 0.03 | (0.09,0.21) | <.001 |
|  | Other | -0.03 | 0.45 | (-0.91,0.85) |  |
| Religious affiliation at age 12 (ref: No religion/Atheist/Agnostic) | Buddhism | 0.19 | 0.03 | (0.12,0.26) | <.001 |
|  | Collapsed affiliations with prevalence<3% | 0.24 | 0.08 | (0.08,0.40) |  |

***Supplementary Table S10C. E-Values for Estimates and CI for Japan***

| **Variable** | **Category** | **E-Value for Estimate** | **E-Value for 95% CI** |
| --- | --- | --- | --- |
| Relationship with mother (ref: Very bad/Somewhat bad) | Very good/Somewhat good | 1.31 | 1.15 |
| Relationship with father (ref: Very bad/Somewhat bad) | Very good/Somewhat good | 1.50 | 1.40 |
| Parent marital status (ref: Married) | Divorced | 1.36 | 1.08 |
|  | Single, never married | 1.18 | 1.00 |
|  | One or both parents had died | 1.30 | 1.00 |
| Subjective financial status of family growing up (ref: Got by) | Lived comfortably | 1.57 | 1.48 |
|  | Found it difficult | 1.21 | 1.00 |
|  | Found it very difficult | 1.61 | 1.35 |
| Abuse (ref: No) | Yes | 1.04 | 1.00 |
| Felt like an outsider in the family (ref: No) | Yes | 1.21 | 1.00 |
| Self-rated health growing up (ref: Good) | Excellent | 3.01 | 2.83 |
|  | Very good | 1.86 | 1.77 |
|  | Fair | 1.75 | 1.63 |
|  | Poor | 1.85 | 1.56 |
| Immigration status (ref: Born in this country) | Born in another country | 1.70 | 1.29 |
| Age 12 religious service attendance (ref: Never) | At least 1/week | 1.82 | 1.52 |
|  | 1-3/month | 1.90 | 1.70 |
|  | <1/month | 1.38 | 1.27 |
| Birth year (ref: 1998-2005; current age: 18-24) | 1993-1998; age 25-29 | 1.37 | 1.00 |
|  | 1983-1993; age 30-39 | 1.21 | 1.00 |
|  | 1973-1983; age 40-49 | 1.12 | 1.00 |
|  | 1963-1973; age 50-59 | 1.41 | 1.20 |
|  | 1953-1963; age 60-69 | 1.89 | 1.72 |
|  | 1943-1953; age 70-79 | 2.57 | 2.38 |
|  | 1943 or earlier; age 80+ | 2.93 | 2.59 |
| Gender (ref:Male) | Female | 1.34 | 1.24 |
|  | Other | 1.13 | 1.00 |
| Religious affiliation at age 12 (ref: No religion/Atheist/Agnostic) | Buddhism | 1.40 | 1.30 |
|  | Collapsed affiliations with prevalence<3% | 1.47 | 1.23 |

***Supplementary Table11A. Nationally Representative Childhood Descriptive Statistics for Kenya***

| **Variable** | **Category** | **N (%)** |
| --- | --- | --- |
| Relationship with mother | Very good | 9418 (83%) |
|  | Somewhat good | 1435 (13%) |
|  | Somewhat bad | 130 (1%) |
|  | Very bad | 100 (1%) |
|  | Does not apply | 240 (2%) |
|  | Missing | 66 (1%) |
| Relationships with father | Very good | 7958 (70%) |
|  | Somewhat good | 1896 (17%) |
|  | Somewhat bad | 216 (2%) |
|  | Very bad | 220 (2%) |
|  | Does not apply | 967 (8%) |
|  | Missing | 132 (1%) |
| Parent marital status | Parents married | 9238 (81%) |
|  | Divorced | 697 (6%) |
|  | Single, never married | 681 (6%) |
|  | One or both parents had died | 471 (4%) |
|  | Missing | 301 (3%) |
| Subjective financial status of family growing up | Lived comfortably | 3026 (27%) |
|  | Got by | 3279 (29%) |
|  | Found it difficult | 4071 (36%) |
|  | Found it very difficult | 994 (9%) |
|  | Missing | 19 (0%) |
| Abuse | Yes | 1300 (11%) |
|  | No | 10039 (88%) |
|  | Missing | 49 (0%) |
| Felt like an outsider in the family | Yes | 1223 (11%) |
|  | No | 10114 (89%) |
|  | Missing | 52 (0%) |
| Self-rated health growing up | Excellent | 4449 (39%) |
|  | Very good | 2598 (23%) |
|  | Good | 2582 (23%) |
|  | Fair | 1384 (12%) |
|  | Poor | 349 (3%) |
|  | Missing | 26 (0%) |
| Immigration status | Born in this country | 11270 (99%) |
|  | Born in another country | 117 (1%) |
|  | Missing | 2 (0%) |
| Age 12 religious service attendance | At least 1/week | 9189 (81%) |
|  | 1-3/month | 1687 (15%) |
|  | <1/ month | 236 (2%) |
|  | Never | 198 (2%) |
|  | Missing | 79 (1%) |
| Year of birth | 1998-2005; current age: 18-24 | 2868 (25%) |
|  | 1993-1998; age 25-29 | 2035 (18%) |
|  | 1983-1993; age 30-39 | 2564 (23%) |
|  | 1973-1983; age 40-49 | 1708 (15%) |
|  | 1963-1973; age 50-59 | 1072 (9%) |
|  | 1953-1963; age 60-69 | 710 (6%) |
|  | 1943-1953; age 70-79 | 360 (3%) |
|  | 1943 or earlier; age 80+ | 67 (1%) |
|  | Missing | 5 (0%) |
| Gender | Male | 5567 (49%) |
|  | Female | 5813 (51%) |
|  | Other | 2 (0%) |
|  | Missing | 7 (0%) |
| Religious affiliation at age 12 | Christianity | 10369 (91%) |
|  | Islam | 916 (8%) |
|  | Buddhism | 5 (0%) |
|  | Judaism | 6 (0%) |
|  | Sikhism | 0 (0%) |
|  | Baha’i | 3 (0%) |
|  | Jainism | 1 (0%) |
|  | Primal, Animist, or Folk religion | 13 (0%) |
|  | Some other religion | 0 (0%) |
|  | No religion/Atheist/Agnostic | 67 (1%) |
|  | Missing | 9 (0%) |
| Race/ethnicity | Luhya | 1943 (17%) |
|  | Luo | 1120 (10%) |
|  | Kalenjin | 1377 (12%) |
|  | Kamba | 1299 (11%) |
|  | Kikuyu | 2119 (19%) |
|  | Kisii | 789 (7%) |
|  | Maasai | 237 (2%) |
|  | Meru | 630 (6%) |
|  | Kenyan Somali/Somali | 396 (3%) |
|  | Miji Kenda tribes | 708 (6%) |
|  | Embu | 197 (2%) |
|  | Other | 548 (5%) |
|  | Missing | 27 (0%) |

***Supplementary Table S11B. Regression of Promoting Good on Childhood Predictors for Kenya***

| **Variable** | **Category** | **Estimate** | **SE** | **95% CI** | **Global P-value** |
| --- | --- | --- | --- | --- | --- |
| Relationship with mother (ref: Very bad/Somewhat bad) | Very good/Somewhat good | 0.07 | 0.22 | (-0.36,0.50) | 0.742 |
| Relationship with father (ref: Very bad/Somewhat bad) | Very good/Somewhat good | -0.07 | 0.16 | (-0.38,0.25) | 0.675 |
| Parent marital status (ref: Married) | Divorced | -0.26 | 0.15 | (-0.56,0.04) | <.001 |
|  | Single, never married | -0.64 | 0.16 | (-0.95,-0.33) |  |
|  | One or both parents had died | -0.11 | 0.19 | (-0.49,0.26) |  |
| Subjective financial status of family growing up (ref: Got by) | Lived comfortably | 0.12 | 0.09 | (-0.05,0.30) | 0.295 |
|  | Found it difficult | 0.01 | 0.08 | (-0.15,0.16) |  |
|  | Found it very difficult | -0.11 | 0.14 | (-0.38,0.16) |  |
| Abuse (ref: No) | Yes | -0.46 | 0.11 | (-0.67,-0.24) | <.001 |
| Felt like an outsider in the family (ref: No) | Yes | -0.25 | 0.13 | (-0.50,0.01) | 0.055 |
| Self-rated health growing up (ref: Good) | Excellent | 0.20 | 0.09 | (0.02,0.37) | 0.145 |
|  | Very good | 0.13 | 0.09 | (-0.05,0.30) |  |
|  | Fair | 0.02 | 0.12 | (-0.22,0.26) |  |
|  | Poor | 0.03 | 0.26 | (-0.48,0.54) |  |
| Immigration status (ref: Born in this country) | Born in another country | -0.29 | 0.32 | (-0.92,0.34) | 0.369 |
| Age 12 religious service attendance (ref: Never) | At least 1/week | -0.14 | 0.31 | (-0.74,0.47) | 0.492 |
|  | 1-3/month | -0.27 | 0.32 | (-0.90,0.36) |  |
|  | <1/month | -0.14 | 0.32 | (-0.76,0.48) |  |
| Birth year (ref: 1998-2005; current age: 18-24) | 1993-1998; age 25-29 | -0.05 | 0.07 | (-0.20,0.10) | 0.023 |
|  | 1983-1993; age 30-39 | -0.14 | 0.08 | (-0.31,0.02) |  |
|  | 1973-1983; age 40-49 | -0.17 | 0.09 | (-0.35,0.02) |  |
|  | 1963-1973; age 50-59 | -0.36 | 0.14 | (-0.62,-0.09) |  |
|  | 1953-1963; age 60-69 | -0.54 | 0.17 | (-0.87,-0.21) |  |
|  | 1943-1953; age 70-79 | -0.32 | 0.28 | (-0.87,0.24) |  |
|  | 1943 or earlier; age 80+ | -0.03 | 0.48 | (-0.98,0.92) |  |
| Gender (ref:Male) | Female | -0.05 | 0.06 | (-0.16,0.07) | <.001 |
|  | Other | 1.99 | 0.16 | (1.68,2.31) |  |
| Religious affiliation at age 12 (ref: Christianity) | Islam | -0.06 | 0.24 | (-0.53,0.41) | 0.967 |
|  | Collapsed affiliations with prevalence<3% | 0.00 | 0.45 | (-0.89,0.89) |  |
| Race/ethnicity (ref: plurality) | Race/ethnicity minority | -0.08 | 0.09 | (-0.26,0.11) | 0.416 |

***Supplementary Table S11C. E-Values for Estimates and CI for Kenya***

| **Variable** | **Category** | **E-Value for Estimate** | **E-Value for 95% CI** |
| --- | --- | --- | --- |
| Relationship with mother (ref: Very bad/Somewhat bad) | Very good/Somewhat good | 1.18 | 1.00 |
| Relationship with father (ref: Very bad/Somewhat bad) | Very good/Somewhat good | 1.18 | 1.00 |
| Parent marital status (ref: Married) | Divorced | 1.41 | 1.00 |
|  | Single, never married | 1.79 | 1.48 |
|  | One or both parents had died | 1.24 | 1.00 |
| Subjective financial status of family growing up (ref: Got by) | Lived comfortably | 1.26 | 1.00 |
|  | Found it difficult | 1.06 | 1.00 |
|  | Found it very difficult | 1.24 | 1.00 |
| Abuse (ref: No) | Yes | 1.61 | 1.39 |
| Felt like an outsider in the family (ref: No) | Yes | 1.40 | 1.00 |
| Self-rated health growing up (ref: Good) | Excellent | 1.34 | 1.10 |
|  | Very good | 1.26 | 1.00 |
|  | Fair | 1.09 | 1.00 |
|  | Poor | 1.10 | 1.00 |
| Immigration status (ref: Born in this country) | Born in another country | 1.44 | 1.00 |
| Age 12 religious service attendance (ref: Never) | At least 1/week | 1.27 | 1.00 |
|  | 1-3/month | 1.42 | 1.00 |
|  | <1/month | 1.27 | 1.00 |
| Birth year (ref: 1998-2005; current age: 18-24) | 1993-1998; age 25-29 | 1.15 | 1.00 |
|  | 1983-1993; age 30-39 | 1.28 | 1.00 |
|  | 1973-1983; age 40-49 | 1.31 | 1.00 |
|  | 1963-1973; age 50-59 | 1.51 | 1.22 |
|  | 1953-1963; age 60-69 | 1.69 | 1.36 |
|  | 1943-1953; age 70-79 | 1.47 | 1.00 |
|  | 1943 or earlier; age 80+ | 1.11 | 1.00 |
| Gender (ref:Male) | Female | 1.14 | 1.00 |
|  | Other | 3.35 | 2.94 |
| Religious affiliation at age 12 (ref: Christianity) | Islam | 1.16 | 1.00 |
|  | Collapsed affiliations with prevalence<3% | 1.03 | 1.00 |
| Race/ethnicity (ref: plurality) | Race/ethnicity minority | 1.19 | 1.00 |

***Supplementary Table S12A. Nationally Representative Childhood Descriptive Statistics for Mexico***

| **Variable** | **Category** | **N (%)** |
| --- | --- | --- |
| Relationship with mother | Very good | 3912 (68%) |
|  | Somewhat good | 1340 (23%) |
|  | Somewhat bad | 177 (3%) |
|  | Very bad | 90 (2%) |
|  | Does not apply | 177 (3%) |
|  | Missing | 80 (1%) |
| Relationships with father | Very good | 3089 (53%) |
|  | Somewhat good | 1556 (27%) |
|  | Somewhat bad | 335 (6%) |
|  | Very bad | 267 (5%) |
|  | Does not apply | 470 (8%) |
|  | Missing | 60 (1%) |
| Parent marital status | Parents married | 3999 (69%) |
|  | Divorced | 341 (6%) |
|  | Single, never married | 827 (14%) |
|  | One or both parents had died | 176 (3%) |
|  | Missing | 432 (7%) |
| Subjective financial status of family growing up | Lived comfortably | 1775 (31%) |
|  | Got by | 1872 (32%) |
|  | Found it difficult | 1712 (30%) |
|  | Found it very difficult | 369 (6%) |
|  | Missing | 48 (1%) |
| Abuse | Yes | 905 (16%) |
|  | No | 4604 (80%) |
|  | Missing | 267 (5%) |
| Felt like an outsider in the family | Yes | 772 (13%) |
|  | No | 4897 (85%) |
|  | Missing | 107 (2%) |
| Self-rated health growing up | Excellent | 1860 (32%) |
|  | Very good | 1350 (23%) |
|  | Good | 1677 (29%) |
|  | Fair | 743 (13%) |
|  | Poor | 133 (2%) |
|  | Missing | 14 (0%) |
| Immigration status | Born in this country | 5517 (96%) |
|  | Born in another country | 108 (2%) |
|  | Missing | 151 (3%) |
| Age 12 religious service attendance | At least 1/week | 2514 (44%) |
|  | 1-3/month | 1162 (20%) |
|  | <1/ month | 1087 (19%) |
|  | Never | 944 (16%) |
|  | Missing | 69 (1%) |
| Year of birth | 1998-2005; current age: 18-24 | 986 (17%) |
|  | 1993-1998; age 25-29 | 623 (11%) |
|  | 1983-1993; age 30-39 | 1312 (23%) |
|  | 1973-1983; age 40-49 | 1027 (18%) |
|  | 1963-1973; age 50-59 | 873 (15%) |
|  | 1953-1963; age 60-69 | 611 (11%) |
|  | 1943-1953; age 70-79 | 277 (5%) |
|  | 1943 or earlier; age 80+ | 68 (1%) |
| Gender | Male | 2755 (48%) |
|  | Female | 2997 (52%) |
|  | Other | 3 (0%) |
|  | Missing | 21 (0%) |
| Religious affiliation at age 12 | Christianity | 5337 (92%) |
|  | Islam | 6 (0%) |
|  | Hinduism | 1 (0%) |
|  | Buddhism | 1 (0%) |
|  | Judaism | 8 (0%) |
|  | Sikhism | 4 (0%) |
|  | Baha’i | 1 (0%) |
|  | Shinto | 2 (0%) |
|  | Taoism | 5 (0%) |
|  | Primal, Animist, or Folk religion | 2 (0%) |
|  | Some other religion | 7 (0%) |
|  | No religion/Atheist/Agnostic | 328 (6%) |
|  | Missing | 74 (1%) |
| Race/ethnicity | White | 1116 (19%) |
|  | Mestizo | 2762 (48%) |
|  | Indigenous | 594 (10%) |
|  | Black | 108 (2%) |
|  | Mulatto | 63 (1%) |
|  | Other | 339 (6%) |
|  | Missing | 794 (14%) |

***Supplementary Table S12B. Regression of Promoting Good on Childhood Predictors for Mexico***

| **Variable** | **Category** | **Estimate** | **SE** | **95% CI** | **Global P-value** |
| --- | --- | --- | --- | --- | --- |
| Relationship with mother (ref: Very bad/Somewhat bad) | Very good/Somewhat good | 0.15 | 0.13 | (-0.10,0.40) | 0.213 |
| Relationship with father (ref: Very bad/Somewhat bad) | Very good/Somewhat good | 0.02 | 0.09 | (-0.16,0.20) | 0.822 |
| Parent marital status (ref: Married) | Divorced | 0.08 | 0.12 | (-0.17,0.33) | 0.523 |
|  | Single, never married | -0.08 | 0.09 | (-0.26,0.10) |  |
|  | One or both parents had died | -0.08 | 0.16 | (-0.39,0.22) |  |
| Subjective financial status of family growing up (ref: Got by) | Lived comfortably | 0.28 | 0.06 | (0.16,0.40) | <.001 |
|  | Found it difficult | 0.17 | 0.07 | (0.03,0.30) |  |
|  | Found it very difficult | 0.12 | 0.16 | (-0.20,0.44) |  |
| Abuse (ref: No) | Yes | -0.12 | 0.07 | (-0.26,0.03) | 0.091 |
| Felt like an outsider in the family (ref: No) | Yes | -0.22 | 0.10 | (-0.41,-0.02) | 0.028 |
| Self-rated health growing up (ref: Good) | Excellent | 0.36 | 0.06 | (0.23,0.48) | <.001 |
|  | Very good | 0.12 | 0.07 | (-0.02,0.25) |  |
|  | Fair | -0.15 | 0.10 | (-0.35,0.05) |  |
|  | Poor | 0.18 | 0.19 | (-0.19,0.55) |  |
| Immigration status (ref: Born in this country) | Born in another country | -0.41 | 0.18 | (-0.77,-0.06) | 0.022 |
| Age 12 religious service attendance (ref: Never) | At least 1/week | 0.25 | 0.08 | (0.09,0.41) | <.001 |
|  | 1-3/month | -0.03 | 0.09 | (-0.21,0.16) |  |
|  | <1/month | 0.03 | 0.09 | (-0.15,0.21) |  |
| Birth year (ref: 1998-2005; current age: 18-24) | 1993-1998; age 25-29 | 0.17 | 0.09 | (-0.01,0.34) | <.001 |
|  | 1983-1993; age 30-39 | 0.36 | 0.08 | (0.20,0.51) |  |
|  | 1973-1983; age 40-49 | 0.43 | 0.08 | (0.27,0.60) |  |
|  | 1963-1973; age 50-59 | 0.42 | 0.09 | (0.24,0.61) |  |
|  | 1953-1963; age 60-69 | 0.36 | 0.10 | (0.16,0.57) |  |
|  | 1943-1953; age 70-79 | 0.26 | 0.15 | (-0.03,0.55) |  |
|  | 1943 or earlier; age 80+ | -0.13 | 0.33 | (-0.79,0.52) |  |
| Gender (ref:Male) | Female | -0.02 | 0.05 | (-0.12,0.09) | 0.938 |
|  | Other | 0.01 | 0.56 | (-1.07,1.10) |  |
| Religious affiliation at age 12 (ref: No religion/Atheist/Agnostic) | Christianity | 0.10 | 0.12 | (-0.14,0.33) | 0.572 |
|  | Collapsed affiliations with prevalence<3% | -0.19 | 0.44 | (-1.06,0.68) |  |
| Race/ethnicity (ref: plurality) | Race/ethnicity minority | -0.07 | 0.06 | (-0.19,0.06) | 0.167 |

***Supplementary Table S12C. E-Values for Estimates and CI for Mexico***

| **Variable** | **Category** | **E-Value for Estimate** | **E-Value for 95% CI** |
| --- | --- | --- | --- |
| Relationship with mother (ref: Very bad/Somewhat bad) | Very good/Somewhat good | 1.40 | 1.00 |
| Relationship with father (ref: Very bad/Somewhat bad) | Very good/Somewhat good | 1.11 | 1.00 |
| Parent marital status (ref: Married) | Divorced | 1.26 | 1.00 |
|  | Single, never married | 1.27 | 1.00 |
|  | One or both parents had died | 1.27 | 1.00 |
| Subjective financial status of family growing up (ref: Got by) | Lived comfortably | 1.63 | 1.43 |
|  | Found it difficult | 1.43 | 1.15 |
|  | Found it very difficult | 1.34 | 1.00 |
| Abuse (ref: No) | Yes | 1.34 | 1.00 |
| Felt like an outsider in the family (ref: No) | Yes | 1.52 | 1.13 |
| Self-rated health growing up (ref: Good) | Excellent | 1.75 | 1.54 |
|  | Very good | 1.34 | 1.00 |
|  | Fair | 1.40 | 1.00 |
|  | Poor | 1.45 | 1.00 |
| Immigration status (ref: Born in this country) | Born in another country | 1.85 | 1.22 |
| Age 12 religious service attendance (ref: Never) | At least 1/week | 1.58 | 1.29 |
|  | 1-3/month | 1.14 | 1.00 |
|  | <1/month | 1.15 | 1.00 |
| Birth year (ref: 1998-2005; current age: 18-24) | 1993-1998; age 25-29 | 1.43 | 1.00 |
|  | 1983-1993; age 30-39 | 1.75 | 1.49 |
|  | 1973-1983; age 40-49 | 1.88 | 1.61 |
|  | 1963-1973; age 50-59 | 1.86 | 1.55 |
|  | 1953-1963; age 60-69 | 1.76 | 1.42 |
|  | 1943-1953; age 70-79 | 1.60 | 1.00 |
|  | 1943 or earlier; age 80+ | 1.37 | 1.00 |
| Gender (ref:Male) | Female | 1.11 | 1.00 |
|  | Other | 1.10 | 1.00 |
| Religious affiliation at age 12 (ref: No religion/Atheist/Agnostic) | Christianity | 1.30 | 1.00 |
|  | Collapsed affiliations with prevalence<3% | 1.47 | 1.00 |
| Race/ethnicity (ref: plurality) | Race/ethnicity minority | 1.24 | 1.00 |

***Supplementary Table S13A. Nationally Representative Childhood Descriptive Statistics for Nigeria***

| **Variable** | **Category** | **N (%)** |
| --- | --- | --- |
| Relationship with mother | Very good | 5986 (88%) |
|  | Somewhat good | 648 (9%) |
|  | Somewhat bad | 62 (1%) |
|  | Very bad | 18 (0%) |
|  | Does not apply | 104 (2%) |
|  | Missing | 9 (0%) |
| Relationships with father | Very good | 5578 (82%) |
|  | Somewhat good | 924 (14%) |
|  | Somewhat bad | 76 (1%) |
|  | Very bad | 43 (1%) |
|  | Does not apply | 177 (3%) |
|  | Missing | 29 (0%) |
| Parent marital status | Parents married | 5568 (82%) |
|  | Divorced | 307 (5%) |
|  | Single, never married | 335 (5%) |
|  | One or both parents had died | 462 (7%) |
|  | Missing | 154 (2%) |
| Subjective financial status of family growing up | Lived comfortably | 2192 (32%) |
|  | Got by | 2381 (35%) |
|  | Found it difficult | 1661 (24%) |
|  | Found it very difficult | 563 (8%) |
|  | Missing | 29 (0%) |
| Abuse | Yes | 880 (13%) |
|  | No | 5851 (86%) |
|  | Missing | 96 (1%) |
| Felt like an outsider in the family | Yes | 669 (10%) |
|  | No | 6059 (89%) |
|  | Missing | 99 (1%) |
| Self-rated health growing up | Excellent | 2644 (39%) |
|  | Very good | 2613 (38%) |
|  | Good | 1152 (17%) |
|  | Fair | 306 (4%) |
|  | Poor | 98 (1%) |
|  | Missing | 14 (0%) |
| Immigration status | Born in this country | 6779 (99%) |
|  | Born in another country | 47 (1%) |
|  | Missing | 1 (0%) |
| Age 12 religious service attendance | At least 1/week | 5907 (87%) |
|  | 1-3/month | 600 (9%) |
|  | <1/ month | 136 (2%) |
|  | Never | 138 (2%) |
|  | Missing | 45 (1%) |
| Year of birth | 1998-2005; current age: 18-24 | 1533 (22%) |
|  | 1993-1998; age 25-29 | 1193 (17%) |
|  | 1983-1993; age 30-39 | 1943 (28%) |
|  | 1973-1983; age 40-49 | 1059 (16%) |
|  | 1963-1973; age 50-59 | 619 (9%) |
|  | 1953-1963; age 60-69 | 296 (4%) |
|  | 1943-1953; age 70-79 | 133 (2%) |
|  | 1943 or earlier; age 80+ | 50 (1%) |
| Gender | Male | 3371 (49%) |
|  | Female | 3456 (51%) |
|  | Other | 0 (0%) |
| Religious affiliation at age 12 | Christianity | 3463 (51%) |
|  | Islam | 3314 (49%) |
|  | Buddhism | 0 (0%) |
|  | Confucianism | 0 (0%) |
|  | Primal, Animist, or Folk religion | 17 (0%) |
|  | No religion/Atheist/Agnostic | 19 (0%) |
|  | Missing | 14 (0%) |
| Race/ethnicity | Hausa | 2342 (34%) |
|  | Yoruba | 1230 (18%) |
|  | Igbo (Ibo) | 1111 (16%) |
|  | Edo | 116 (2%) |
|  | Urhobo | 38 (1%) |
|  | Fulani | 266 (4%) |
|  | Kanuri | 31 (0%) |
|  | Tiv | 198 (3%) |
|  | Efik | 48 (1%) |
|  | Ijaw | 110 (2%) |
|  | Igala | 77 (1%) |
|  | Ibibio | 180 (3%) |
|  | Idoma | 61 (1%) |
|  | Other | 1014 (15%) |
|  | Missing | 4 (0%) |

***Supplementary Table S13B. Regression of Promoting Good on Childhood Predictors for Nigeria***

| **Variable** | **Category** | **Estimate** | **SE** | **95% CI** | **Global P-value** |
| --- | --- | --- | --- | --- | --- |
| Relationship with mother (ref: Very bad/Somewhat bad) | Very good/Somewhat good | -0.08 | 0.28 | (-0.62,0.46) | 0.761 |
| Relationship with father (ref: Very bad/Somewhat bad) | Very good/Somewhat good | -0.13 | 0.22 | (-0.56,0.29) | 0.535 |
| Parent marital status (ref: Married) | Divorced | 0.01 | 0.19 | (-0.37,0.39) | 0.006 |
|  | Single, never married | -0.37 | 0.18 | (-0.72,-0.02) |  |
|  | One or both parents had died | -0.50 | 0.17 | (-0.84,-0.16) |  |
| Subjective financial status of family growing up (ref: Got by) | Lived comfortably | -0.02 | 0.10 | (-0.22,0.18) | 0.419 |
|  | Found it difficult | 0.11 | 0.10 | (-0.08,0.31) |  |
|  | Found it very difficult | -0.14 | 0.18 | (-0.49,0.20) |  |
| Abuse (ref: No) | Yes | -0.28 | 0.12 | (-0.52,-0.03) | 0.025 |
| Felt like an outsider in the family (ref: No) | Yes | -0.04 | 0.12 | (-0.27,0.19) | 0.713 |
| Self-rated health growing up (ref: Good) | Excellent | 0.08 | 0.12 | (-0.15,0.31) | 0.554 |
|  | Very good | 0.04 | 0.11 | (-0.18,0.25) |  |
|  | Fair | -0.26 | 0.19 | (-0.63,0.12) |  |
|  | Poor | 0.03 | 0.29 | (-0.55,0.60) |  |
| Immigration status (ref: Born in this country) | Born in another country | -0.76 | 0.37 | (-1.48,-0.04) | 0.038 |
| Age 12 religious service attendance (ref: Never) | At least 1/week | -0.08 | 0.24 | (-0.56,0.41) | 0.032 |
|  | 1-3/month | -0.11 | 0.28 | (-0.67,0.45) |  |
|  | <1/month | -0.77 | 0.32 | (-1.40,-0.13) |  |
| Birth year (ref: 1998-2005; current age: 18-24) | 1993-1998; age 25-29 | 0.06 | 0.10 | (-0.14,0.25) | 0.004 |
|  | 1983-1993; age 30-39 | 0.06 | 0.10 | (-0.13,0.25) |  |
|  | 1973-1983; age 40-49 | 0.04 | 0.12 | (-0.21,0.28) |  |
|  | 1963-1973; age 50-59 | 0.37 | 0.16 | (0.06,0.68) |  |
|  | 1953-1963; age 60-69 | 0.17 | 0.29 | (-0.40,0.74) |  |
|  | 1943-1953; age 70-79 | 0.32 | 0.43 | (-0.53,1.16) |  |
|  | 1943 or earlier; age 80+ | -2.13 | 1.07 | (-4.23,-0.03) |  |
| Gender (ref:Male) | Female | -0.01 | 0.07 | (-0.15,0.14) | 0.565 |
|  | Other | -0.17 | 0.17 | (-0.50,0.15) |  |
| Religious affiliation at age 12 (ref: Christianity) | Islam | -0.30 | 0.10 | (-0.50,-0.10) | 0.014 |
|  | Collapsed affiliations with prevalence<3% | -0.09 | 0.80 | (-1.67,1.48) |  |
| Race/ethnicity (ref: plurality) | Race/ethnicity minority | -0.04 | 0.11 | (-0.26,0.19) | 0.746 |

***Supplementary Table S13C. E-Values for Estimates and CI for Nigeria***

| **Variable** | **Category** | **E-Value for Estimate** | **E-Value for 95% CI** |
| --- | --- | --- | --- |
| Relationship with mother (ref: Very bad/Somewhat bad) | Very good/Somewhat good | 1.22 | 1.00 |
| Relationship with father (ref: Very bad/Somewhat bad) | Very good/Somewhat good | 1.31 | 1.00 |
| Parent marital status (ref: Married) | Divorced | 1.07 | 1.00 |
|  | Single, never married | 1.62 | 1.11 |
|  | One or both parents had died | 1.79 | 1.36 |
| Subjective financial status of family growing up (ref: Got by) | Lived comfortably | 1.09 | 1.00 |
|  | Found it difficult | 1.28 | 1.00 |
|  | Found it very difficult | 1.32 | 1.00 |
| Abuse (ref: No) | Yes | 1.51 | 1.14 |
| Felt like an outsider in the family (ref: No) | Yes | 1.15 | 1.00 |
| Self-rated health growing up (ref: Good) | Excellent | 1.23 | 1.00 |
|  | Very good | 1.14 | 1.00 |
|  | Fair | 1.48 | 1.00 |
|  | Poor | 1.12 | 1.00 |
| Immigration status (ref: Born in this country) | Born in another country | 2.11 | 1.15 |
| Age 12 religious service attendance (ref: Never) | At least 1/week | 1.22 | 1.00 |
|  | 1-3/month | 1.28 | 1.00 |
|  | <1/month | 2.12 | 1.31 |
| Birth year (ref: 1998-2005; current age: 18-24) | 1993-1998; age 25-29 | 1.18 | 1.00 |
|  | 1983-1993; age 30-39 | 1.19 | 1.00 |
|  | 1973-1983; age 40-49 | 1.15 | 1.00 |
|  | 1963-1973; age 50-59 | 1.62 | 1.19 |
|  | 1953-1963; age 60-69 | 1.36 | 1.00 |
|  | 1943-1953; age 70-79 | 1.55 | 1.00 |
|  | 1943 or earlier; age 80+ | 4.42 | 1.16 |
| Gender (ref:Male) | Female | 1.06 | 1.00 |
|  | Other | 1.37 | 1.00 |
| Religious affiliation at age 12 (ref: Christianity) | Islam | 1.53 | 1.25 |
|  | Collapsed affiliations with prevalence<3% | 1.25 | 1.00 |
| Race/ethnicity (ref: plurality) | Race/ethnicity minority | 1.14 | 1.00 |

***Supplementary Table S14A. Nationally Representative Childhood Descriptive Statistics for Philippines***

| **Variable** | **Category** | **N (%)** |
| --- | --- | --- |
| Relationship with mother | Very good | 3333 (63%) |
|  | Somewhat good | 1703 (32%) |
|  | Somewhat bad | 124 (2%) |
|  | Very bad | 39 (1%) |
|  | Does not apply | 59 (1%) |
|  | Missing | 35 (1%) |
| Relationships with father | Very good | 3443 (65%) |
|  | Somewhat good | 1429 (27%) |
|  | Somewhat bad | 159 (3%) |
|  | Very bad | 58 (1%) |
|  | Does not apply | 108 (2%) |
|  | Missing | 95 (2%) |
| Parent marital status | Parents married | 4575 (86%) |
|  | Divorced | 64 (1%) |
|  | Single, never married | 517 (10%) |
|  | One or both parents had died | 51 (1%) |
|  | Missing | 85 (2%) |
| Subjective financial status of family growing up | Lived comfortably | 937 (18%) |
|  | Got by | 3006 (57%) |
|  | Found it difficult | 1055 (20%) |
|  | Found it very difficult | 291 (6%) |
|  | Missing | 3 (0%) |
| Abuse | Yes | 420 (8%) |
|  | No | 4837 (91%) |
|  | Missing | 35 (1%) |
| Felt like an outsider in the family | Yes | 395 (7%) |
|  | No | 4884 (92%) |
|  | Missing | 13 (0%) |
| Self-rated health growing up | Excellent | 1041 (20%) |
|  | Very good | 559 (11%) |
|  | Good | 2174 (41%) |
|  | Fair | 1246 (24%) |
|  | Poor | 272 (5%) |
|  | Missing | 0 (0%) |
| Immigration status | Born in this country | 5284 (100%) |
|  | Born in another country | 8 (0%) |
| Age 12 religious service attendance | At least 1/week | 2453 (46%) |
|  | 1-3/month | 1699 (32%) |
|  | <1/ month | 892 (17%) |
|  | Never | 201 (4%) |
|  | Missing | 47 (1%) |
| Year of birth | 1998-2005; current age: 18-24 | 1073 (20%) |
|  | 1993-1998; age 25-29 | 695 (13%) |
|  | 1983-1993; age 30-39 | 1160 (22%) |
|  | 1973-1983; age 40-49 | 972 (18%) |
|  | 1963-1973; age 50-59 | 732 (14%) |
|  | 1953-1963; age 60-69 | 495 (9%) |
|  | 1943-1953; age 70-79 | 143 (3%) |
|  | 1943 or earlier; age 80+ | 23 (0%) |
| Gender | Male | 2625 (50%) |
|  | Female | 2643 (50%) |
|  | Other | 13 (0%) |
|  | Missing | 11 (0%) |
| Religious affiliation at age 12 | Christianity | 4968 (94%) |
|  | Islam | 276 (5%) |
|  | Buddhism | 1 (0%) |
|  | Sikhism | 4 (0%) |
|  | Baha’i | 1 (0%) |
|  | Primal, Animist, or Folk religion | 14 (0%) |
|  | Some other religion | 9 (0%) |
|  | No religion/Atheist/Agnostic | 9 (0%) |
|  | Missing | 11 (0%) |
| Race/ethnicity | Tagalog | 1691 (32%) |
|  | Cebuano | 656 (12%) |
|  | Ilocano/Ilokano | 429 (8%) |
|  | Visayan/Bisaya | 739 (14%) |
|  | Ilonggo/Hiligaynon | 428 (8%) |
|  | Bicolano/Bikolano | 300 (6%) |
|  | Waray | 216 (4%) |
|  | Tausug | 94 (2%) |
|  | Maranao | 39 (1%) |
|  | Maguindanaoan | 84 (2%) |
|  | Chinese-Filipino | 3 (0%) |
|  | Kapampangan | 107 (2%) |
|  | Pangasinense | 107 (2%) |
|  | Zamboangueno | 51 (1%) |
|  | Masbateno | 54 (1%) |
|  | Aeta | 1 (0%) |
|  | Igorot | 42 (1%) |
|  | Mangyan | 2 (0%) |
|  | Badjao | 2 (0%) |
|  | Other | 244 (5%) |
|  | Missing | 3 (0%) |

***Supplementary Table S14B. Regression of Promoting Good on Childhood Predictors for Philippines***

| **Variable** | **Category** | **Estimate** | **SE** | **95% CI** | **Global P-value** |
| --- | --- | --- | --- | --- | --- |
| Relationship with mother (ref: Very bad/Somewhat bad) | Very good/Somewhat good | 0.20 | 0.19 | (-0.17,0.56) | 0.266 |
| Relationship with father (ref: Very bad/Somewhat bad) | Very good/Somewhat good | -0.09 | 0.15 | (-0.40,0.21) | 0.532 |
| Parent marital status (ref: Married) | Divorced | -0.08 | 0.30 | (-0.67,0.52) | 0.035 |
|  | Single, never married | 0.17 | 0.09 | (-0.01,0.35) |  |
|  | One or both parents had died | 0.55 | 0.25 | (0.06,1.04) |  |
| Subjective financial status of family growing up (ref: Got by) | Lived comfortably | 0.30 | 0.08 | (0.14,0.46) | 0.003 |
|  | Found it difficult | 0.03 | 0.09 | (-0.15,0.20) |  |
|  | Found it very difficult | 0.07 | 0.22 | (-0.35,0.49) |  |
| Abuse (ref: No) | Yes | -0.18 | 0.15 | (-0.47,0.11) | 0.227 |
| Felt like an outsider in the family (ref: No) | Yes | -0.26 | 0.15 | (-0.55,0.03) | 0.081 |
| Self-rated health growing up (ref: Good) | Excellent | 0.11 | 0.09 | (-0.06,0.29) | 0.006 |
|  | Very good | -0.05 | 0.12 | (-0.29,0.19) |  |
|  | Fair | -0.16 | 0.08 | (-0.32,0.00) |  |
|  | Poor | -0.41 | 0.20 | (-0.80,-0.02) |  |
| Immigration status (ref: Born in this country) | Born in another country | -0.82 | 0.74 | (-2.27,0.64) | 0.270 |
| Age 12 religious service attendance (ref: Never) | At least 1/week | 0.49 | 0.25 | (-0.01,0.98) | 0.221 |
|  | 1-3/month | 0.50 | 0.26 | (-0.02,1.01) |  |
|  | <1/month | 0.42 | 0.27 | (-0.10,0.94) |  |
| Birth year (ref: 1998-2005; current age: 18-24) | 1993-1998; age 25-29 | 0.36 | 0.11 | (0.14,0.58) | 0.012 |
|  | 1983-1993; age 30-39 | 0.30 | 0.09 | (0.12,0.49) |  |
|  | 1973-1983; age 40-49 | 0.15 | 0.11 | (-0.07,0.37) |  |
|  | 1963-1973; age 50-59 | 0.15 | 0.14 | (-0.11,0.42) |  |
|  | 1953-1963; age 60-69 | 0.24 | 0.16 | (-0.08,0.55) |  |
|  | 1943-1953; age 70-79 | 0.15 | 0.23 | (-0.31,0.61) |  |
|  | 1943 or earlier; age 80+ | -0.67 | 0.67 | (-1.98,0.64) |  |
| Gender (ref:Male) | Female | 0.06 | 0.07 | (-0.07,0.19) | 0.070 |
|  | Other | -0.83 | 0.41 | (-1.63,-0.02) |  |
| Religious affiliation at age 12 (ref: Christianity) | Islam | 0.00 | 0.19 | (-0.36,0.36) | 0.986 |
|  | Collapsed affiliations with prevalence<3% | -0.06 | 0.38 | (-0.80,0.67) |  |
| Race/ethnicity (ref: plurality) | Race/ethnicity minority | -0.07 | 0.07 | (-0.22,0.07) | 0.315 |

***Supplementary Table S14C. E-Values for Estimates and CI for Philippines***

| **Variable** | **Category** | **E-Value for Estimate** | **E-Value for 95% CI** |
| --- | --- | --- | --- |
| Relationship with mother (ref: Very bad/Somewhat bad) | Very good/Somewhat good | 1.42 | 1.00 |
| Relationship with father (ref: Very bad/Somewhat bad) | Very good/Somewhat good | 1.26 | 1.00 |
| Parent marital status (ref: Married) | Divorced | 1.23 | 1.00 |
|  | Single, never married | 1.38 | 1.00 |
|  | One or both parents had died | 1.90 | 1.20 |
| Subjective financial status of family growing up (ref: Got by) | Lived comfortably | 1.56 | 1.33 |
|  | Found it difficult | 1.13 | 1.00 |
|  | Found it very difficult | 1.22 | 1.00 |
| Abuse (ref: No) | Yes | 1.39 | 1.00 |
| Felt like an outsider in the family (ref: No) | Yes | 1.51 | 1.00 |
| Self-rated health growing up (ref: Good) | Excellent | 1.29 | 1.00 |
|  | Very good | 1.17 | 1.00 |
|  | Fair | 1.36 | 1.00 |
|  | Poor | 1.71 | 1.11 |
| Immigration status (ref: Born in this country) | Born in another country | 2.28 | 1.00 |
| Age 12 religious service attendance (ref: Never) | At least 1/week | 1.81 | 1.00 |
|  | 1-3/month | 1.83 | 1.00 |
|  | <1/month | 1.72 | 1.00 |
| Birth year (ref: 1998-2005; current age: 18-24) | 1993-1998; age 25-29 | 1.64 | 1.33 |
|  | 1983-1993; age 30-39 | 1.57 | 1.30 |
|  | 1973-1983; age 40-49 | 1.35 | 1.00 |
|  | 1963-1973; age 50-59 | 1.35 | 1.00 |
|  | 1953-1963; age 60-69 | 1.47 | 1.00 |
|  | 1943-1953; age 70-79 | 1.35 | 1.00 |
|  | 1943 or earlier; age 80+ | 2.07 | 1.00 |
| Gender (ref:Male) | Female | 1.19 | 1.00 |
|  | Other | 2.29 | 1.11 |
| Religious affiliation at age 12 (ref: Christianity) | Islam | 1.01 | 1.00 |
|  | Collapsed affiliations with prevalence<3% | 1.21 | 1.00 |
| Race/ethnicity (ref: plurality) | Race/ethnicity minority | 1.23 | 1.00 |

***Supplementary Table S15A. Nationally Representative Childhood Descriptive Statistics for Poland***

| **Variable** | **Category** | **N (%)** |
| --- | --- | --- |
| Relationship with mother | Very good | 4879 (47%) |
|  | Somewhat good | 4973 (48%) |
|  | Somewhat bad | 285 (3%) |
|  | Very bad | 58 (1%) |
|  | Does not apply | 80 (1%) |
|  | Missing | 112 (1%) |
| Relationships with father | Very good | 4231 (41%) |
|  | Somewhat good | 4984 (48%) |
|  | Somewhat bad | 516 (5%) |
|  | Very bad | 78 (1%) |
|  | Does not apply | 407 (4%) |
|  | Missing | 173 (2%) |
| Parent marital status | Parents married | 8972 (86%) |
|  | Divorced | 587 (6%) |
|  | Single, never married | 193 (2%) |
|  | One or both parents had died | 313 (3%) |
|  | Missing | 324 (3%) |
| Subjective financial status of family growing up | Lived comfortably | 1384 (13%) |
|  | Got by | 6257 (60%) |
|  | Found it difficult | 2133 (21%) |
|  | Found it very difficult | 509 (5%) |
|  | Missing | 106 (1%) |
| Abuse | Yes | 325 (3%) |
|  | No | 10009 (96%) |
|  | Missing | 55 (1%) |
| Felt like an outsider in the family | Yes | 490 (5%) |
|  | No | 9615 (93%) |
|  | Missing | 284 (3%) |
| Self-rated health growing up | Excellent | 2676 (26%) |
|  | Very good | 5371 (52%) |
|  | Good | 1779 (17%) |
|  | Fair | 406 (4%) |
|  | Poor | 123 (1%) |
|  | Missing | 34 (0%) |
| Immigration status | Born in this country | 10258 (99%) |
|  | Born in another country | 108 (1%) |
|  | Missing | 23 (0%) |
| Age 12 religious service attendance | At least 1/week | 4751 (46%) |
|  | 1-3/month | 2689 (26%) |
|  | <1/ month | 2161 (21%) |
|  | Never | 354 (3%) |
|  | Missing | 434 (4%) |
| Year of birth | 1998-2005; current age: 18-24 | 955 (9%) |
|  | 1993-1998; age 25-29 | 761 (7%) |
|  | 1983-1993; age 30-39 | 2159 (21%) |
|  | 1973-1983; age 40-49 | 1956 (19%) |
|  | 1963-1973; age 50-59 | 1670 (16%) |
|  | 1953-1963; age 60-69 | 1909 (18%) |
|  | 1943-1953; age 70-79 | 833 (8%) |
|  | 1943 or earlier; age 80+ | 145 (1%) |
|  | Missing | 1 (0%) |
| Gender | Male | 4974 (48%) |
|  | Female | 5387 (52%) |
|  | Other | 3 (0%) |
|  | Missing | 26 (0%) |
| Religious affiliation at age 12 | Christianity | 9861 (95%) |
|  | Islam | 3 (0%) |
|  | Buddhism | 2 (0%) |
|  | Sikhism | 1 (0%) |
|  | Primal, Animist, or Folk religion | 5 (0%) |
|  | No religion/Atheist/Agnostic | 482 (5%) |
|  | Missing | 35 (0%) |
| Race/ethnicity | Polish | 10309 (99%) |
|  | German | 4 (0%) |
|  | Belarussian | 2 (0%) |
|  | Ukrainian | 38 (0%) |
|  | Silesia | 14 (0%) |
|  | Kashubians | 3 (0%) |
|  | Other | 4 (0%) |
|  | Missing | 14 (0%) |

***Supplementary Table S15B. Regression of Promoting Good on Childhood Predictors for Poland***

| **Variable** | **Category** | **Estimate** | **SE** | **95% CI** | **Global P-value** |
| --- | --- | --- | --- | --- | --- |
| Relationship with mother (ref: Very bad/Somewhat bad) | Very good/Somewhat good | 0.07 | 0.19 | (-0.31,0.44) | 0.635 |
| Relationship with father (ref: Very bad/Somewhat bad) | Very good/Somewhat good | 0.25 | 0.14 | (-0.03,0.53) | 0.082 |
| Parent marital status (ref: Married) | Divorced | -0.31 | 0.10 | (-0.51,-0.11) | <.001 |
|  | Single, never married | -0.59 | 0.20 | (-0.99,-0.19) |  |
|  | One or both parents had died | -0.24 | 0.25 | (-0.74,0.26) |  |
| Subjective financial status of family growing up (ref: Got by) | Lived comfortably | 0.03 | 0.08 | (-0.13,0.18) | 0.343 |
|  | Found it difficult | 0.04 | 0.06 | (-0.08,0.16) |  |
|  | Found it very difficult | -0.27 | 0.18 | (-0.63,0.08) |  |
| Abuse (ref: No) | Yes | -0.24 | 0.16 | (-0.55,0.07) | 0.128 |
| Felt like an outsider in the family (ref: No) | Yes | -0.36 | 0.16 | (-0.67,-0.05) | 0.021 |
| Self-rated health growing up (ref: Good) | Excellent | 0.48 | 0.10 | (0.29,0.67) | <.001 |
|  | Very good | 0.13 | 0.08 | (-0.03,0.29) |  |
|  | Fair | -0.17 | 0.18 | (-0.53,0.19) |  |
|  | Poor | 0.07 | 0.25 | (-0.42,0.57) |  |
| Immigration status (ref: Born in this country) | Born in another country | -0.58 | 0.32 | (-1.21,0.05) | 0.069 |
| Age 12 religious service attendance (ref: Never) | At least 1/week | 1.01 | 0.20 | (0.62,1.40) | <.001 |
|  | 1-3/month | 0.65 | 0.17 | (0.31,1.00) |  |
|  | <1/month | 0.40 | 0.19 | (0.02,0.78) |  |
| Birth year (ref: 1998-2005; current age: 18-24) | 1993-1998; age 25-29 | -0.01 | 0.10 | (-0.21,0.19) | 0.040 |
|  | 1983-1993; age 30-39 | 0.10 | 0.10 | (-0.08,0.29) |  |
|  | 1973-1983; age 40-49 | -0.02 | 0.10 | (-0.22,0.17) |  |
|  | 1963-1973; age 50-59 | -0.01 | 0.10 | (-0.21,0.20) |  |
|  | 1953-1963; age 60-69 | 0.23 | 0.11 | (0.02,0.44) |  |
|  | 1943-1953; age 70-79 | 0.26 | 0.17 | (-0.07,0.59) |  |
|  | 1943 or earlier; age 80+ | 0.21 | 0.27 | (-0.32,0.74) |  |
| Gender (ref:Male) | Female | 0.20 | 0.05 | (0.10,0.29) | <.001 |
|  | Other | -1.12 | 0.41 | (-1.92,-0.31) |  |
| Religious affiliation at age 12 (ref: No religion/Atheist/Agnostic) | Christianity | -0.48 | 0.19 | (-0.85,-0.11) | <.001 |
|  | Collapsed affiliations with prevalence<3% | -1.90 | 1.04 | (-4.24,0.43) |  |
| Race/ethnicity (ref: plurality) | Race/ethnicity minority | 0.42 | 0.30 | (-0.16,1.01) | 0.153 |

***Supplementary Table S15C. E-Values for Estimates and CI for Poland***

| **Variable** | **Category** | **E-Value for Estimate** | **E-Value for 95% CI** |
| --- | --- | --- | --- |
| Relationship with mother (ref: Very bad/Somewhat bad) | Very good/Somewhat good | 1.23 | 1.00 |
| Relationship with father (ref: Very bad/Somewhat bad) | Very good/Somewhat good | 1.56 | 1.00 |
| Parent marital status (ref: Married) | Divorced | 1.66 | 1.33 |
|  | Single, never married | 2.12 | 1.47 |
|  | One or both parents had died | 1.55 | 1.00 |
| Subjective financial status of family growing up (ref: Got by) | Lived comfortably | 1.14 | 1.00 |
|  | Found it difficult | 1.17 | 1.00 |
|  | Found it very difficult | 1.60 | 1.00 |
| Abuse (ref: No) | Yes | 1.54 | 1.00 |
| Felt like an outsider in the family (ref: No) | Yes | 1.73 | 1.19 |
| Self-rated health growing up (ref: Good) | Excellent | 1.94 | 1.63 |
|  | Very good | 1.36 | 1.00 |
|  | Fair | 1.42 | 1.00 |
|  | Poor | 1.25 | 1.00 |
| Immigration status (ref: Born in this country) | Born in another country | 2.10 | 1.00 |
| Age 12 religious service attendance (ref: Never) | At least 1/week | 2.89 | 2.17 |
|  | 1-3/month | 2.23 | 1.67 |
|  | <1/month | 1.81 | 1.13 |
| Birth year (ref: 1998-2005; current age: 18-24) | 1993-1998; age 25-29 | 1.08 | 1.00 |
|  | 1983-1993; age 30-39 | 1.31 | 1.00 |
|  | 1973-1983; age 40-49 | 1.13 | 1.00 |
|  | 1963-1973; age 50-59 | 1.06 | 1.00 |
|  | 1953-1963; age 60-69 | 1.53 | 1.11 |
|  | 1943-1953; age 70-79 | 1.58 | 1.00 |
|  | 1943 or earlier; age 80+ | 1.49 | 1.00 |
| Gender (ref:Male) | Female | 1.47 | 1.31 |
|  | Other | 3.11 | 1.67 |
| Religious affiliation at age 12 (ref: No religion/Atheist/Agnostic) | Christianity | 1.93 | 1.32 |
|  | Collapsed affiliations with prevalence<3% | 5.19 | 1.00 |
| Race/ethnicity (ref: plurality) | Race/ethnicity minority | 1.84 | 1.00 |

***Supplementary Table S16A. Nationally Representative Childhood Descriptive Statistics for South Africa***

| **Variable** | **Category** | **N (%)** |
| --- | --- | --- |
| Relationship with mother | Very good | 2186 (82%) |
|  | Somewhat good | 263 (10%) |
|  | Somewhat bad | 51 (2%) |
|  | Very bad | 39 (1%) |
|  | Does not apply | 90 (3%) |
|  | Missing | 21 (1%) |
| Relationships with father | Very good | 1656 (62%) |
|  | Somewhat good | 333 (13%) |
|  | Somewhat bad | 86 (3%) |
|  | Very bad | 159 (6%) |
|  | Does not apply | 331 (12%) |
|  | Missing | 85 (3%) |
| Parent marital status | Parents married | 1321 (50%) |
|  | Divorced | 131 (5%) |
|  | Single, never married | 904 (34%) |
|  | One or both parents had died | 140 (5%) |
|  | Missing | 155 (6%) |
| Subjective financial status of family growing up | Lived comfortably | 1050 (40%) |
|  | Got by | 875 (33%) |
|  | Found it difficult | 432 (16%) |
|  | Found it very difficult | 289 (11%) |
|  | Missing | 5 (0%) |
| Abuse | Yes | 450 (17%) |
|  | No | 2149 (81%) |
|  | Missing | 52 (2%) |
| Felt like an outsider in the family | Yes | 434 (16%) |
|  | No | 2211 (83%) |
|  | Missing | 6 (0%) |
| Self-rated health growing up | Excellent | 1225 (46%) |
|  | Very good | 590 (22%) |
|  | Good | 370 (14%) |
|  | Fair | 266 (10%) |
|  | Poor | 183 (7%) |
|  | Missing | 17 (1%) |
| Immigration status | Born in this country | 2511 (95%) |
|  | Born in another country | 139 (5%) |
|  | Missing | 1 (0%) |
| Age 12 religious service attendance | At least 1/week | 1681 (63%) |
|  | 1-3/month | 552 (21%) |
|  | <1/ month | 175 (7%) |
|  | Never | 217 (8%) |
|  | Missing | 26 (1%) |
| Year of birth | 1998-2005; current age: 18-24 | 461 (17%) |
|  | 1993-1998; age 25-29 | 364 (14%) |
|  | 1983-1993; age 30-39 | 655 (25%) |
|  | 1973-1983; age 40-49 | 522 (20%) |
|  | 1963-1973; age 50-59 | 309 (12%) |
|  | 1953-1963; age 60-69 | 195 (7%) |
|  | 1943-1953; age 70-79 | 120 (5%) |
|  | 1943 or earlier; age 80+ | 17 (1%) |
|  | Missing | 9 (0%) |
| Gender | Male | 1288 (49%) |
|  | Female | 1356 (51%) |
|  | Other | 2 (0%) |
|  | Missing | 4 (0%) |
| Religious affiliation at age 12 | Christianity | 2323 (88%) |
|  | Islam | 52 (2%) |
|  | Hinduism | 2 (0%) |
|  | Buddhism | 11 (0%) |
|  | Shinto | 2 (0%) |
|  | Taoism | 1 (0%) |
|  | Primal, Animist, or Folk religion | 117 (4%) |
|  | Some other religion | 7 (0%) |
|  | No religion/Atheist/Agnostic | 107 (4%) |
|  | Missing | 27 (1%) |
| Race/ethnicity | Black | 2381 (90%) |
|  | Asian/Indian | 6 (0%) |
|  | Colored | 252 (10%) |
|  | White | 8 (0%) |
|  | Other | 1 (0%) |
|  | Missing | 3 (0%) |

***Supplementary Table S16B. Regression of Promoting Good on Childhood Predictors for South Africa***

| **Variable** | **Category** | **Estimate** | **SE** | **95% CI** | **Global P-value** |
| --- | --- | --- | --- | --- | --- |
| Relationship with mother (ref: Very bad/Somewhat bad) | Very good/Somewhat good | 0.15 | 0.35 | (-0.54,0.85) | 0.659 |
| Relationship with father (ref: Very bad/Somewhat bad) | Very good/Somewhat good | -0.26 | 0.17 | (-0.60,0.07) | 0.115 |
| Parent marital status (ref: Married) | Divorced | 0.39 | 0.20 | (-0.01,0.79) | 0.140 |
|  | Single, never married | -0.12 | 0.13 | (-0.38,0.15) |  |
|  | One or both parents had died | -0.10 | 0.32 | (-0.73,0.52) |  |
| Subjective financial status of family growing up (ref: Got by) | Lived comfortably | -0.01 | 0.16 | (-0.32,0.31) | 0.906 |
|  | Found it difficult | 0.07 | 0.18 | (-0.29,0.42) |  |
|  | Found it very difficult | -0.11 | 0.25 | (-0.59,0.38) |  |
| Abuse (ref: No) | Yes | -0.53 | 0.17 | (-0.86,-0.19) | 0.002 |
| Felt like an outsider in the family (ref: No) | Yes | 0.16 | 0.17 | (-0.17,0.49) | 0.335 |
| Self-rated health growing up (ref: Good) | Excellent | 0.16 | 0.18 | (-0.19,0.51) | 0.260 |
|  | Very good | 0.40 | 0.19 | (0.03,0.77) |  |
|  | Fair | 0.04 | 0.24 | (-0.44,0.52) |  |
|  | Poor | 0.19 | 0.31 | (-0.41,0.80) |  |
| Immigration status (ref: Born in this country) | Born in another country | -0.38 | 0.30 | (-0.97,0.21) | 0.203 |
| Age 12 religious service attendance (ref: Never) | At least 1/week | -0.01 | 0.32 | (-0.64,0.62) | 0.949 |
|  | 1-3/month | 0.01 | 0.32 | (-0.62,0.64) |  |
|  | <1/month | -0.14 | 0.38 | (-0.88,0.61) |  |
| Birth year (ref: 1998-2005; current age: 18-24) | 1993-1998; age 25-29 | -0.35 | 0.20 | (-0.75,0.04) | 0.002 |
|  | 1983-1993; age 30-39 | 0.19 | 0.18 | (-0.16,0.55) |  |
|  | 1973-1983; age 40-49 | -0.18 | 0.19 | (-0.55,0.19) |  |
|  | 1963-1973; age 50-59 | 0.11 | 0.23 | (-0.34,0.56) |  |
|  | 1953-1963; age 60-69 | -0.85 | 0.41 | (-1.66,-0.03) |  |
|  | 1943-1953; age 70-79 | 0.28 | 0.38 | (-0.47,1.02) |  |
|  | 1943 or earlier; age 80+ | 1.02 | 0.54 | (-0.06,2.09) |  |
| Gender (ref:Male) | Female | -0.06 | 0.13 | (-0.31,0.19) | 0.050 |
|  | Other | -0.75 | 0.31 | (-1.36,-0.15) |  |
| Religious affiliation at age 12 (ref: No religion/Atheist/Agnostic) | Christianity | 0.06 | 0.43 | (-0.79,0.91) | 0.963 |
|  | Primal, Animist, or Folk religion | 0.07 | 0.50 | (-0.91,1.04) |  |
|  | Collapsed affiliations with prevalence<3% | 0.22 | 0.55 | (-0.86,1.29) |  |
| Race/ethnicity (ref: plurality) | Race/ethnicity minority | 0.44 | 0.25 | (-0.05,0.92) | 0.078 |

***Supplementary Table S16C. E-Values for Estimates and CI for South Africa***

| **Variable** | **Category** | **E-Value for Estimate** | **E-Value for 95% CI** |
| --- | --- | --- | --- |
| Relationship with mother (ref: Very bad/Somewhat bad) | Very good/Somewhat good | 1.32 | 1.00 |
| Relationship with father (ref: Very bad/Somewhat bad) | Very good/Somewhat good | 1.45 | 1.00 |
| Parent marital status (ref: Married) | Divorced | 1.60 | 1.00 |
|  | Single, never married | 1.26 | 1.00 |
|  | One or both parents had died | 1.25 | 1.00 |
| Subjective financial status of family growing up (ref: Got by) | Lived comfortably | 1.05 | 1.00 |
|  | Found it difficult | 1.19 | 1.00 |
|  | Found it very difficult | 1.25 | 1.00 |
| Abuse (ref: No) | Yes | 1.75 | 1.37 |
| Felt like an outsider in the family (ref: No) | Yes | 1.33 | 1.00 |
| Self-rated health growing up (ref: Good) | Excellent | 1.32 | 1.00 |
|  | Very good | 1.61 | 1.13 |
|  | Fair | 1.14 | 1.00 |
|  | Poor | 1.37 | 1.00 |
| Immigration status (ref: Born in this country) | Born in another country | 1.58 | 1.00 |
| Age 12 religious service attendance (ref: Never) | At least 1/week | 1.05 | 1.00 |
|  | 1-3/month | 1.06 | 1.00 |
|  | <1/month | 1.30 | 1.00 |
| Birth year (ref: 1998-2005; current age: 18-24) | 1993-1998; age 25-29 | 1.56 | 1.00 |
|  | 1983-1993; age 30-39 | 1.36 | 1.00 |
|  | 1973-1983; age 40-49 | 1.35 | 1.00 |
|  | 1963-1973; age 50-59 | 1.25 | 1.00 |
|  | 1953-1963; age 60-69 | 2.12 | 1.14 |
|  | 1943-1953; age 70-79 | 1.47 | 1.00 |
|  | 1943 or earlier; age 80+ | 2.32 | 1.00 |
| Gender (ref:Male) | Female | 1.17 | 1.00 |
|  | Other | 2.00 | 1.31 |
| Religious affiliation at age 12 (ref: No religion/Atheist/Agnostic) | Christianity | 1.18 | 1.00 |
|  | Primal, Animist, or Folk religion | 1.19 | 1.00 |
|  | Collapsed affiliations with prevalence<3% | 1.39 | 1.00 |
| Race/ethnicity (ref: plurality) | Race/ethnicity minority | 1.65 | 1.00 |

***Supplementary Table S17A. Nationally Representative Childhood Descriptive Statistics for Spain***

| **Variable** | **Category** | **N (%)** |
| --- | --- | --- |
| Relationship with mother | Very good | 4557 (72%) |
|  | Somewhat good | 1258 (20%) |
|  | Somewhat bad | 248 (4%) |
|  | Very bad | 92 (1%) |
|  | Does not apply | 107 (2%) |
|  | Missing | 28 (0%) |
| Relationships with father | Very good | 4131 (66%) |
|  | Somewhat good | 1397 (22%) |
|  | Somewhat bad | 309 (5%) |
|  | Very bad | 178 (3%) |
|  | Does not apply | 243 (4%) |
|  | Missing | 33 (1%) |
| Parent marital status | Parents married | 5285 (84%) |
|  | Divorced | 378 (6%) |
|  | Single, never married | 312 (5%) |
|  | One or both parents had died | 126 (2%) |
|  | Missing | 188 (3%) |
| Subjective financial status of family growing up | Lived comfortably | 2041 (32%) |
|  | Got by | 2956 (47%) |
|  | Found it difficult | 1154 (18%) |
|  | Found it very difficult | 110 (2%) |
|  | Missing | 29 (0%) |
| Abuse | Yes | 659 (10%) |
|  | No | 5510 (88%) |
|  | Missing | 122 (2%) |
| Felt like an outsider in the family | Yes | 579 (9%) |
|  | No | 5637 (90%) |
|  | Missing | 75 (1%) |
| Self-rated health growing up | Excellent | 2450 (39%) |
|  | Very good | 2286 (36%) |
|  | Good | 1235 (20%) |
|  | Fair | 164 (3%) |
|  | Poor | 135 (2%) |
|  | Missing | 20 (0%) |
| Immigration status | Born in this country | 5479 (87%) |
|  | Born in another country | 788 (13%) |
|  | Missing | 23 (0%) |
| Age 12 religious service attendance | At least 1/week | 2391 (38%) |
|  | 1-3/month | 1132 (18%) |
|  | <1/ month | 1287 (20%) |
|  | Never | 1445 (23%) |
|  | Missing | 36 (1%) |
| Year of birth | 1998-2005; current age: 18-24 | 594 (9%) |
|  | 1993-1998; age 25-29 | 450 (7%) |
|  | 1983-1993; age 30-39 | 1111 (18%) |
|  | 1973-1983; age 40-49 | 1396 (22%) |
|  | 1963-1973; age 50-59 | 1252 (20%) |
|  | 1953-1963; age 60-69 | 977 (16%) |
|  | 1943-1953; age 70-79 | 467 (7%) |
|  | 1943 or earlier; age 80+ | 43 (1%) |
| Gender | Male | 3142 (50%) |
|  | Female | 3119 (50%) |
|  | Other | 6 (0%) |
|  | Missing | 22 (0%) |
| Religious affiliation at age 12 | Christianity | 5119 (81%) |
|  | Islam | 132 (2%) |
|  | Hinduism | 5 (0%) |
|  | Buddhism | 8 (0%) |
|  | Judaism | 5 (0%) |
|  | Sikhism | 2 (0%) |
|  | Confucianism | 1 (0%) |
|  | Primal, Animist, or Folk religion | 4 (0%) |
|  | Some other religion | 13 (0%) |
|  | No religion/Atheist/Agnostic | 972 (15%) |
|  | Missing | 29 (0%) |

***Supplementary Table S17B. Regression of Promoting Good on Childhood Predictors for Spain***

| **Variable** | **Category** | **Estimate** | **SE** | **95% CI** | **Global P-value** |
| --- | --- | --- | --- | --- | --- |
| Relationship with mother (ref: Very bad/Somewhat bad) | Very good/Somewhat good | 0.48 | 0.14 | (0.20,0.75) | <.001 |
| Relationship with father (ref: Very bad/Somewhat bad) | Very good/Somewhat good | 0.17 | 0.10 | (-0.03,0.36) | 0.097 |
| Parent marital status (ref: Married) | Divorced | -0.10 | 0.12 | (-0.33,0.12) | 0.374 |
|  | Single, never married | -0.20 | 0.13 | (-0.45,0.06) |  |
|  | One or both parents had died | 0.02 | 0.19 | (-0.34,0.39) |  |
| Subjective financial status of family growing up (ref: Got by) | Lived comfortably | 0.12 | 0.06 | (0.00,0.24) | 0.022 |
|  | Found it difficult | 0.22 | 0.08 | (0.07,0.37) |  |
|  | Found it very difficult | -0.03 | 0.28 | (-0.58,0.52) |  |
| Abuse (ref: No) | Yes | -0.03 | 0.09 | (-0.21,0.16) | 0.751 |
| Felt like an outsider in the family (ref: No) | Yes | -0.07 | 0.10 | (-0.27,0.12) | 0.410 |
| Self-rated health growing up (ref: Good) | Excellent | 0.51 | 0.08 | (0.35,0.67) | <.001 |
|  | Very good | 0.26 | 0.08 | (0.10,0.41) |  |
|  | Fair | 0.35 | 0.18 | (-0.00,0.71) |  |
|  | Poor | 0.36 | 0.24 | (-0.12,0.83) |  |
| Immigration status (ref: Born in this country) | Born in another country | 0.55 | 0.07 | (0.42,0.69) | <.001 |
| Age 12 religious service attendance (ref: Never) | At least 1/week | 0.30 | 0.08 | (0.15,0.45) | <.001 |
|  | 1-3/month | 0.13 | 0.08 | (-0.02,0.29) |  |
|  | <1/month | -0.15 | 0.09 | (-0.32,0.02) |  |
| Birth year (ref: 1998-2005; current age: 18-24) | 1993-1998; age 25-29 | 0.14 | 0.12 | (-0.10,0.39) | 0.002 |
|  | 1983-1993; age 30-39 | 0.09 | 0.10 | (-0.11,0.29) |  |
|  | 1973-1983; age 40-49 | 0.27 | 0.10 | (0.08,0.46) |  |
|  | 1963-1973; age 50-59 | 0.29 | 0.10 | (0.08,0.50) |  |
|  | 1953-1963; age 60-69 | 0.23 | 0.12 | (-0.00,0.46) |  |
|  | 1943-1953; age 70-79 | 0.54 | 0.15 | (0.24,0.84) |  |
|  | 1943 or earlier; age 80+ | 0.63 | 0.32 | (0.01,1.26) |  |
| Gender (ref:Male) | Female | 0.07 | 0.05 | (-0.03,0.18) | 0.377 |
|  | Other | 0.04 | 0.35 | (-0.64,0.73) |  |
| Religious affiliation at age 12 (ref: No religion/Atheist/Agnostic) | Christianity | 0.17 | 0.08 | (0.01,0.33) | 0.106 |
|  | Collapsed affiliations with prevalence<3% | 0.16 | 0.19 | (-0.21,0.53) |  |

***Supplementary Table S17C. E-Values for Estimates and CI for Spain***

| **Variable** | **Category** | **E-Value for Estimate** | **E-Value for 95% CI** |
| --- | --- | --- | --- |
| Relationship with mother (ref: Very bad/Somewhat bad) | Very good/Somewhat good | 1.91 | 1.48 |
| Relationship with father (ref: Very bad/Somewhat bad) | Very good/Somewhat good | 1.42 | 1.00 |
| Parent marital status (ref: Married) | Divorced | 1.30 | 1.00 |
|  | Single, never married | 1.47 | 1.00 |
|  | One or both parents had died | 1.13 | 1.00 |
| Subjective financial status of family growing up (ref: Got by) | Lived comfortably | 1.33 | 1.03 |
|  | Found it difficult | 1.50 | 1.23 |
|  | Found it very difficult | 1.15 | 1.00 |
| Abuse (ref: No) | Yes | 1.14 | 1.00 |
| Felt like an outsider in the family (ref: No) | Yes | 1.24 | 1.00 |
| Self-rated health growing up (ref: Good) | Excellent | 1.96 | 1.72 |
|  | Very good | 1.56 | 1.30 |
|  | Fair | 1.71 | 1.00 |
|  | Poor | 1.72 | 1.00 |
| Immigration status (ref: Born in this country) | Born in another country | 2.03 | 1.81 |
| Age 12 religious service attendance (ref: Never) | At least 1/week | 1.63 | 1.40 |
|  | 1-3/month | 1.36 | 1.00 |
|  | <1/month | 1.39 | 1.00 |
| Birth year (ref: 1998-2005; current age: 18-24) | 1993-1998; age 25-29 | 1.38 | 1.00 |
|  | 1983-1993; age 30-39 | 1.28 | 1.00 |
|  | 1973-1983; age 40-49 | 1.58 | 1.26 |
|  | 1963-1973; age 50-59 | 1.61 | 1.27 |
|  | 1953-1963; age 60-69 | 1.52 | 1.00 |
|  | 1943-1953; age 70-79 | 2.01 | 1.54 |
|  | 1943 or earlier; age 80+ | 2.16 | 1.08 |
| Gender (ref:Male) | Female | 1.25 | 1.00 |
|  | Other | 1.18 | 1.00 |
| Religious affiliation at age 12 (ref: No religion/Atheist/Agnostic) | Christianity | 1.42 | 1.09 |
|  | Collapsed affiliations with prevalence<3% | 1.41 | 1.00 |

***Supplementary Table S18A. Nationally Representative Childhood Descriptive Statistics for Sweden***

| **Variable** | **Category** | **N (%)** |
| --- | --- | --- |
| Relationship with mother | Very good | 8743 (58%) |
|  | Somewhat good | 4513 (30%) |
|  | Somewhat bad | 1194 (8%) |
|  | Very bad | 371 (2%) |
|  | Does not apply | 216 (1%) |
|  | Missing | 30 (0%) |
| Relationships with father | Very good | 7134 (47%) |
|  | Somewhat good | 4885 (32%) |
|  | Somewhat bad | 1588 (11%) |
|  | Very bad | 725 (5%) |
|  | Does not apply | 720 (5%) |
|  | Missing | 16 (0%) |
| Parent marital status | Parents married | 10887 (72%) |
|  | Divorced | 1927 (13%) |
|  | Single, never married | 1747 (12%) |
|  | One or both parents had died | 362 (2%) |
|  | Missing | 145 (1%) |
| Subjective financial status of family growing up | Lived comfortably | 5951 (39%) |
|  | Got by | 7717 (51%) |
|  | Found it difficult | 1238 (8%) |
|  | Found it very difficult | 140 (1%) |
|  | Missing | 22 (0%) |
| Abuse | Yes | 2288 (15%) |
|  | No | 12735 (85%) |
|  | Missing | 45 (0%) |
| Felt like an outsider in the family | Yes | 1867 (12%) |
|  | No | 13034 (86%) |
|  | Missing | 168 (1%) |
| Self-rated health growing up | Excellent | 5733 (38%) |
|  | Very good | 5124 (34%) |
|  | Good | 2669 (18%) |
|  | Fair | 1108 (7%) |
|  | Poor | 397 (3%) |
|  | Missing | 38 (0%) |
| Immigration status | Born in this country | 13922 (92%) |
|  | Born in another country | 1052 (7%) |
|  | Missing | 94 (1%) |
| Age 12 religious service attendance | At least 1/week | 955 (6%) |
|  | 1-3/month | 1362 (9%) |
|  | <1/ month | 6224 (41%) |
|  | Never | 6472 (43%) |
|  | Missing | 54 (0%) |
| Year of birth | 1998-2005; current age: 18-24 | 1515 (10%) |
|  | 1993-1998; age 25-29 | 1399 (9%) |
|  | 1983-1993; age 30-39 | 2398 (16%) |
|  | 1973-1983; age 40-49 | 2221 (15%) |
|  | 1963-1973; age 50-59 | 2493 (17%) |
|  | 1953-1963; age 60-69 | 2168 (14%) |
|  | 1943-1953; age 70-79 | 2253 (15%) |
|  | 1943 or earlier; age 80+ | 621 (4%) |
| Gender | Male | 7536 (50%) |
|  | Female | 7493 (50%) |
|  | Other | 27 (0%) |
|  | Missing | 12 (0%) |
| Religious affiliation at age 12 | Christianity | 10617 (70%) |
|  | Islam | 462 (3%) |
|  | Hinduism | 16 (0%) |
|  | Buddhism | 41 (0%) |
|  | Judaism | 51 (0%) |
|  | Sikhism | 9 (0%) |
|  | Baha’i | 3 (0%) |
|  | Shinto | 1 (0%) |
|  | Confucianism | 4 (0%) |
|  | Primal, Animist, or Folk religion | 31 (0%) |
|  | Some other religion | 69 (0%) |
|  | No religion/Atheist/Agnostic | 3738 (25%) |
|  | Missing | 26 (0%) |

***Supplementary Table S18B. Regression of Promoting Good on Childhood Predictors for Sweden***

| **Variable** | **Category** | **Estimate** | **SE** | **95% CI** | **Global P-value** |
| --- | --- | --- | --- | --- | --- |
| Relationship with mother (ref: Very bad/Somewhat bad) | Very good/Somewhat good | 0.04 | 0.06 | (-0.08,0.16) | 0.515 |
| Relationship with father (ref: Very bad/Somewhat bad) | Very good/Somewhat good | 0.10 | 0.05 | (-0.01,0.20) | 0.061 |
| Parent marital status (ref: Married) | Divorced | 0.14 | 0.06 | (0.03,0.25) | 0.082 |
|  | Single, never married | 0.00 | 0.06 | (-0.11,0.12) |  |
|  | One or both parents had died | 0.06 | 0.12 | (-0.17,0.29) |  |
| Subjective financial status of family growing up (ref: Got by) | Lived comfortably | 0.10 | 0.04 | (0.03,0.17) | 0.032 |
|  | Found it difficult | 0.05 | 0.07 | (-0.08,0.18) |  |
|  | Found it very difficult | 0.22 | 0.24 | (-0.24,0.69) |  |
| Abuse (ref: No) | Yes | 0.13 | 0.05 | (0.03,0.23) | 0.012 |
| Felt like an outsider in the family (ref: No) | Yes | -0.09 | 0.06 | (-0.21,0.04) | 0.153 |
| Self-rated health growing up (ref: Good) | Excellent | 0.78 | 0.05 | (0.68,0.88) | <.001 |
|  | Very good | 0.29 | 0.05 | (0.20,0.39) |  |
|  | Fair | 0.07 | 0.08 | (-0.09,0.22) |  |
|  | Poor | -0.15 | 0.14 | (-0.42,0.12) |  |
| Immigration status (ref: Born in this country) | Born in another country | 0.17 | 0.07 | (0.04,0.31) | 0.013 |
| Age 12 religious service attendance (ref: Never) | At least 1/week | 0.42 | 0.07 | (0.27,0.56) | <.001 |
|  | 1-3/month | 0.13 | 0.06 | (0.01,0.25) |  |
|  | <1/month | 0.15 | 0.04 | (0.08,0.22) |  |
| Birth year (ref: 1998-2005; current age: 18-24) | 1993-1998; age 25-29 | 0.28 | 0.08 | (0.13,0.43) | <.001 |
|  | 1983-1993; age 30-39 | 0.24 | 0.07 | (0.10,0.38) |  |
|  | 1973-1983; age 40-49 | 0.46 | 0.07 | (0.32,0.59) |  |
|  | 1963-1973; age 50-59 | 0.61 | 0.07 | (0.47,0.74) |  |
|  | 1953-1963; age 60-69 | 0.72 | 0.07 | (0.58,0.86) |  |
|  | 1943-1953; age 70-79 | 0.74 | 0.07 | (0.60,0.89) |  |
|  | 1943 or earlier; age 80+ | 0.68 | 0.09 | (0.49,0.86) |  |
| Gender (ref:Male) | Female | 0.17 | 0.03 | (0.10,0.23) | <.001 |
|  | Other | 0.50 | 0.41 | (-0.29,1.30) |  |
| Religious affiliation at age 12 (ref: No religion/Atheist/Agnostic) | Christianity | 0.15 | 0.04 | (0.07,0.23) | 0.001 |
|  | Islam | 0.34 | 0.14 | (0.07,0.61) |  |
|  | Collapsed affiliations with prevalence<3% | 0.20 | 0.17 | (-0.14,0.54) |  |

***Supplementary Table S18C. E-Values for Estimates and CI for Sweden***

| **Variable** | **Category** | **E-Value for Estimate** | **E-Value for 95% CI** |
| --- | --- | --- | --- |
| Relationship with mother (ref: Very bad/Somewhat bad) | Very good/Somewhat good | 1.17 | 1.00 |
| Relationship with father (ref: Very bad/Somewhat bad) | Very good/Somewhat good | 1.29 | 1.00 |
| Parent marital status (ref: Married) | Divorced | 1.36 | 1.14 |
|  | Single, never married | 1.05 | 1.00 |
|  | One or both parents had died | 1.21 | 1.00 |
| Subjective financial status of family growing up (ref: Got by) | Lived comfortably | 1.29 | 1.14 |
|  | Found it difficult | 1.19 | 1.00 |
|  | Found it very difficult | 1.50 | 1.00 |
| Abuse (ref: No) | Yes | 1.35 | 1.14 |
| Felt like an outsider in the family (ref: No) | Yes | 1.27 | 1.00 |
| Self-rated health growing up (ref: Good) | Excellent | 2.39 | 2.22 |
|  | Very good | 1.62 | 1.46 |
|  | Fair | 1.23 | 1.00 |
|  | Poor | 1.39 | 1.00 |
| Immigration status (ref: Born in this country) | Born in another country | 1.42 | 1.16 |
| Age 12 religious service attendance (ref: Never) | At least 1/week | 1.81 | 1.58 |
|  | 1-3/month | 1.35 | 1.08 |
|  | <1/month | 1.38 | 1.25 |
| Birth year (ref: 1998-2005; current age: 18-24) | 1993-1998; age 25-29 | 1.59 | 1.34 |
|  | 1983-1993; age 30-39 | 1.53 | 1.30 |
|  | 1973-1983; age 40-49 | 1.87 | 1.65 |
|  | 1963-1973; age 50-59 | 2.11 | 1.89 |
|  | 1953-1963; age 60-69 | 2.30 | 2.07 |
|  | 1943-1953; age 70-79 | 2.33 | 2.09 |
|  | 1943 or earlier; age 80+ | 2.22 | 1.92 |
| Gender (ref:Male) | Female | 1.41 | 1.30 |
|  | Other | 1.94 | 1.00 |
| Religious affiliation at age 12 (ref: No religion/Atheist/Agnostic) | Christianity | 1.38 | 1.23 |
|  | Islam | 1.69 | 1.25 |
|  | Collapsed affiliations with prevalence<3% | 1.47 | 1.00 |

***Supplementary Table S19A. Nationally Representative Childhood Descriptive Statistics for Tanzania***

| **Variable** | **Category** | **N (%)** |
| --- | --- | --- |
| Relationship with mother | Very good | 7739 (85%) |
|  | Somewhat good | 796 (9%) |
|  | Somewhat bad | 84 (1%) |
|  | Very bad | 84 (1%) |
|  | Does not apply | 303 (3%) |
|  | Missing | 70 (1%) |
| Relationships with father | Very good | 6831 (75%) |
|  | Somewhat good | 1101 (12%) |
|  | Somewhat bad | 203 (2%) |
|  | Very bad | 247 (3%) |
|  | Does not apply | 550 (6%) |
|  | Missing | 142 (2%) |
| Parent marital status | Parents married | 6929 (76%) |
|  | Divorced | 678 (7%) |
|  | Single, never married | 751 (8%) |
|  | One or both parents had died | 313 (3%) |
|  | Missing | 404 (4%) |
| Subjective financial status of family growing up | Lived comfortably | 2611 (29%) |
|  | Got by | 2909 (32%) |
|  | Found it difficult | 2679 (30%) |
|  | Found it very difficult | 814 (9%) |
|  | Missing | 61 (1%) |
| Abuse | Yes | 716 (8%) |
|  | No | 8328 (92%) |
|  | Missing | 32 (0%) |
| Felt like an outsider in the family | Yes | 734 (8%) |
|  | No | 8320 (92%) |
|  | Missing | 22 (0%) |
| Self-rated health growing up | Excellent | 2406 (27%) |
|  | Very good | 2036 (22%) |
|  | Good | 2946 (32%) |
|  | Fair | 1177 (13%) |
|  | Poor | 456 (5%) |
|  | Missing | 54 (1%) |
| Immigration status | Born in this country | 9048 (100%) |
|  | Born in another country | 25 (0%) |
|  | Missing | 1 (0%) |
| Age 12 religious service attendance | At least 1/week | 5580 (61%) |
|  | 1-3/month | 2383 (26%) |
|  | <1/ month | 333 (4%) |
|  | Never | 595 (7%) |
|  | Missing | 184 (2%) |
| Year of birth | 1998-2005; current age: 18-24 | 2284 (25%) |
|  | 1993-1998; age 25-29 | 1349 (15%) |
|  | 1983-1993; age 30-39 | 2060 (23%) |
|  | 1973-1983; age 40-49 | 1503 (17%) |
|  | 1963-1973; age 50-59 | 912 (10%) |
|  | 1953-1963; age 60-69 | 575 (6%) |
|  | 1943-1953; age 70-79 | 297 (3%) |
|  | 1943 or earlier; age 80+ | 93 (1%) |
|  | Missing | 2 (0%) |
| Gender | Male | 4299 (47%) |
|  | Female | 4776 (53%) |
| Religious affiliation at age 12 | Christianity | 5651 (62%) |
|  | Islam | 3060 (34%) |
|  | Baha’i | 1 (0%) |
|  | Primal, Animist, or Folk religion | 11 (0%) |
|  | No religion/Atheist/Agnostic | 345 (4%) |
|  | Missing | 7 (0%) |
| Race/ethnicity | African | 9060 (100%) |
|  | Indian | 3 (0%) |
|  | Arab | 11 (0%) |
|  | Missing | 2 (0%) |

***Supplementary Table S19B. Regression of Promoting Good on Childhood Predictors for Tanzania***

| **Variable** | **Category** | **Estimate** | **SE** | **95% CI** | **Global P-value** |
| --- | --- | --- | --- | --- | --- |
| Relationship with mother (ref: Very bad/Somewhat bad) | Very good/Somewhat good | 0.53 | 0.22 | (0.11,0.96) | 0.014 |
| Relationship with father (ref: Very bad/Somewhat bad) | Very good/Somewhat good | -0.17 | 0.15 | (-0.46,0.12) | 0.255 |
| Parent marital status (ref: Married) | Divorced | -0.11 | 0.14 | (-0.39,0.17) | 0.413 |
|  | Single, never married | 0.18 | 0.14 | (-0.09,0.46) |  |
|  | One or both parents had died | 0.04 | 0.18 | (-0.31,0.39) |  |
| Subjective financial status of family growing up (ref: Got by) | Lived comfortably | -0.12 | 0.09 | (-0.29,0.05) | 0.559 |
|  | Found it difficult | -0.05 | 0.09 | (-0.23,0.12) |  |
|  | Found it very difficult | -0.10 | 0.15 | (-0.38,0.19) |  |
| Abuse (ref: No) | Yes | -0.25 | 0.15 | (-0.54,0.04) | 0.084 |
| Felt like an outsider in the family (ref: No) | Yes | -0.22 | 0.14 | (-0.50,0.06) | 0.121 |
| Self-rated health growing up (ref: Good) | Excellent | 0.28 | 0.09 | (0.09,0.46) | 0.003 |
|  | Very good | 0.24 | 0.11 | (0.02,0.46) |  |
|  | Fair | 0.27 | 0.12 | (0.03,0.51) |  |
|  | Poor | -0.26 | 0.19 | (-0.63,0.11) |  |
| Immigration status (ref: Born in this country) | Born in another country | 0.18 | 0.52 | (-0.84,1.20) | 0.727 |
| Age 12 religious service attendance (ref: Never) | At least 1/week | -0.10 | 0.18 | (-0.45,0.25) | 0.098 |
|  | 1-3/month | -0.34 | 0.20 | (-0.73,0.04) |  |
|  | <1/month | -0.21 | 0.29 | (-0.78,0.35) |  |
| Birth year (ref: 1998-2005; current age: 18-24) | 1993-1998; age 25-29 | 0.12 | 0.11 | (-0.10,0.34) | 0.002 |
|  | 1983-1993; age 30-39 | -0.14 | 0.11 | (-0.35,0.07) |  |
|  | 1973-1983; age 40-49 | -0.09 | 0.12 | (-0.32,0.13) |  |
|  | 1963-1973; age 50-59 | -0.37 | 0.14 | (-0.64,-0.10) |  |
|  | 1953-1963; age 60-69 | -0.41 | 0.17 | (-0.74,-0.08) |  |
|  | 1943-1953; age 70-79 | -0.71 | 0.29 | (-1.29,-0.13) |  |
|  | 1943 or earlier; age 80+ | -0.94 | 0.53 | (-1.99,0.11) |  |
| Gender (ref:Male) | Female | -0.12 | 0.08 | (-0.28,0.03) | 0.113 |
| Religious affiliation at age 12 (ref: No religion/Atheist/Agnostic) | Christianity | 0.54 | 0.29 | (-0.05,1.12) | 0.014 |
|  | Islam | 0.47 | 0.30 | (-0.12,1.07) |  |
|  | Collapsed affiliations with prevalence<3% | 1.26 | 0.41 | (0.46,2.07) |  |
| Race/ethnicity (ref: plurality) | Race/ethnicity minority | -0.56 | 0.54 | (-1.63,0.51) | 0.305 |

***Supplementary Table S19C. E-Values for Estimates and CI for Tanzania***

| **Variable** | **Category** | **E-Value for Estimate** | **E-Value for 95% CI** |
| --- | --- | --- | --- |
| Relationship with mother (ref: Very bad/Somewhat bad) | Very good/Somewhat good | 1.69 | 1.24 |
| Relationship with father (ref: Very bad/Somewhat bad) | Very good/Somewhat good | 1.31 | 1.00 |
| Parent marital status (ref: Married) | Divorced | 1.24 | 1.00 |
|  | Single, never married | 1.33 | 1.00 |
|  | One or both parents had died | 1.14 | 1.00 |
| Subjective financial status of family growing up (ref: Got by) | Lived comfortably | 1.25 | 1.00 |
|  | Found it difficult | 1.15 | 1.00 |
|  | Found it very difficult | 1.22 | 1.00 |
| Abuse (ref: No) | Yes | 1.40 | 1.00 |
| Felt like an outsider in the family (ref: No) | Yes | 1.36 | 1.00 |
| Self-rated health growing up (ref: Good) | Excellent | 1.43 | 1.21 |
|  | Very good | 1.39 | 1.09 |
|  | Fair | 1.42 | 1.11 |
|  | Poor | 1.41 | 1.00 |
| Immigration status (ref: Born in this country) | Born in another country | 1.32 | 1.00 |
| Age 12 religious service attendance (ref: Never) | At least 1/week | 1.22 | 1.00 |
|  | 1-3/month | 1.50 | 1.00 |
|  | <1/month | 1.36 | 1.00 |
| Birth year (ref: 1998-2005; current age: 18-24) | 1993-1998; age 25-29 | 1.25 | 1.00 |
|  | 1983-1993; age 30-39 | 1.27 | 1.00 |
|  | 1973-1983; age 40-49 | 1.21 | 1.00 |
|  | 1963-1973; age 50-59 | 1.53 | 1.23 |
|  | 1953-1963; age 60-69 | 1.57 | 1.20 |
|  | 1943-1953; age 70-79 | 1.86 | 1.27 |
|  | 1943 or earlier; age 80+ | 2.09 | 1.00 |
| Gender (ref:Male) | Female | 1.25 | 1.00 |
| Religious affiliation at age 12 (ref: No religion/Atheist/Agnostic) | Christianity | 1.69 | 1.00 |
|  | Islam | 1.63 | 1.00 |
|  | Collapsed affiliations with prevalence<3% | 2.44 | 1.61 |
| Race/ethnicity (ref: plurality) | Race/ethnicity minority | 1.71 | 1.00 |

***Supplementary Table S20A. Nationally Representative Childhood Descriptive Statistics for Türkiye***

| **Variable** | **Category** | **N (%)** |
| --- | --- | --- |
| Relationship with mother | Very good | 970 (66%) |
|  | Somewhat good | 401 (27%) |
|  | Somewhat bad | 48 (3%) |
|  | Very bad | 26 (2%) |
|  | Does not apply | 21 (1%) |
|  | Missing | 7 (0%) |
| Relationships with father | Very good | 795 (54%) |
|  | Somewhat good | 425 (29%) |
|  | Somewhat bad | 73 (5%) |
|  | Very bad | 95 (6%) |
|  | Does not apply | 60 (4%) |
|  | Missing | 25 (2%) |
| Parent marital status | Parents married | 1325 (90%) |
|  | Divorced | 57 (4%) |
|  | Single, never married | 7 (0%) |
|  | One or both parents had died | 61 (4%) |
|  | Missing | 23 (2%) |
| Subjective financial status of family growing up | Lived comfortably | 498 (34%) |
|  | Got by | 647 (44%) |
|  | Found it difficult | 218 (15%) |
|  | Found it very difficult | 108 (7%) |
|  | Missing | 2 (0%) |
| Abuse | Yes | 158 (11%) |
|  | No | 1290 (88%) |
|  | Missing | 25 (2%) |
| Felt like an outsider in the family | Yes | 157 (11%) |
|  | No | 1306 (89%) |
|  | Missing | 9 (1%) |
| Self-rated health growing up | Excellent | 377 (26%) |
|  | Very good | 410 (28%) |
|  | Good | 419 (28%) |
|  | Fair | 220 (15%) |
|  | Poor | 47 (3%) |
|  | Missing | 0 (0%) |
| Immigration status | Born in this country | 1415 (96%) |
|  | Born in another country | 58 (4%) |
| Age 12 religious service attendance | At least 1/week | 609 (41%) |
|  | 1-3/month | 238 (16%) |
|  | <1/ month | 225 (15%) |
|  | Never | 383 (26%) |
|  | Missing | 18 (1%) |
| Year of birth | 1998-2005; current age: 18-24 | 222 (15%) |
|  | 1993-1998; age 25-29 | 152 (10%) |
|  | 1983-1993; age 30-39 | 315 (21%) |
|  | 1973-1983; age 40-49 | 312 (21%) |
|  | 1963-1973; age 50-59 | 225 (15%) |
|  | 1953-1963; age 60-69 | 164 (11%) |
|  | 1943-1953; age 70-79 | 65 (4%) |
|  | 1943 or earlier; age 80+ | 18 (1%) |
| Gender | Male | 754 (51%) |
|  | Female | 719 (49%) |
| Religious affiliation at age 12 | Christianity | 1 (0%) |
|  | Islam | 1439 (98%) |
|  | Judaism | 1 (0%) |
|  | No religion/Atheist/Agnostic | 13 (1%) |
|  | Missing | 19 (1%) |
| Race/ethnicity | Turkish | 1030 (70%) |
|  | Kurdish/Zaza | 252 (17%) |
|  | Arab | 51 (3%) |
|  | Laz | 25 (2%) |
|  | Circassian | 19 (1%) |
|  | Bosnian | 5 (0%) |
|  | Armenian | 1 (0%) |
|  | Georgian | 4 (0%) |
|  | Uyghur | 1 (0%) |
|  | Albanian | 8 (1%) |
|  | Greek | 1 (0%) |
|  | Azeri | 9 (1%) |
|  | Other | 58 (4%) |
|  | Missing | 9 (1%) |

***Supplementary Table S20B. Regression of Promoting Good on Childhood Predictors for Türkiye***

| **Variable** | **Category** | **Estimate** | **SE** | **95% CI** | **Global P-value** |
| --- | --- | --- | --- | --- | --- |
| Relationship with mother (ref: Very bad/Somewhat bad) | Very good/Somewhat good | 0.51 | 0.36 | (-0.19,1.21) | 0.153 |
| Relationship with father (ref: Very bad/Somewhat bad) | Very good/Somewhat good | 0.08 | 0.23 | (-0.37,0.54) | 0.712 |
| Parent marital status (ref: Married) | Divorced | 0.53 | 0.35 | (-0.15,1.21) | 0.337 |
|  | Single, never married | 0.35 | 0.62 | (-0.85,1.56) |  |
|  | One or both parents had died | -0.32 | 0.39 | (-1.08,0.43) |  |
| Subjective financial status of family growing up (ref: Got by) | Lived comfortably | 0.42 | 0.17 | (0.09,0.76) | 0.051 |
|  | Found it difficult | -0.07 | 0.25 | (-0.56,0.43) |  |
|  | Found it very difficult | -0.11 | 0.40 | (-0.90,0.68) |  |
| Abuse (ref: No) | Yes | -0.14 | 0.23 | (-0.60,0.32) | 0.548 |
| Felt like an outsider in the family (ref: No) | Yes | -0.05 | 0.23 | (-0.51,0.41) | 0.822 |
| Self-rated health growing up (ref: Good) | Excellent | -0.15 | 0.21 | (-0.57,0.26) | 0.325 |
|  | Very good | 0.04 | 0.19 | (-0.33,0.42) |  |
|  | Fair | -0.28 | 0.26 | (-0.79,0.23) |  |
|  | Poor | -0.97 | 0.59 | (-2.13,0.18) |  |
| Immigration status (ref: Born in this country) | Born in another country | 0.56 | 0.38 | (-0.19,1.31) | 0.142 |
| Age 12 religious service attendance (ref: Never) | At least 1/week | 0.27 | 0.22 | (-0.16,0.69) | 0.159 |
|  | 1-3/month | 0.44 | 0.22 | (0.00,0.88) |  |
|  | <1/month | 0.05 | 0.24 | (-0.43,0.53) |  |
| Birth year (ref: 1998-2005; current age: 18-24) | 1993-1998; age 25-29 | 0.51 | 0.25 | (0.01,1.01) | 0.019 |
|  | 1983-1993; age 30-39 | 0.27 | 0.23 | (-0.19,0.73) |  |
|  | 1973-1983; age 40-49 | 0.63 | 0.22 | (0.20,1.06) |  |
|  | 1963-1973; age 50-59 | 0.46 | 0.26 | (-0.04,0.96) |  |
|  | 1953-1963; age 60-69 | -0.04 | 0.33 | (-0.69,0.60) |  |
|  | 1943-1953; age 70-79 | -1.05 | 0.60 | (-2.23,0.14) |  |
|  | 1943 or earlier; age 80+ | 0.32 | 0.54 | (-0.75,1.38) |  |
| Gender (ref:Male) | Female | 0.05 | 0.17 | (-0.29,0.38) | 0.778 |
| Religious affiliation at age 12 (ref: Islam) | Collapsed affiliations with prevalence<3% | 0.33 | 0.46 | (-0.57,1.22) | 0.460 |
| Race/ethnicity (ref: plurality) | Race/ethnicity minority | -0.45 | 0.19 | (-0.81,-0.08) | 0.016 |

***Supplementary Table S20C. E-Values for Estimates and CI for Türkiye***

| **Variable** | **Category** | **E-Value for Estimate** | **E-Value for 95% CI** |
| --- | --- | --- | --- |
| Relationship with mother (ref: Very bad/Somewhat bad) | Very good/Somewhat good | 1.73 | 1.00 |
| Relationship with father (ref: Very bad/Somewhat bad) | Very good/Somewhat good | 1.22 | 1.00 |
| Parent marital status (ref: Married) | Divorced | 1.75 | 1.00 |
|  | Single, never married | 1.55 | 1.00 |
|  | One or both parents had died | 1.52 | 1.00 |
| Subjective financial status of family growing up (ref: Got by) | Lived comfortably | 1.63 | 1.23 |
|  | Found it difficult | 1.19 | 1.00 |
|  | Found it very difficult | 1.26 | 1.00 |
| Abuse (ref: No) | Yes | 1.29 | 1.00 |
| Felt like an outsider in the family (ref: No) | Yes | 1.15 | 1.00 |
| Self-rated health growing up (ref: Good) | Excellent | 1.32 | 1.00 |
|  | Very good | 1.15 | 1.00 |
|  | Fair | 1.47 | 1.00 |
|  | Poor | 2.26 | 1.00 |
| Immigration status (ref: Born in this country) | Born in another country | 1.78 | 1.00 |
| Age 12 religious service attendance (ref: Never) | At least 1/week | 1.45 | 1.00 |
|  | 1-3/month | 1.66 | 1.05 |
|  | <1/month | 1.16 | 1.00 |
| Birth year (ref: 1998-2005; current age: 18-24) | 1993-1998; age 25-29 | 1.72 | 1.05 |
|  | 1983-1993; age 30-39 | 1.46 | 1.00 |
|  | 1973-1983; age 40-49 | 1.86 | 1.37 |
|  | 1963-1973; age 50-59 | 1.67 | 1.00 |
|  | 1953-1963; age 60-69 | 1.15 | 1.00 |
|  | 1943-1953; age 70-79 | 2.36 | 1.00 |
|  | 1943 or earlier; age 80+ | 1.51 | 1.00 |
| Gender (ref:Male) | Female | 1.16 | 1.00 |
| Religious affiliation at age 12 (ref: Islam) | Collapsed affiliations with prevalence<3% | 1.52 | 1.00 |
| Race/ethnicity (ref: plurality) | Race/ethnicity minority | 1.66 | 1.22 |

***Supplementary Table S21A. Nationally Representative Childhood Descriptive Statistics for United Kingdom***

| **Variable** | **Category** | **N (%)** |
| --- | --- | --- |
| Relationship with mother | Very good | 3435 (64%) |
|  | Somewhat good | 1338 (25%) |
|  | Somewhat bad | 325 (6%) |
|  | Very bad | 150 (3%) |
|  | Does not apply | 92 (2%) |
|  | Missing | 27 (1%) |
| Relationships with father | Very good | 2907 (54%) |
|  | Somewhat good | 1383 (26%) |
|  | Somewhat bad | 407 (8%) |
|  | Very bad | 321 (6%) |
|  | Does not apply | 321 (6%) |
|  | Missing | 29 (1%) |
| Parent marital status | Parents married | 4343 (81%) |
|  | Divorced | 481 (9%) |
|  | Single, never married | 315 (6%) |
|  | One or both parents had died | 154 (3%) |
|  | Missing | 75 (1%) |
| Subjective financial status of family growing up | Lived comfortably | 2552 (48%) |
|  | Got by | 1933 (36%) |
|  | Found it difficult | 632 (12%) |
|  | Found it very difficult | 230 (4%) |
|  | Missing | 22 (0%) |
| Abuse | Yes | 864 (16%) |
|  | No | 4455 (83%) |
|  | Missing | 49 (1%) |
| Felt like an outsider in the family | Yes | 1017 (19%) |
|  | No | 4308 (80%) |
|  | Missing | 43 (1%) |
| Self-rated health growing up | Excellent | 2154 (40%) |
|  | Very good | 1736 (32%) |
|  | Good | 995 (19%) |
|  | Fair | 332 (6%) |
|  | Poor | 130 (2%) |
|  | Missing | 20 (0%) |
| Immigration status | Born in this country | 4659 (87%) |
|  | Born in another country | 682 (13%) |
|  | Missing | 27 (0%) |
| Age 12 religious service attendance | At least 1/week | 1732 (32%) |
|  | 1-3/month | 733 (14%) |
|  | <1/ month | 903 (17%) |
|  | Never | 1972 (37%) |
|  | Missing | 28 (1%) |
| Year of birth | 1998-2005; current age: 18-24 | 490 (9%) |
|  | 1993-1998; age 25-29 | 391 (7%) |
|  | 1983-1993; age 30-39 | 946 (18%) |
|  | 1973-1983; age 40-49 | 827 (15%) |
|  | 1963-1973; age 50-59 | 949 (18%) |
|  | 1953-1963; age 60-69 | 889 (17%) |
|  | 1943-1953; age 70-79 | 711 (13%) |
|  | 1943 or earlier; age 80+ | 163 (3%) |
|  | Missing | 1 (0%) |
| Gender | Male | 2557 (48%) |
|  | Female | 2789 (52%) |
|  | Other | 14 (0%) |
|  | Missing | 9 (0%) |
| Religious affiliation at age 12 | Christianity | 3461 (64%) |
|  | Islam | 230 (4%) |
|  | Hinduism | 88 (2%) |
|  | Buddhism | 15 (0%) |
|  | Judaism | 59 (1%) |
|  | Sikhism | 30 (1%) |
|  | Baha’i | 5 (0%) |
|  | Jainism | 0 (0%) |
|  | Taoism | 2 (0%) |
|  | Confucianism | 3 (0%) |
|  | Primal, Animist, or Folk religion | 22 (0%) |
|  | Some other religion | 24 (0%) |
|  | No religion/Atheist/Agnostic | 1409 (26%) |
|  | Missing | 21 (0%) |
| Race/ethnicity | Asian | 426 (8%) |
|  | Black | 152 (3%) |
|  | White | 4647 (87%) |
|  | Other | 96 (2%) |
|  | Missing | 47 (1%) |

***Supplementary Table S21B. Regression of Promoting Good on Childhood Predictors for United Kingdom***

| **Variable** | **Category** | **Estimate** | **SE** | **95% CI** | **Global P-value** |
| --- | --- | --- | --- | --- | --- |
| Relationship with mother (ref: Very bad/Somewhat bad) | Very good/Somewhat good | 0.09 | 0.14 | (-0.18,0.36) | 0.492 |
| Relationship with father (ref: Very bad/Somewhat bad) | Very good/Somewhat good | 0.16 | 0.11 | (-0.05,0.37) | 0.121 |
| Parent marital status (ref: Married) | Divorced | -0.21 | 0.13 | (-0.47,0.06) | 0.265 |
|  | Single, never married | -0.26 | 0.19 | (-0.63,0.12) |  |
|  | One or both parents had died | -0.14 | 0.20 | (-0.53,0.25) |  |
| Subjective financial status of family growing up (ref: Got by) | Lived comfortably | 0.11 | 0.08 | (-0.04,0.26) | 0.220 |
|  | Found it difficult | -0.13 | 0.14 | (-0.39,0.14) |  |
|  | Found it very difficult | -0.09 | 0.21 | (-0.51,0.33) |  |
| Abuse (ref: No) | Yes | -0.06 | 0.11 | (-0.27,0.15) | 0.580 |
| Felt like an outsider in the family (ref: No) | Yes | 0.10 | 0.10 | (-0.10,0.29) | 0.315 |
| Self-rated health growing up (ref: Good) | Excellent | 0.19 | 0.10 | (-0.01,0.38) | <.001 |
|  | Very good | 0.08 | 0.09 | (-0.11,0.26) |  |
|  | Fair | -0.59 | 0.18 | (-0.95,-0.23) |  |
|  | Poor | -0.15 | 0.24 | (-0.62,0.33) |  |
| Immigration status (ref: Born in this country) | Born in another country | 0.27 | 0.09 | (0.09,0.46) | 0.004 |
| Age 12 religious service attendance (ref: Never) | At least 1/week | 0.33 | 0.10 | (0.14,0.52) | 0.002 |
|  | 1-3/month | 0.33 | 0.11 | (0.11,0.56) |  |
|  | <1/month | 0.12 | 0.10 | (-0.09,0.32) |  |
| Birth year (ref: 1998-2005; current age: 18-24) | 1993-1998; age 25-29 | 0.22 | 0.18 | (-0.12,0.57) | 0.813 |
|  | 1983-1993; age 30-39 | 0.05 | 0.17 | (-0.27,0.38) |  |
|  | 1973-1983; age 40-49 | 0.10 | 0.16 | (-0.22,0.41) |  |
|  | 1963-1973; age 50-59 | 0.20 | 0.16 | (-0.12,0.52) |  |
|  | 1953-1963; age 60-69 | 0.12 | 0.17 | (-0.22,0.46) |  |
|  | 1943-1953; age 70-79 | 0.13 | 0.17 | (-0.21,0.47) |  |
|  | 1943 or earlier; age 80+ | 0.22 | 0.24 | (-0.25,0.68) |  |
| Gender (ref:Male) | Female | 0.00 | 0.07 | (-0.13,0.14) | 0.954 |
|  | Other | -0.34 | 1.21 | (-2.71,2.04) |  |
| Religious affiliation at age 12 (ref: No religion/Atheist/Agnostic) | Christianity | 0.24 | 0.10 | (0.06,0.43) | 0.018 |
|  | Islam | 0.15 | 0.21 | (-0.26,0.56) |  |
|  | Collapsed affiliations with prevalence<3% | -0.15 | 0.19 | (-0.52,0.22) |  |
| Race/ethnicity (ref: plurality) | Race/ethnicity minority | 0.04 | 0.14 | (-0.24,0.32) | 0.743 |

***Supplementary Table S21C. E-Values for Estimates and CI for United Kingdom***

| **Variable** | **Category** | **E-Value for Estimate** | **E-Value for 95% CI** |
| --- | --- | --- | --- |
| Relationship with mother (ref: Very bad/Somewhat bad) | Very good/Somewhat good | 1.27 | 1.00 |
| Relationship with father (ref: Very bad/Somewhat bad) | Very good/Somewhat good | 1.39 | 1.00 |
| Parent marital status (ref: Married) | Divorced | 1.46 | 1.00 |
|  | Single, never married | 1.53 | 1.00 |
|  | One or both parents had died | 1.35 | 1.00 |
| Subjective financial status of family growing up (ref: Got by) | Lived comfortably | 1.30 | 1.00 |
|  | Found it difficult | 1.33 | 1.00 |
|  | Found it very difficult | 1.26 | 1.00 |
| Abuse (ref: No) | Yes | 1.20 | 1.00 |
| Felt like an outsider in the family (ref: No) | Yes | 1.28 | 1.00 |
| Self-rated health growing up (ref: Good) | Excellent | 1.43 | 1.00 |
|  | Very good | 1.25 | 1.00 |
|  | Fair | 2.03 | 1.49 |
|  | Poor | 1.36 | 1.00 |
| Immigration status (ref: Born in this country) | Born in another country | 1.56 | 1.26 |
| Age 12 religious service attendance (ref: Never) | At least 1/week | 1.64 | 1.35 |
|  | 1-3/month | 1.65 | 1.30 |
|  | <1/month | 1.31 | 1.00 |
| Birth year (ref: 1998-2005; current age: 18-24) | 1993-1998; age 25-29 | 1.48 | 1.00 |
|  | 1983-1993; age 30-39 | 1.19 | 1.00 |
|  | 1973-1983; age 40-49 | 1.28 | 1.00 |
|  | 1963-1973; age 50-59 | 1.45 | 1.00 |
|  | 1953-1963; age 60-69 | 1.32 | 1.00 |
|  | 1943-1953; age 70-79 | 1.34 | 1.00 |
|  | 1943 or earlier; age 80+ | 1.47 | 1.00 |
| Gender (ref:Male) | Female | 1.05 | 1.00 |
|  | Other | 1.65 | 1.00 |
| Religious affiliation at age 12 (ref: No religion/Atheist/Agnostic) | Christianity | 1.52 | 1.20 |
|  | Islam | 1.37 | 1.00 |
|  | Collapsed affiliations with prevalence<3% | 1.37 | 1.00 |
| Race/ethnicity (ref: plurality) | Race/ethnicity minority | 1.17 | 1.00 |

***Supplementary Table S22A. Nationally Representative Childhood Descriptive Statistics for United States***

| **Variable** | **Category** | **N (%)** |
| --- | --- | --- |
| Relationship with mother | Very good | 20590 (54%) |
|  | Somewhat good | 11525 (30%) |
|  | Somewhat bad | 3523 (9%) |
|  | Very bad | 1874 (5%) |
|  | Does not apply | 694 (2%) |
|  | Missing | 106 (0%) |
| Relationships with father | Very good | 15313 (40%) |
|  | Somewhat good | 12665 (33%) |
|  | Somewhat bad | 4879 (13%) |
|  | Very bad | 2604 (7%) |
|  | Does not apply | 2811 (7%) |
|  | Missing | 38 (0%) |
| Parent marital status | Parents married | 27415 (72%) |
|  | Divorced | 6325 (17%) |
|  | Single, never married | 3048 (8%) |
|  | One or both parents had died | 1024 (3%) |
|  | Missing | 500 (1%) |
| Subjective financial status of family growing up | Lived comfortably | 15116 (39%) |
|  | Got by | 15682 (41%) |
|  | Found it difficult | 5152 (13%) |
|  | Found it very difficult | 2342 (6%) |
|  | Missing | 19 (0%) |
| Abuse | Yes | 10026 (26%) |
|  | No | 28045 (73%) |
|  | Missing | 242 (1%) |
| Felt like an outsider in the family | Yes | 10185 (27%) |
|  | No | 27714 (72%) |
|  | Missing | 413 (1%) |
| Self-rated health growing up | Excellent | 16866 (44%) |
|  | Very good | 12108 (32%) |
|  | Good | 6444 (17%) |
|  | Fair | 2303 (6%) |
|  | Poor | 520 (1%) |
|  | Missing | 71 (0%) |
| Immigration status | Born in this country | 34865 (91%) |
|  | Born in another country | 3020 (8%) |
|  | Missing | 427 (1%) |
| Age 12 religious service attendance | At least 1/week | 18609 (49%) |
|  | 1-3/month | 6644 (17%) |
|  | <1/ month | 5829 (15%) |
|  | Never | 7085 (18%) |
|  | Missing | 145 (0%) |
| Year of birth | 1998-2005; current age: 18-24 | 2682 (7%) |
|  | 1993-1998; age 25-29 | 3540 (9%) |
|  | 1983-1993; age 30-39 | 7284 (19%) |
|  | 1973-1983; age 40-49 | 5649 (15%) |
|  | 1963-1973; age 50-59 | 6745 (18%) |
|  | 1953-1963; age 60-69 | 6832 (18%) |
|  | 1943-1953; age 70-79 | 4054 (11%) |
|  | 1943 or earlier; age 80+ | 1525 (4%) |
| Gender | Male | 18222 (48%) |
|  | Female | 19562 (51%) |
|  | Other | 392 (1%) |
|  | Missing | 136 (0%) |
| Religious affiliation at age 12 | Christianity | 30444 (79%) |
|  | Islam | 220 (1%) |
|  | Hinduism | 203 (1%) |
|  | Buddhism | 172 (0%) |
|  | Judaism | 787 (2%) |
|  | Sikhism | 47 (0%) |
|  | Baha’i | 4 (0%) |
|  | Jainism | 18 (0%) |
|  | Shinto | 6 (0%) |
|  | Taoism | 17 (0%) |
|  | Confucianism | 8 (0%) |
|  | Primal, Animist, or Folk religion | 67 (0%) |
|  | Some other religion | 359 (1%) |
|  | No religion/Atheist/Agnostic | 5845 (15%) |
|  | Missing | 115 (0%) |
| Race/ethnicity | White | 23605 (62%) |
|  | Other | 997 (3%) |
|  | Black | 4501 (12%) |
|  | Asian | 2466 (6%) |
|  | Hispanic | 6724 (18%) |
|  | Missing | 20 (0%) |

***Supplementary Table S22B. Regression of Promoting Good on Childhood Predictors for United States***

| **Variable** | **Category** | **Estimate** | **SE** | **95% CI** | **Global P-value** |
| --- | --- | --- | --- | --- | --- |
| Relationship with mother (ref: Very bad/Somewhat bad) | Very good/Somewhat good | 0.11 | 0.08 | (-0.06,0.27) | 0.206 |
| Relationship with father (ref: Very bad/Somewhat bad) | Very good/Somewhat good | 0.14 | 0.07 | (-0.01,0.28) | 0.059 |
| Parent marital status (ref: Married) | Divorced | 0.05 | 0.08 | (-0.11,0.20) | 0.334 |
|  | Single, never married | 0.14 | 0.16 | (-0.17,0.46) |  |
|  | One or both parents had died | -0.41 | 0.26 | (-0.92,0.10) |  |
| Subjective financial status of family growing up (ref: Got by) | Lived comfortably | 0.14 | 0.05 | (0.05,0.23) | 0.006 |
|  | Found it difficult | 0.04 | 0.09 | (-0.13,0.21) |  |
|  | Found it very difficult | 0.28 | 0.13 | (0.03,0.53) |  |
| Abuse (ref: No) | Yes | -0.04 | 0.07 | (-0.18,0.10) | 0.553 |
| Felt like an outsider in the family (ref: No) | Yes | -0.05 | 0.08 | (-0.20,0.10) | 0.517 |
| Self-rated health growing up (ref: Good) | Excellent | 0.71 | 0.08 | (0.56,0.86) | <.001 |
|  | Very good | 0.32 | 0.08 | (0.17,0.47) |  |
|  | Fair | -0.26 | 0.19 | (-0.62,0.10) |  |
|  | Poor | -0.03 | 0.39 | (-0.79,0.74) |  |
| Immigration status (ref: Born in this country) | Born in another country | 0.30 | 0.10 | (0.10,0.49) | 0.003 |
| Age 12 religious service attendance (ref: Never) | At least 1/week | 0.28 | 0.08 | (0.12,0.44) | <.001 |
|  | 1-3/month | 0.12 | 0.09 | (-0.06,0.30) |  |
|  | <1/month | 0.08 | 0.10 | (-0.11,0.27) |  |
| Birth year (ref: 1998-2005; current age: 18-24) | 1993-1998; age 25-29 | 0.23 | 0.21 | (-0.18,0.63) | <.001 |
|  | 1983-1993; age 30-39 | 0.48 | 0.18 | (0.12,0.83) |  |
|  | 1973-1983; age 40-49 | 0.47 | 0.18 | (0.11,0.83) |  |
|  | 1963-1973; age 50-59 | 0.63 | 0.17 | (0.29,0.97) |  |
|  | 1953-1963; age 60-69 | 0.86 | 0.17 | (0.53,1.20) |  |
|  | 1943-1953; age 70-79 | 1.04 | 0.17 | (0.70,1.37) |  |
|  | 1943 or earlier; age 80+ | 1.06 | 0.18 | (0.71,1.42) |  |
| Gender (ref:Male) | Female | -0.02 | 0.05 | (-0.11,0.07) | 0.550 |
|  | Other | 0.30 | 0.30 | (-0.30,0.90) |  |
| Religious affiliation at age 12 (ref: No religion/Atheist/Agnostic) | Christianity | 0.22 | 0.10 | (0.03,0.42) | 0.062 |
|  | Collapsed affiliations with prevalence<3% | 0.14 | 0.16 | (-0.18,0.46) |  |
| Race/ethnicity (ref: plurality) | Race/ethnicity minority | 0.09 | 0.05 | (-0.01,0.20) | 0.090 |

***Supplementary Table S22C. E-Values for Estimates and CI for United States***

| **Variable** | **Category** | **E-Value for Estimate** | **E-Value for 95% CI** |
| --- | --- | --- | --- |
| Relationship with mother (ref: Very bad/Somewhat bad) | Very good/Somewhat good | 1.31 | 1.00 |
| Relationship with father (ref: Very bad/Somewhat bad) | Very good/Somewhat good | 1.36 | 1.00 |
| Parent marital status (ref: Married) | Divorced | 1.19 | 1.00 |
|  | Single, never married | 1.37 | 1.00 |
|  | One or both parents had died | 1.80 | 1.00 |
| Subjective financial status of family growing up (ref: Got by) | Lived comfortably | 1.37 | 1.20 |
|  | Found it difficult | 1.16 | 1.00 |
|  | Found it very difficult | 1.60 | 1.15 |
| Abuse (ref: No) | Yes | 1.17 | 1.00 |
| Felt like an outsider in the family (ref: No) | Yes | 1.19 | 1.00 |
| Self-rated health growing up (ref: Good) | Excellent | 2.29 | 2.04 |
|  | Very good | 1.66 | 1.42 |
|  | Fair | 1.56 | 1.00 |
|  | Poor | 1.14 | 1.00 |
| Immigration status (ref: Born in this country) | Born in another country | 1.62 | 1.30 |
| Age 12 religious service attendance (ref: Never) | At least 1/week | 1.60 | 1.34 |
|  | 1-3/month | 1.33 | 1.00 |
|  | <1/month | 1.26 | 1.00 |
| Birth year (ref: 1998-2005; current age: 18-24) | 1993-1998; age 25-29 | 1.51 | 1.00 |
|  | 1983-1993; age 30-39 | 1.90 | 1.34 |
|  | 1973-1983; age 40-49 | 1.90 | 1.32 |
|  | 1963-1973; age 50-59 | 2.15 | 1.61 |
|  | 1953-1963; age 60-69 | 2.55 | 1.99 |
|  | 1943-1953; age 70-79 | 2.88 | 2.27 |
|  | 1943 or earlier; age 80+ | 2.93 | 2.29 |
| Gender (ref:Male) | Female | 1.11 | 1.00 |
|  | Other | 1.63 | 1.00 |
| Religious affiliation at age 12 (ref: No religion/Atheist/Agnostic) | Christianity | 1.51 | 1.16 |
|  | Collapsed affiliations with prevalence<3% | 1.37 | 1.00 |
| Race/ethnicity (ref: plurality) | Race/ethnicity minority | 1.28 | 1.00 |

***Table S23. Population Weighted Meta-Analysis of Regression Results.***

| Variable | Category | Est | 95% CI | SE |
| --- | --- | --- | --- | --- |
| Relationship with mother | (Ref: Very bad/somewhat bad) |  |  |  |
|  | Very good/somewhat good | 0.19 | (-0.00,0.39) | 0.099 |
| Relationship with father | (Ref: Very bad/somewhat bad) |  |  |  |
|  | Very good/somewhat good | -0.05 | (-0.21,0.10) | 0.080 |
| Parent marital status | (Ref: Parents married) |  |  |  |
|  | Divorced | -0.03 | (-0.22,0.16) | 0.096 |
|  | Single, never married | -0.00 | (-0.12,0.12) | 0.061 |
|  | One or both parents had died | 0.13 | (0.02,0.25) | 0.060 |
| Subjective financial status of family growing up | (Ref: Got by) |  |  |  |
|  | Lived comfortably | 0.03 | (-0.04,0.09) | 0.031 |
|  | Found it difficult | -0.06 | (-0.14,0.01) | 0.040 |
|  | Found it very difficult | -0.26 | (-0.36,-0.16) | 0.052 |
| Abuse | (Ref: No) |  |  |  |
|  | Yes | -0.14 | (-0.23,-0.05) | 0.044 |
| Felt like an outsider in the family | (Ref: No) |  |  |  |
|  | Yes | -0.08 | (-0.16,0.01) | 0.042 |
| Self-rated health growing up | (Ref: Good) |  |  |  |
|  | Excellent | 0.29 | (0.22,0.37) | 0.039 |
|  | Very good | 0.15 | (0.09,0.22) | 0.033 |
|  | Fair | -0.18 | (-0.26,-0.09) | 0.045 |
|  | Poor | -0.02 | (-0.20,0.16) | 0.090 |
| Immigration status | (Ref: Born in this country) |  |  |  |
|  | Born in another country | -0.32 | (-0.57,-0.08) | 0.124 |
| Age 12 religious service attendance | (Ref: Never) |  |  |  |
|  | At least 1/week | 0.24 | (0.14,0.33) | 0.047 |
|  | 1-3/month | 0.19 | (0.09,0.28) | 0.050 |
|  | <1/month | 0.05 | (-0.06,0.15) | 0.054 |
| Year of birth | (Ref: 1998-2005; age 18-24) |  |  |  |
|  | 1993-1998; age 25-29 | 0.06 | (-0.03,0.15) | 0.047 |
|  | 1983-1993; age 30-39 | 0.05 | (-0.03,0.13) | 0.042 |
|  | 1973-1983; age 40-49 | 0.03 | (-0.06,0.12) | 0.045 |
|  | 1963-1973; age 50-59 | 0.06 | (-0.04,0.16) | 0.051 |
|  | 1953-1963; age 60-69 | 0.04 | (-0.07,0.15) | 0.058 |
|  | 1943-1953; age 70-79 | 0.03 | (-0.14,0.20) | 0.085 |
|  | 1943 or earlier; age 80+ | -0.42 | (-0.91,0.08) | 0.252 |
| Gender | (Ref: Male) |  |  |  |
|  | Female | 0.06 | (0.01,0.11) | 0.025 |
|  | Other | -0.44 | (-0.80,-0.08) | 0.183 |

***Table S24. Population Weighted Meta-Analysis of E-Values.***

| Variable | Category | evalue | evalue.limit |
| --- | --- | --- | --- |
| Relationship with mother | (Ref: Very bad/somewhat bad) |  |  |
|  | Very good/somewhat good | 1.41 | 1.00 |
| Relationship with father | (Ref: Very bad/somewhat bad) |  |  |
|  | Very good/somewhat good | 1.18 | 1.00 |
| Parent marital status | (Ref: Parents married) |  |  |
|  | Divorced | 1.13 | 1.00 |
|  | Single, never married | 1.03 | 1.00 |
|  | One or both parents had died | 1.32 | 1.09 |
| Subjective financial status of family growing up | (Ref: Got by) |  |  |
|  | Lived comfortably | 1.12 | 1.00 |
|  | Found it difficult | 1.20 | 1.00 |
|  | Found it very difficult | 1.51 | 1.36 |
| Abuse | (Ref: No) |  |  |
|  | Yes | 1.33 | 1.19 |
| Felt like an outsider in the family | (Ref: No) |  |  |
|  | Yes | 1.23 | 1.00 |
| Self-rated health growing up | (Ref: Good) |  |  |
|  | Excellent | 1.55 | 1.45 |
|  | Very good | 1.36 | 1.25 |
|  | Fair | 1.39 | 1.25 |
|  | Poor | 1.11 | 1.00 |
| Immigration status | (Ref: Born in this country) |  |  |
|  | Born in another country | 1.59 | 1.24 |
| Age 12 religious service attendance | (Ref: Never) |  |  |
|  | At least 1/week | 1.47 | 1.34 |
|  | 1-3/month | 1.40 | 1.25 |
|  | <1/month | 1.17 | 1.00 |
| Year of birth | (Ref: 1998-2005; age 18-24) |  |  |
|  | 1993-1998; age 25-29 | 1.20 | 1.00 |
|  | 1983-1993; age 30-39 | 1.18 | 1.00 |
|  | 1973-1983; age 40-49 | 1.13 | 1.00 |
|  | 1963-1973; age 50-59 | 1.19 | 1.00 |
|  | 1953-1963; age 60-69 | 1.15 | 1.00 |
|  | 1943-1953; age 70-79 | 1.13 | 1.00 |
|  | 1943 or earlier; age 80+ | 1.72 | 1.00 |
| Gender | (Ref: Male) |  |  |
|  | Female | 1.20 | 1.07 |
|  | Other | 1.75 | 1.24 |

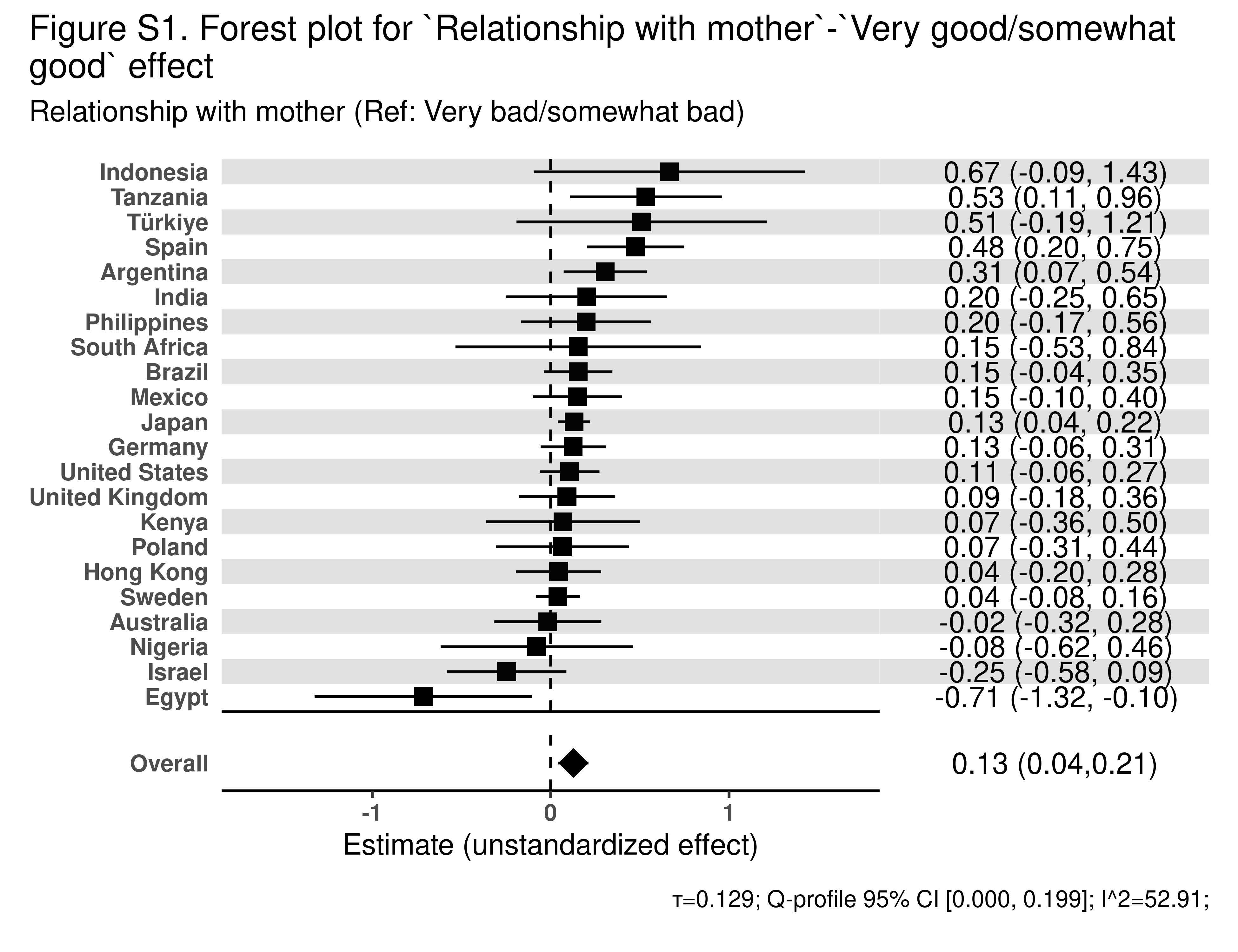

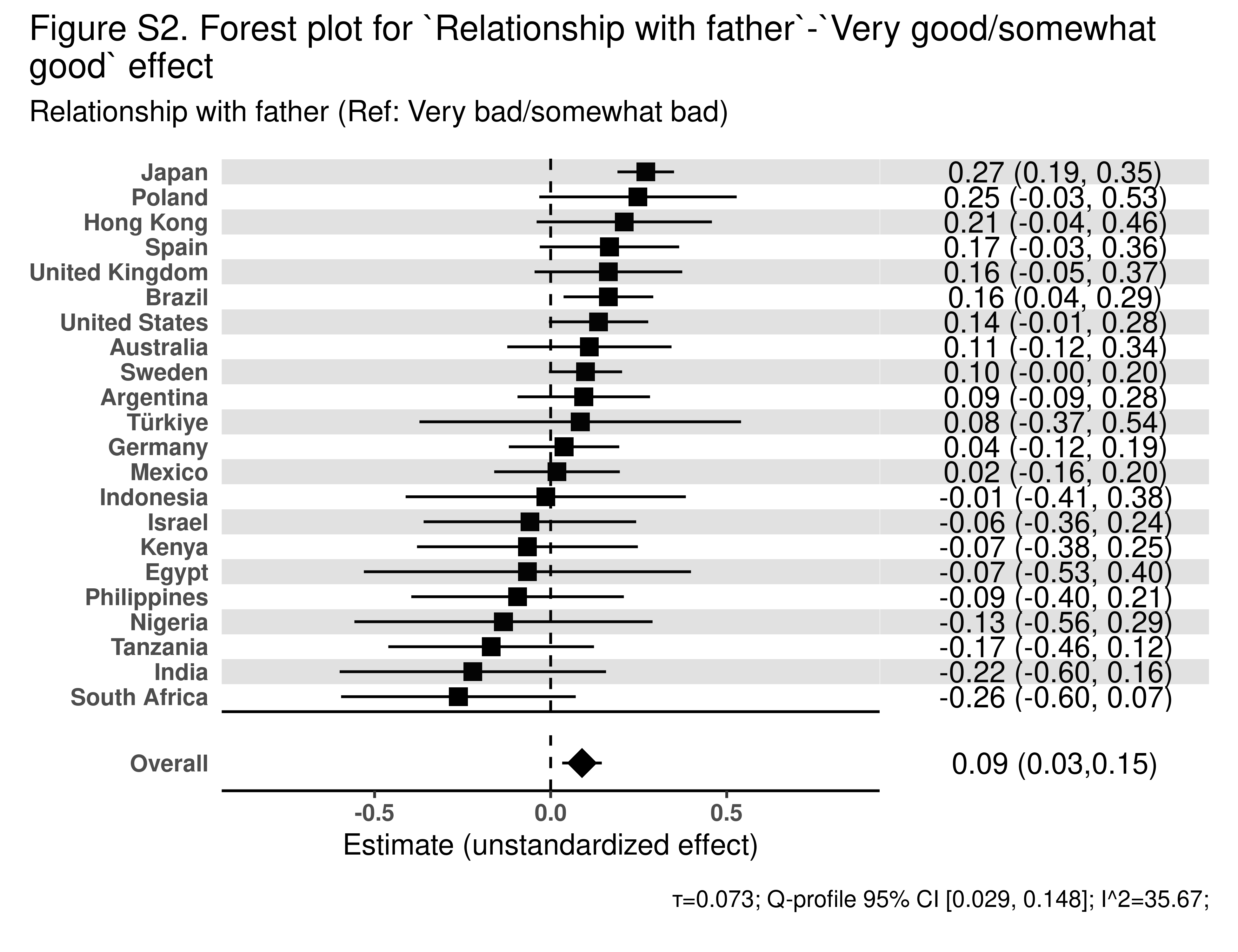

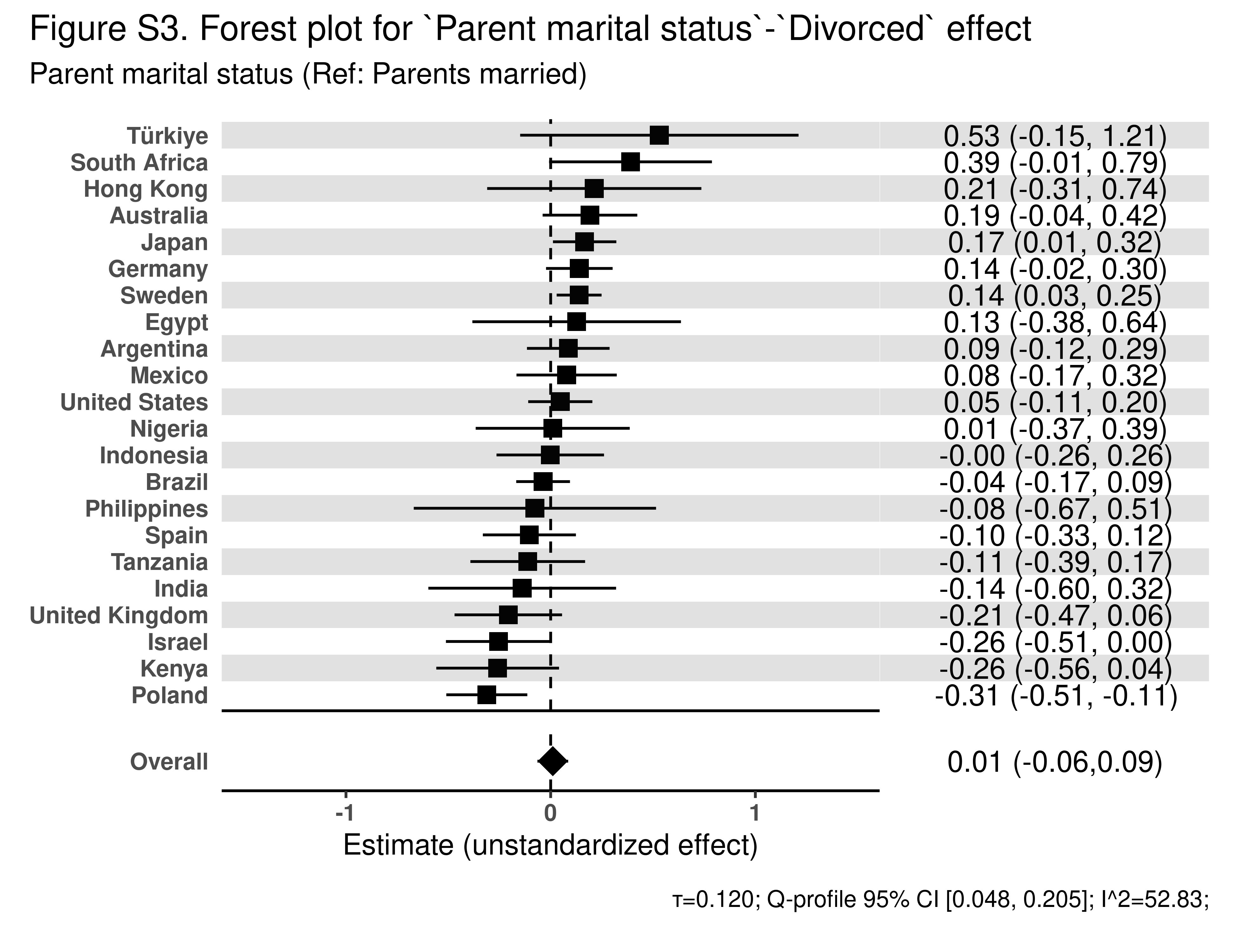

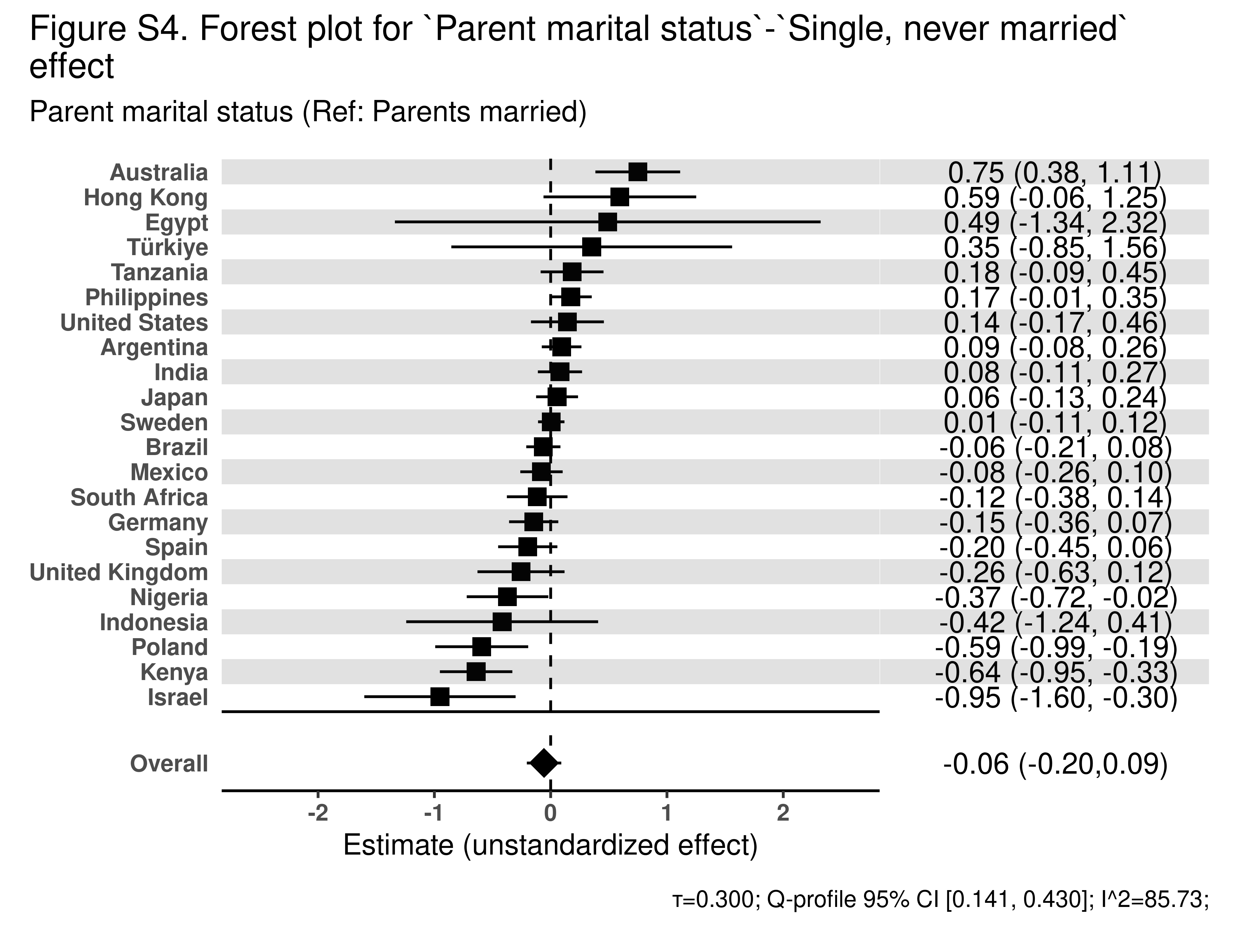

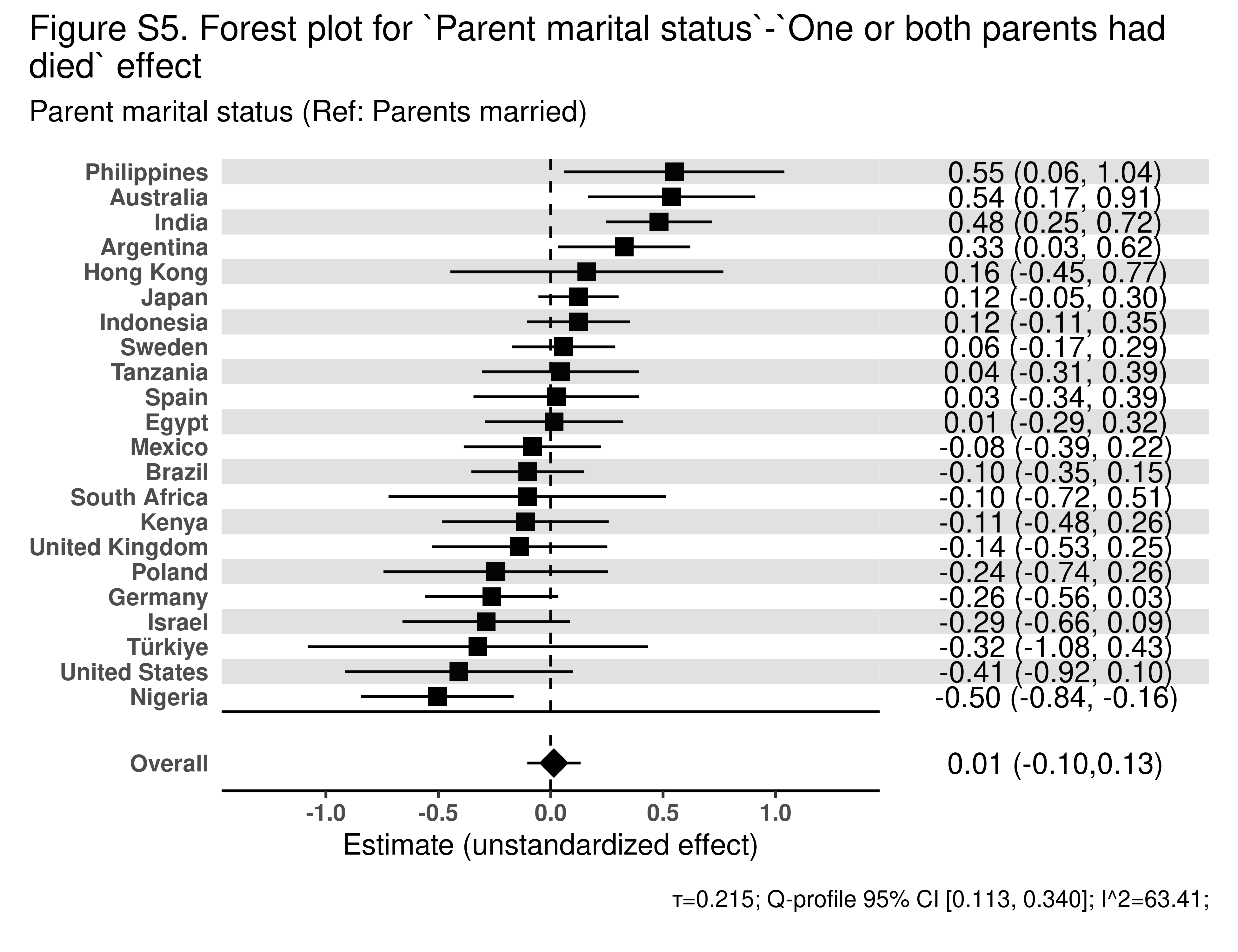

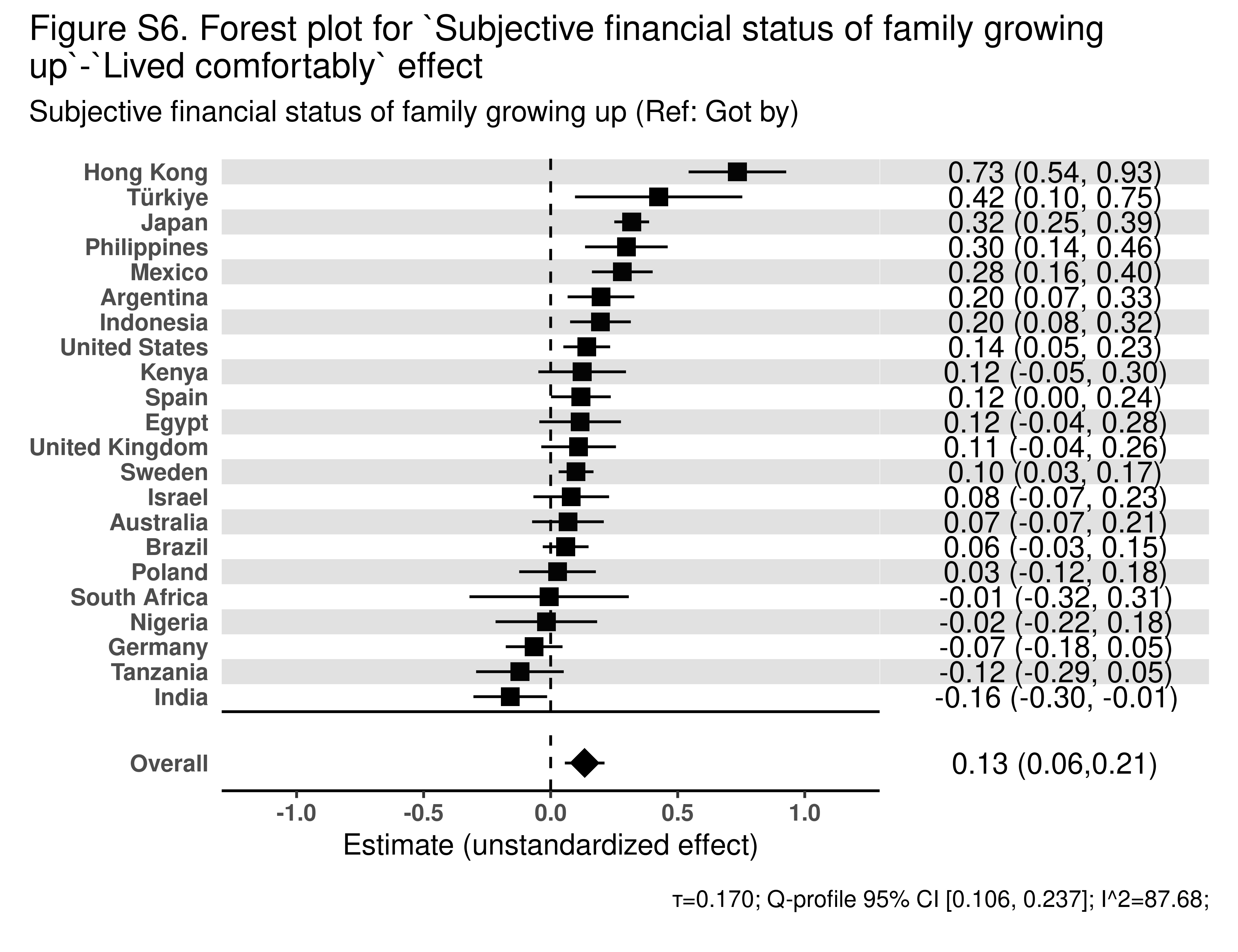

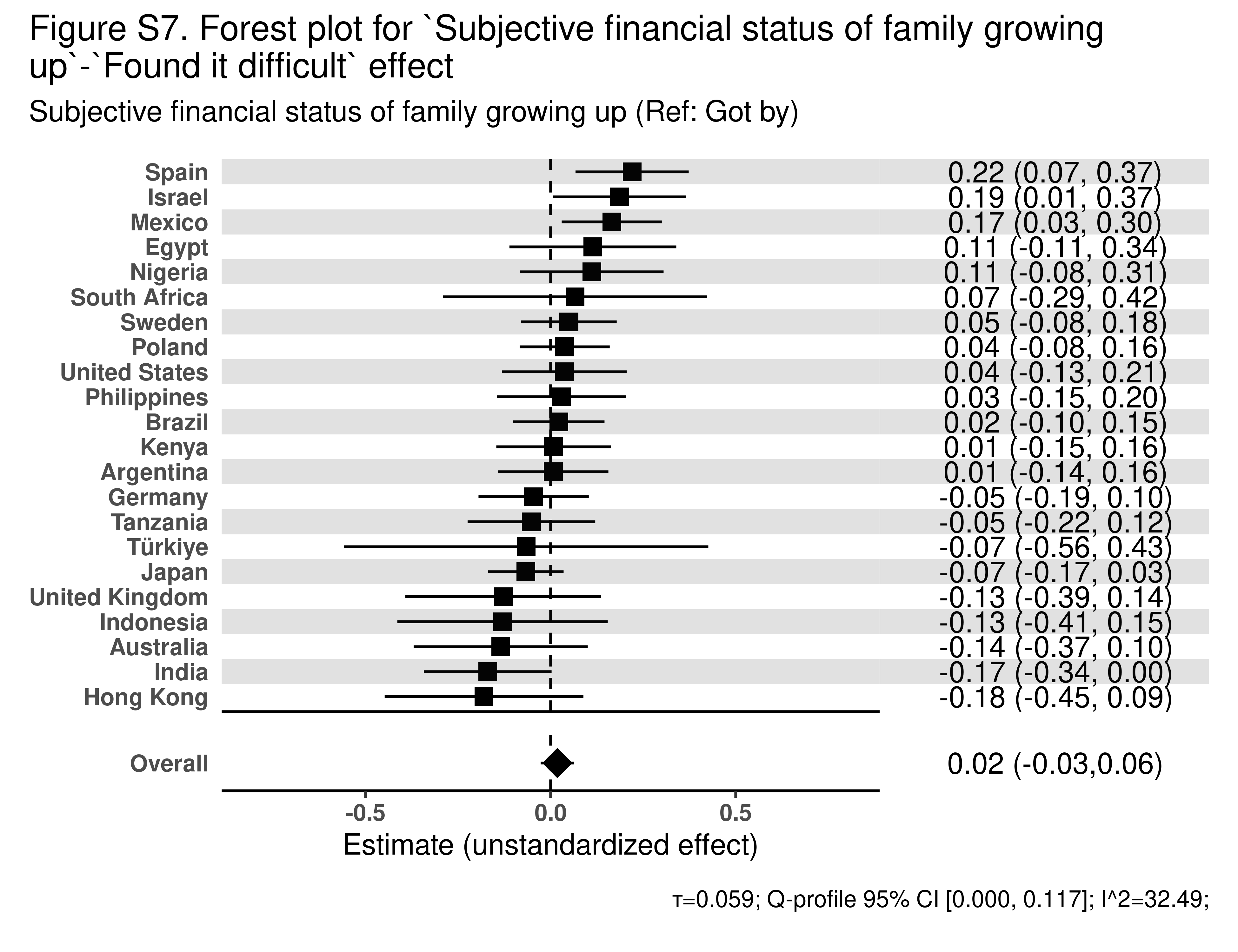

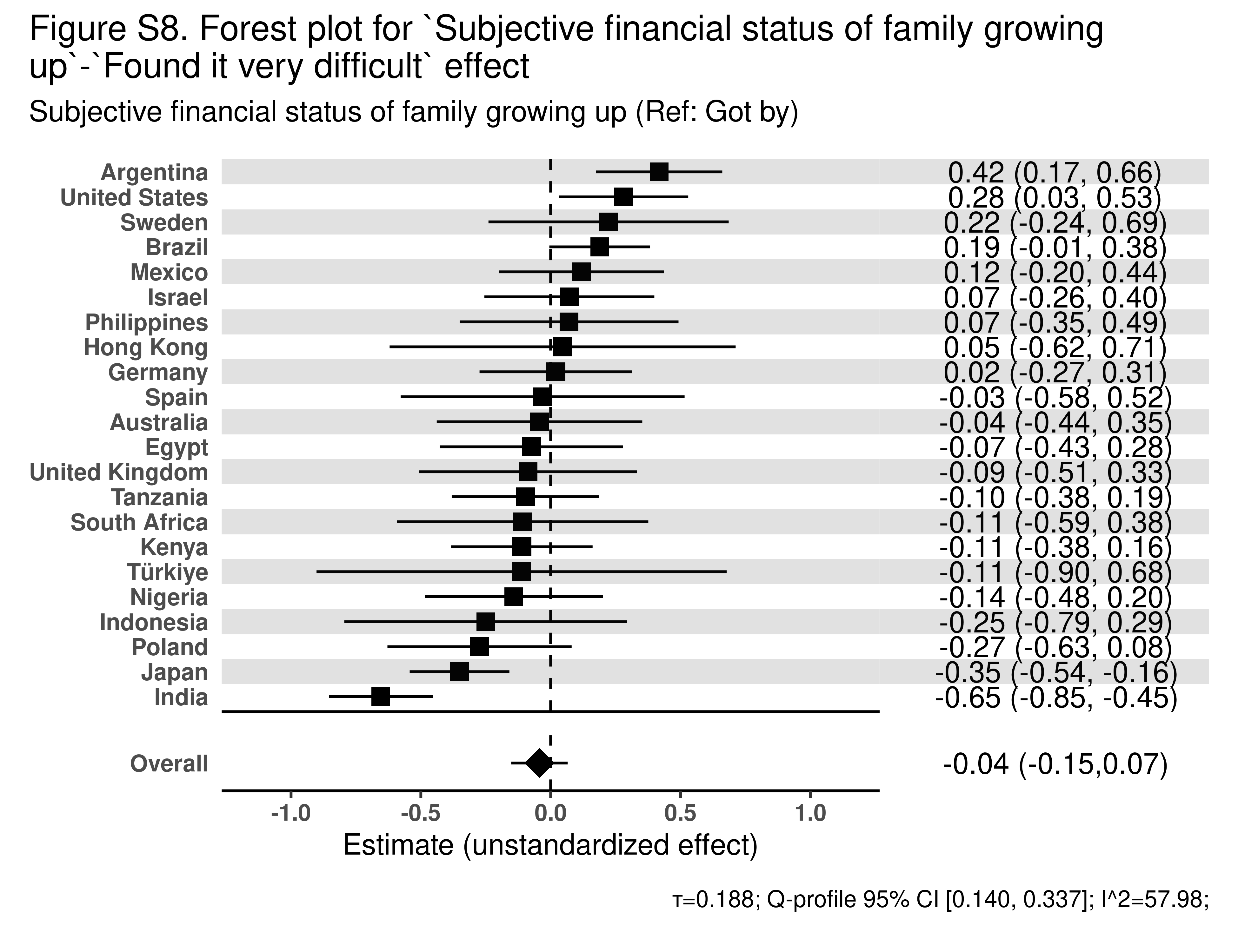

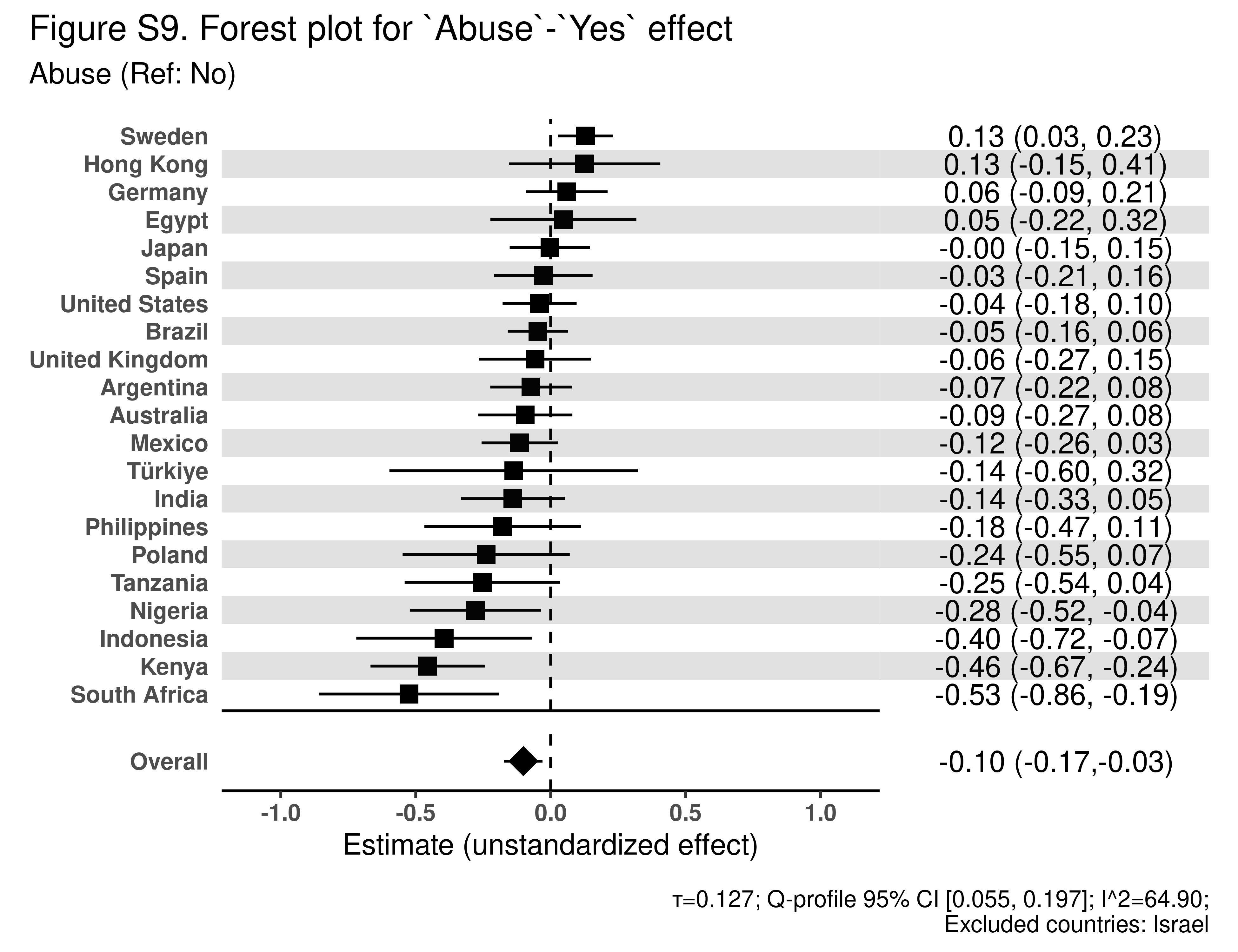

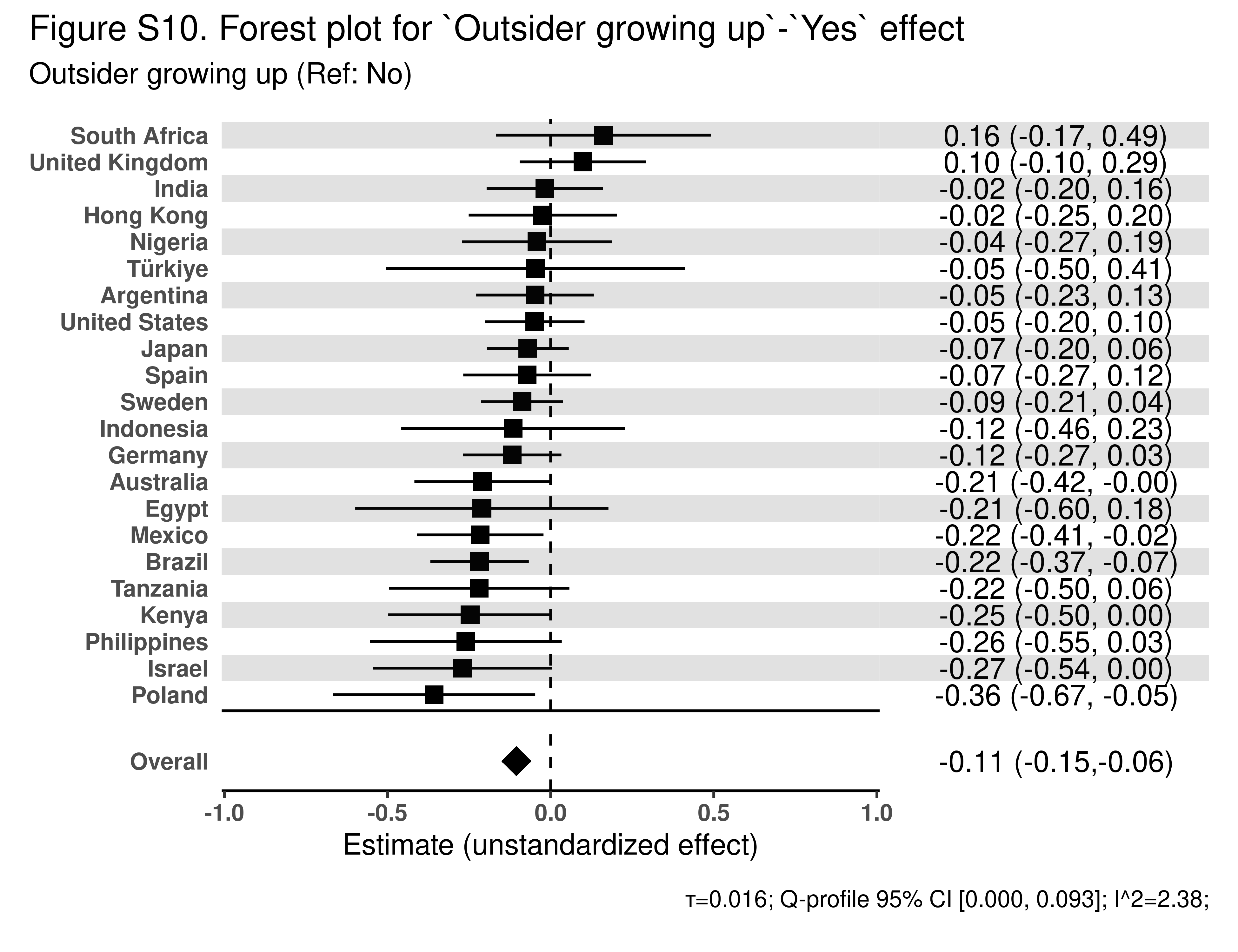

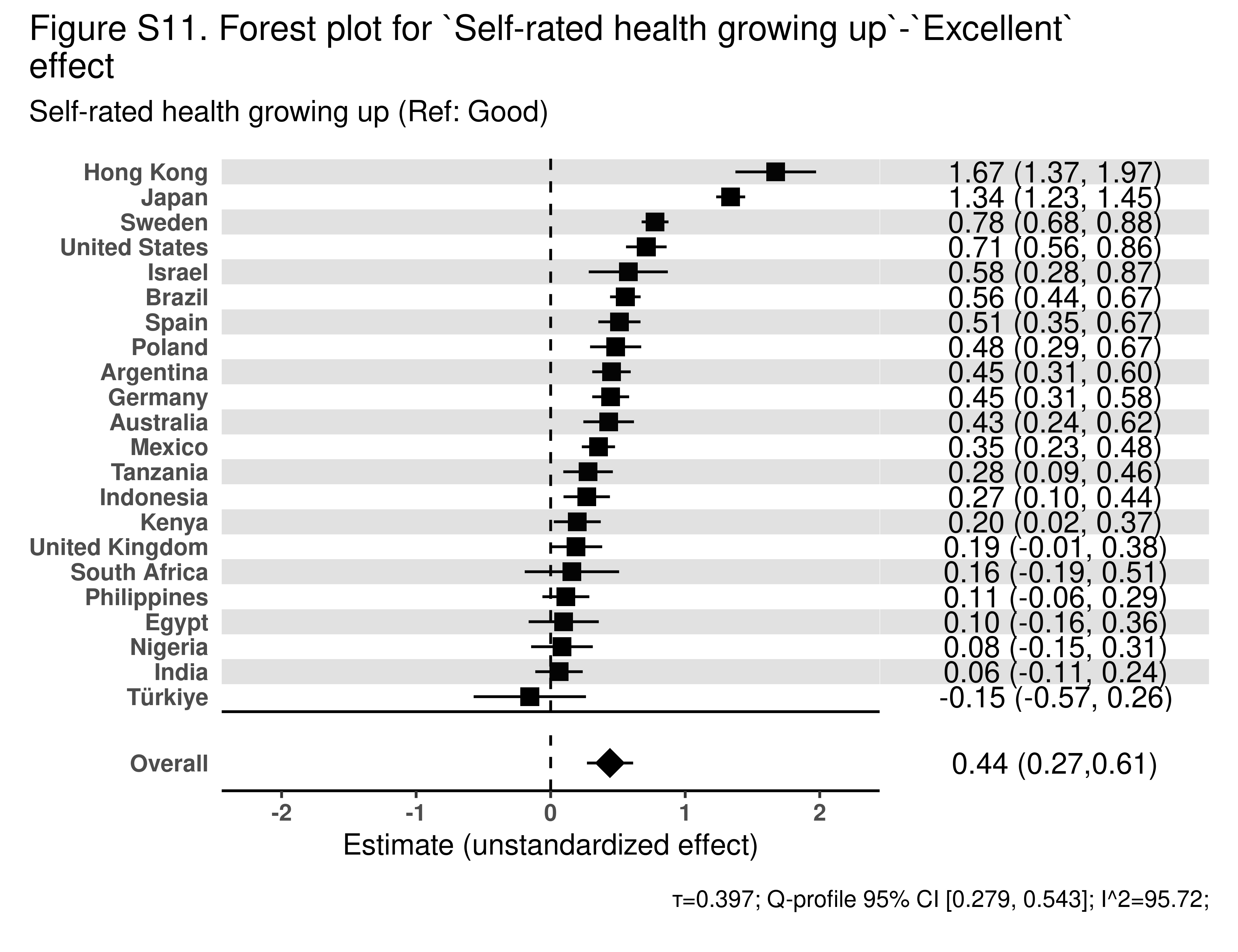

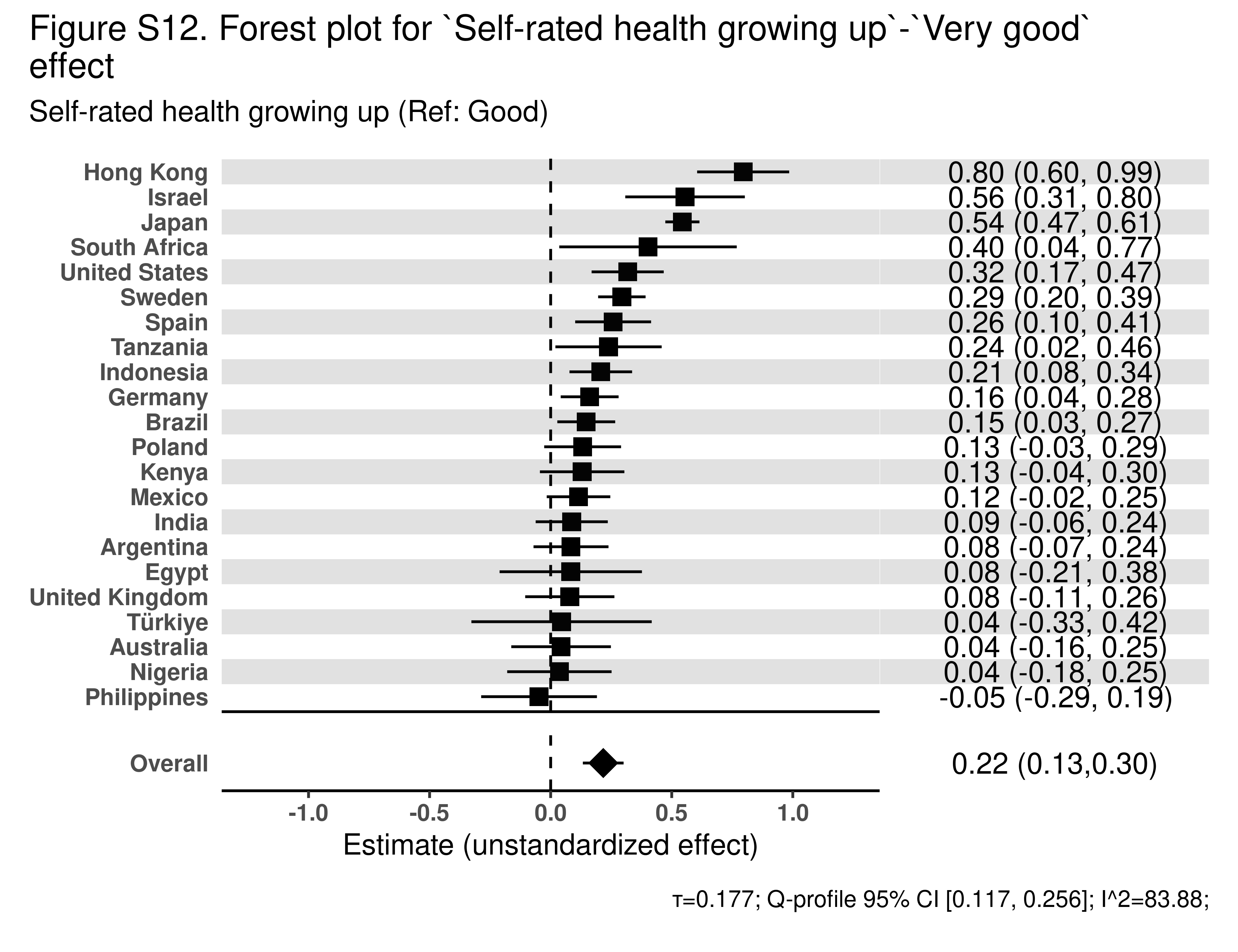

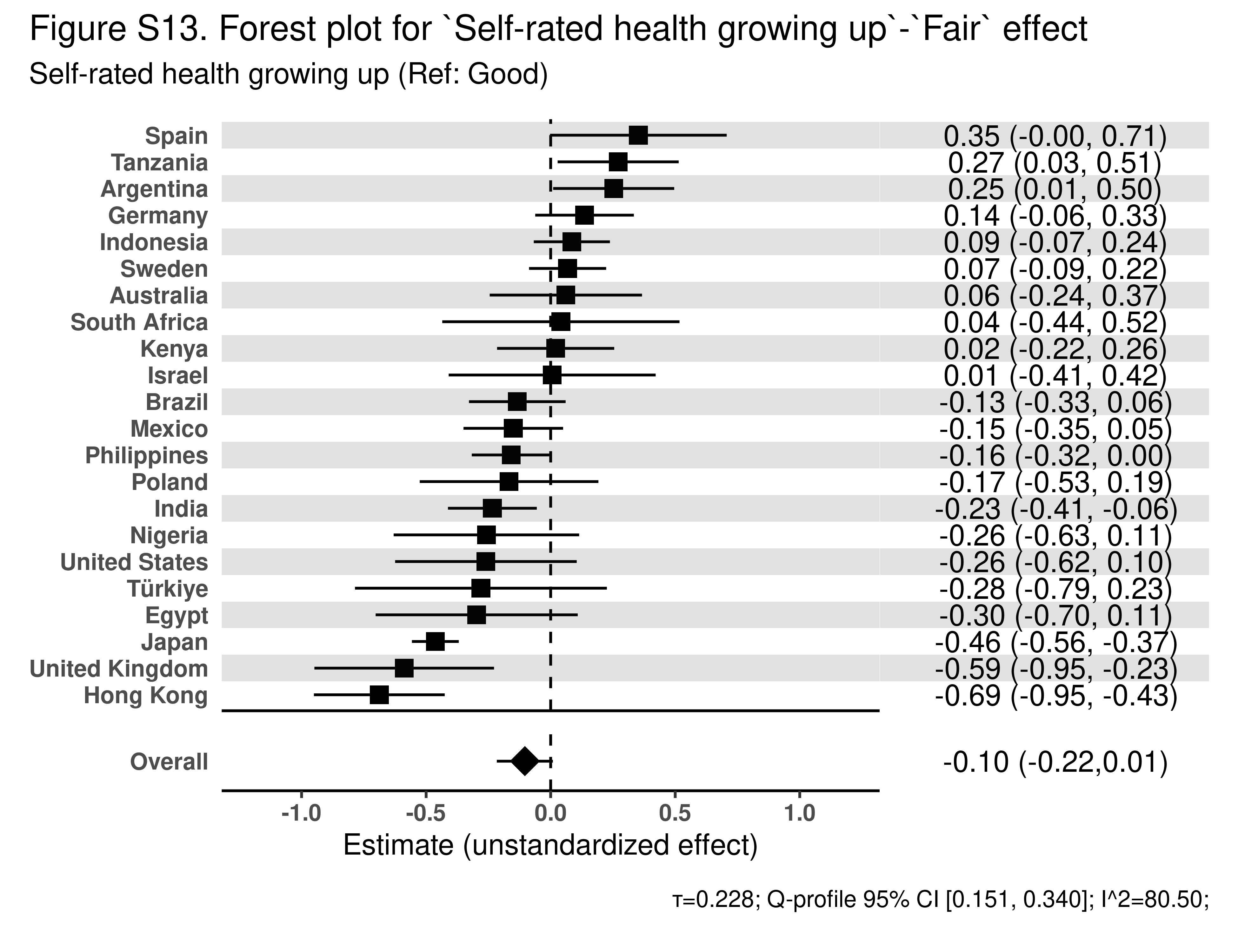

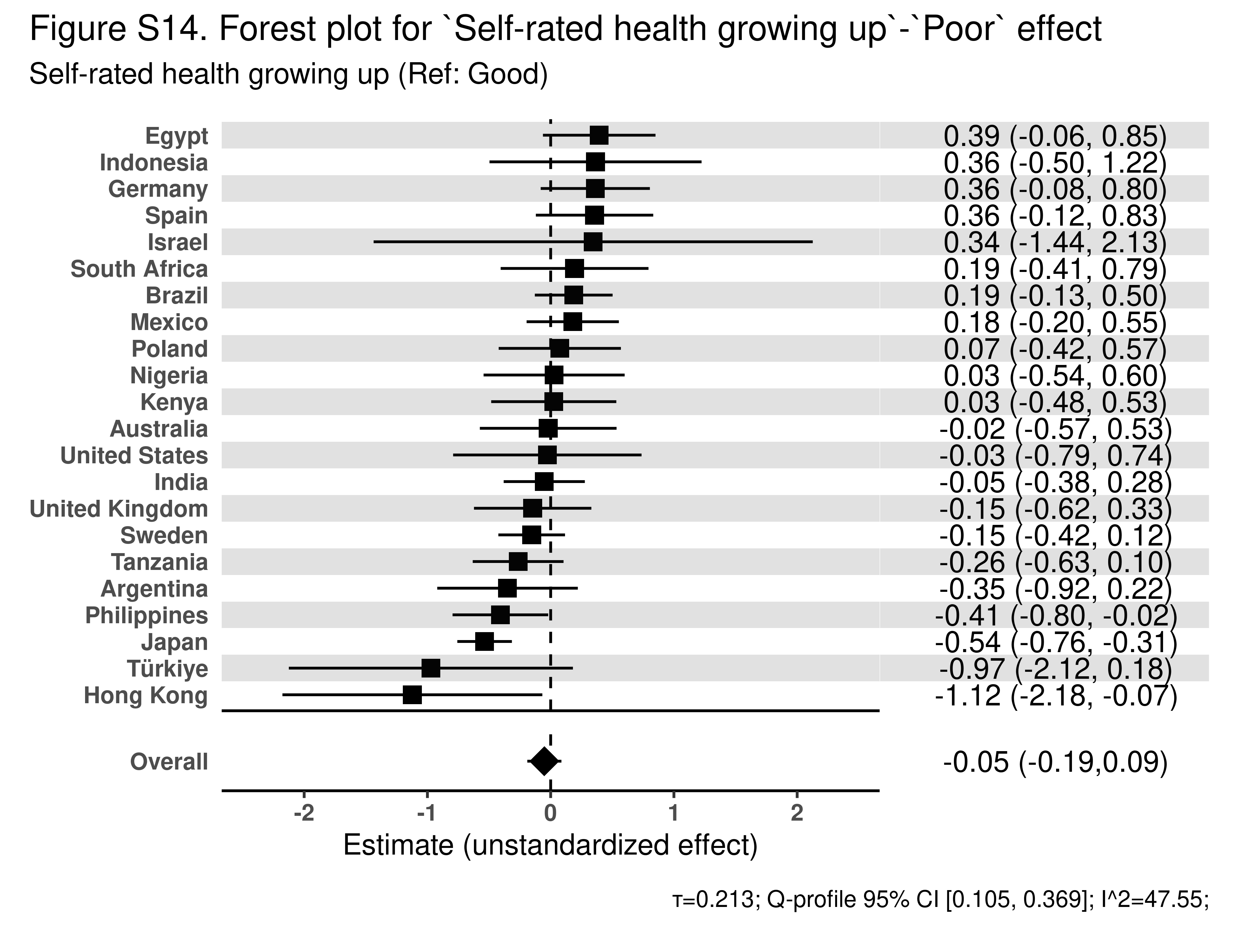

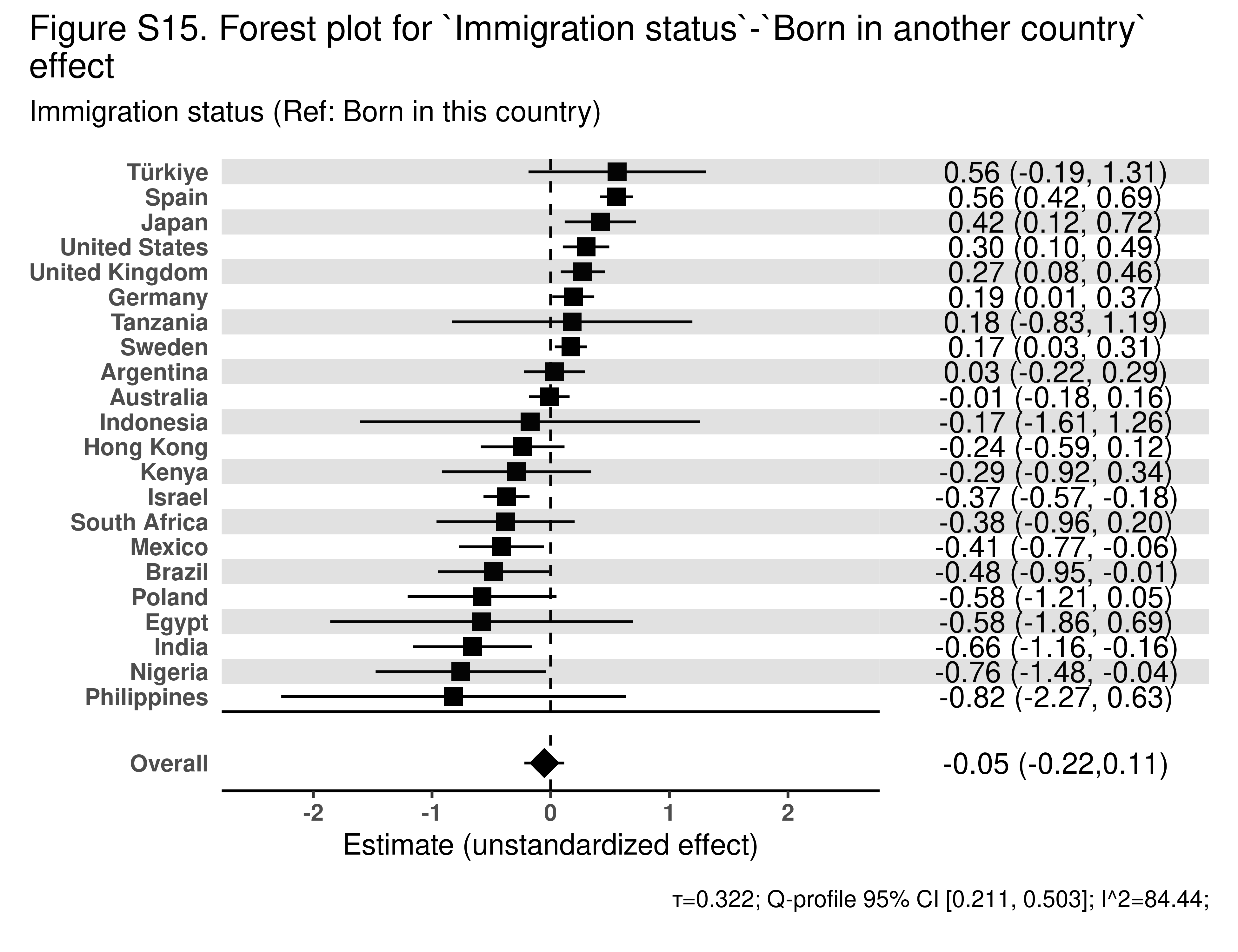

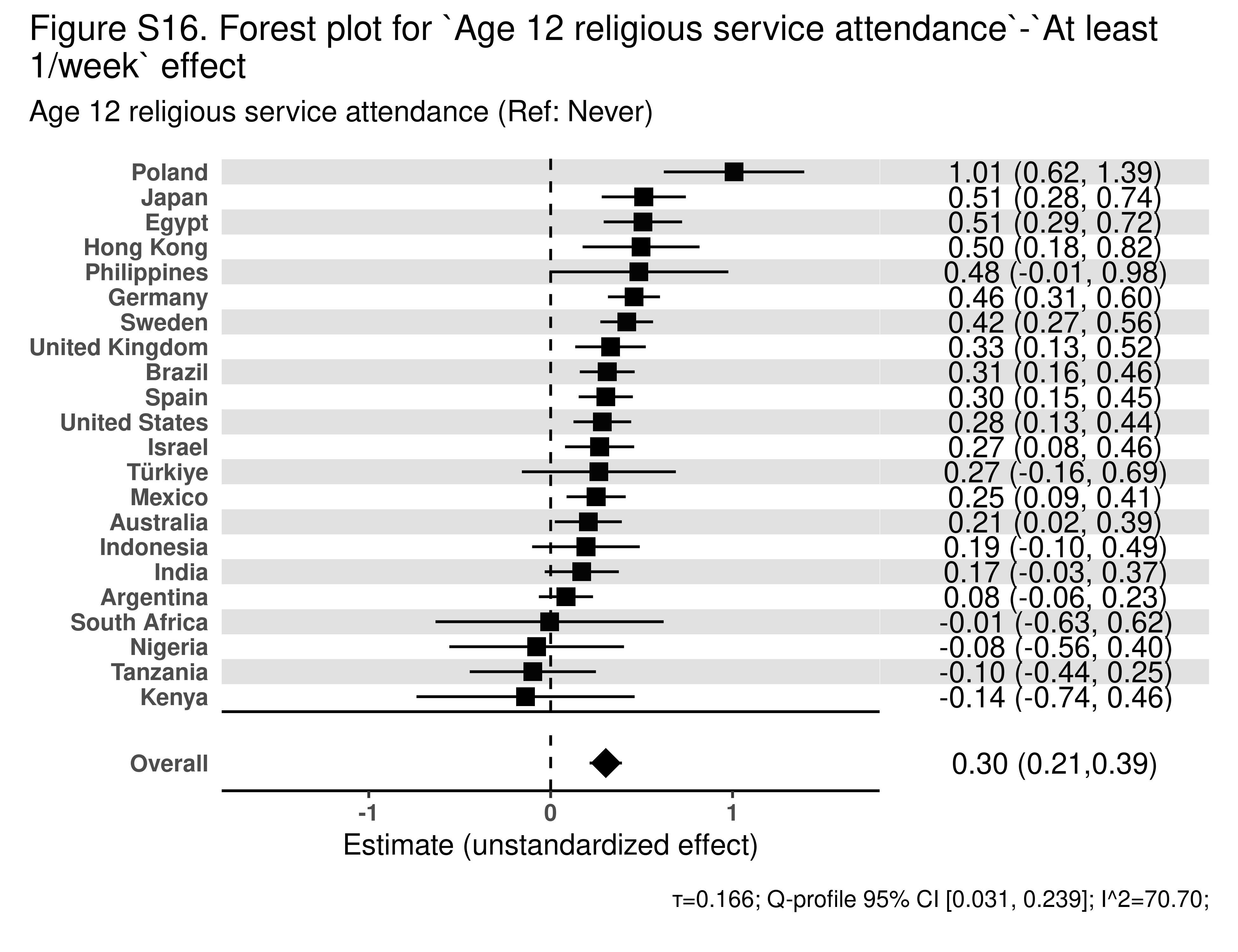

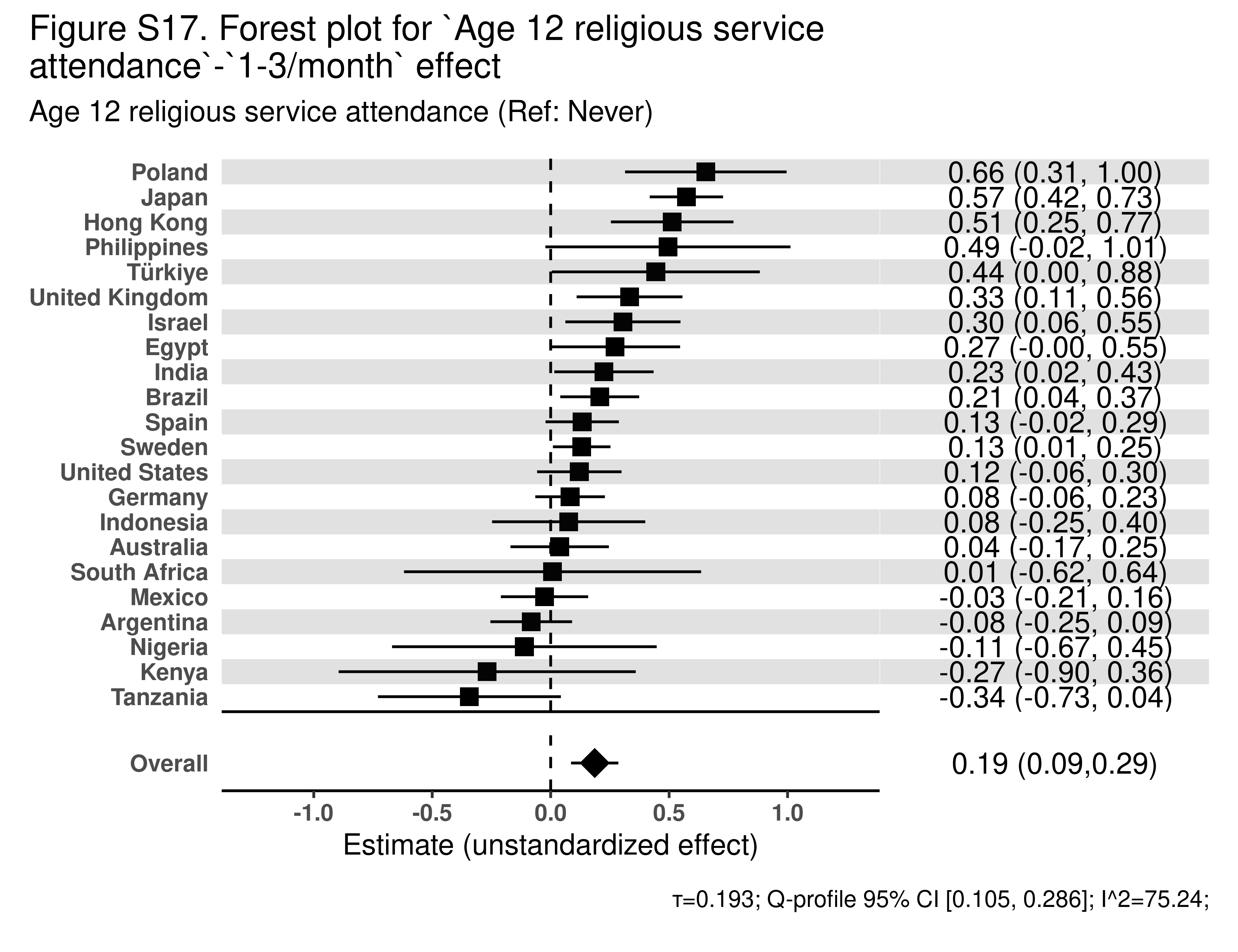

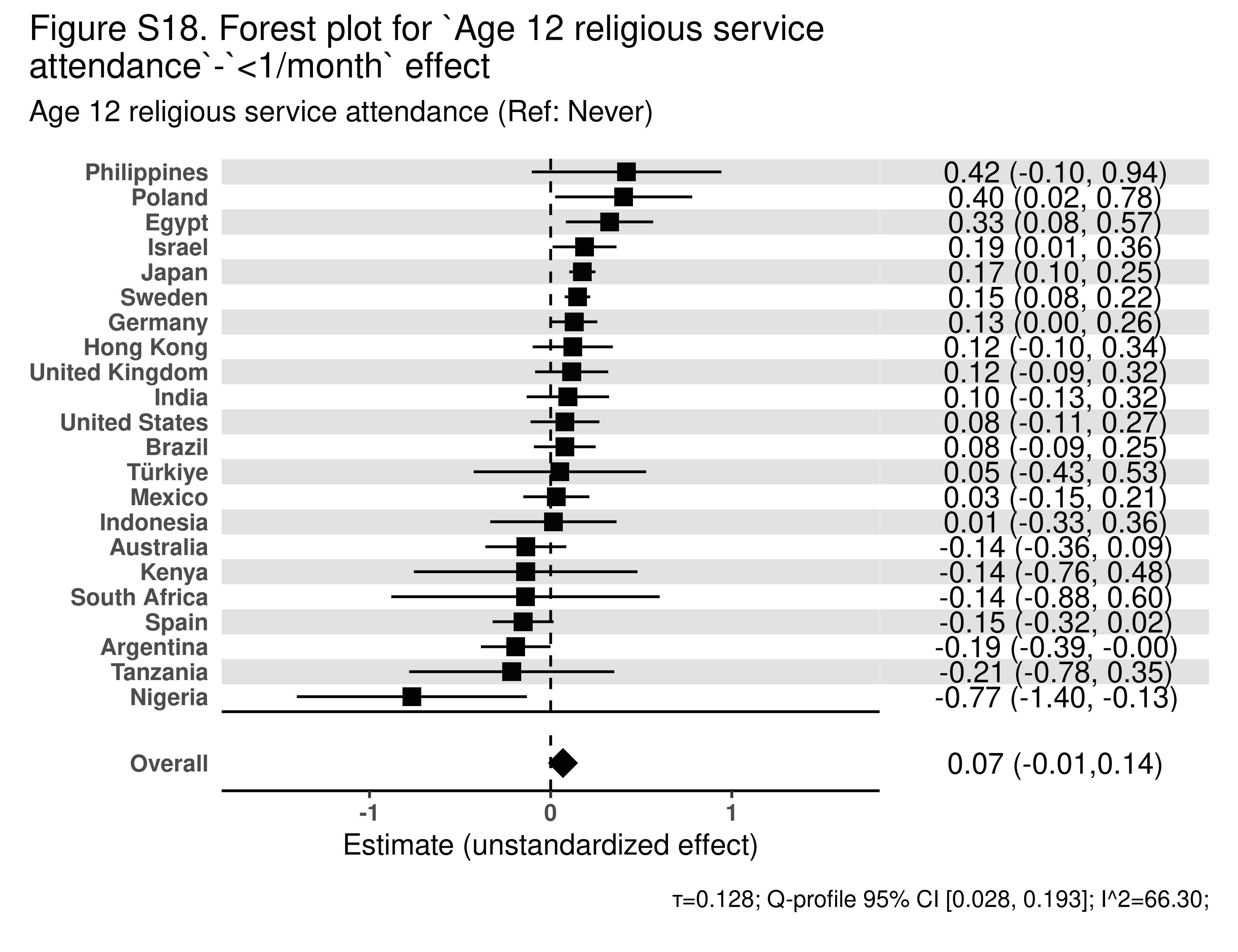

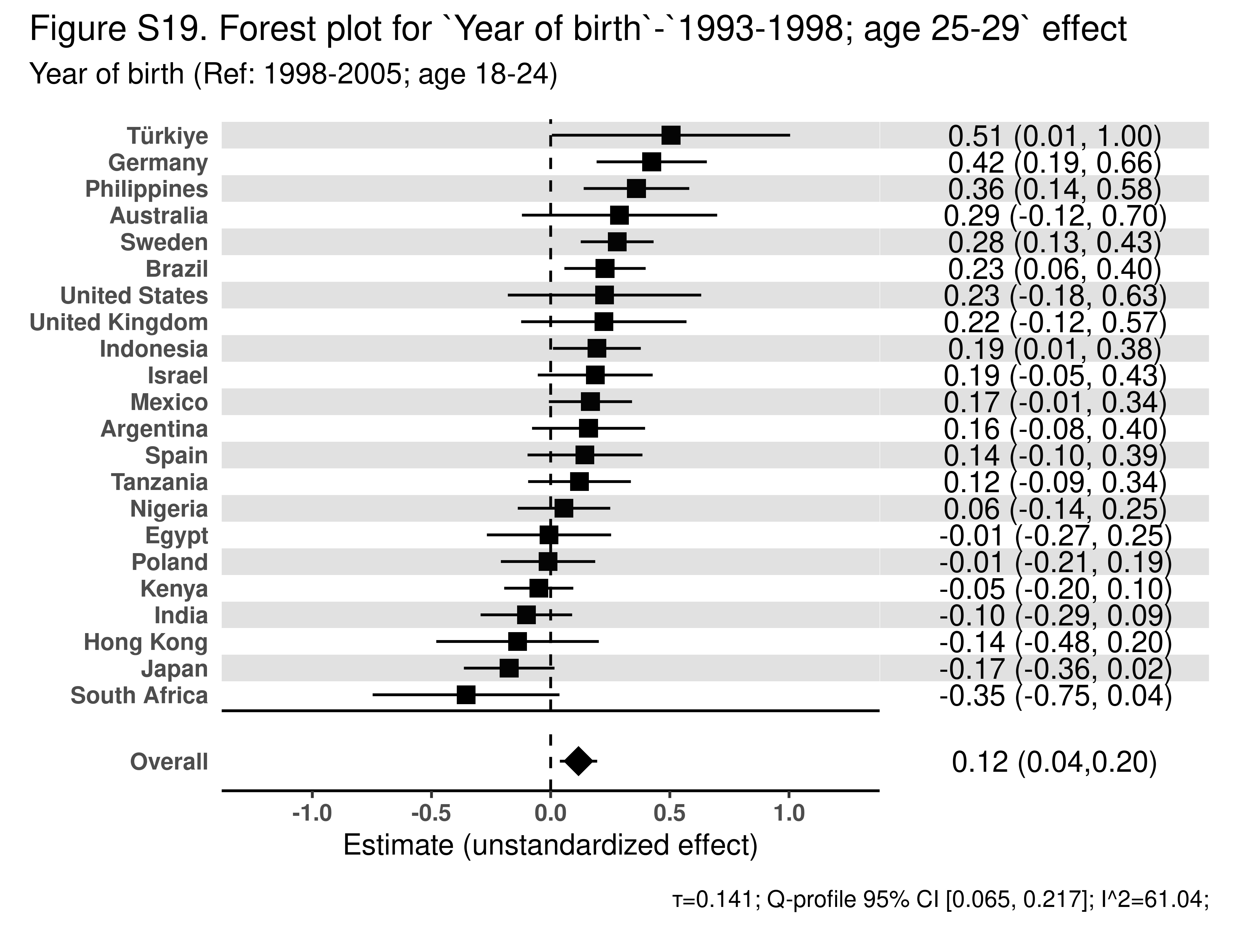

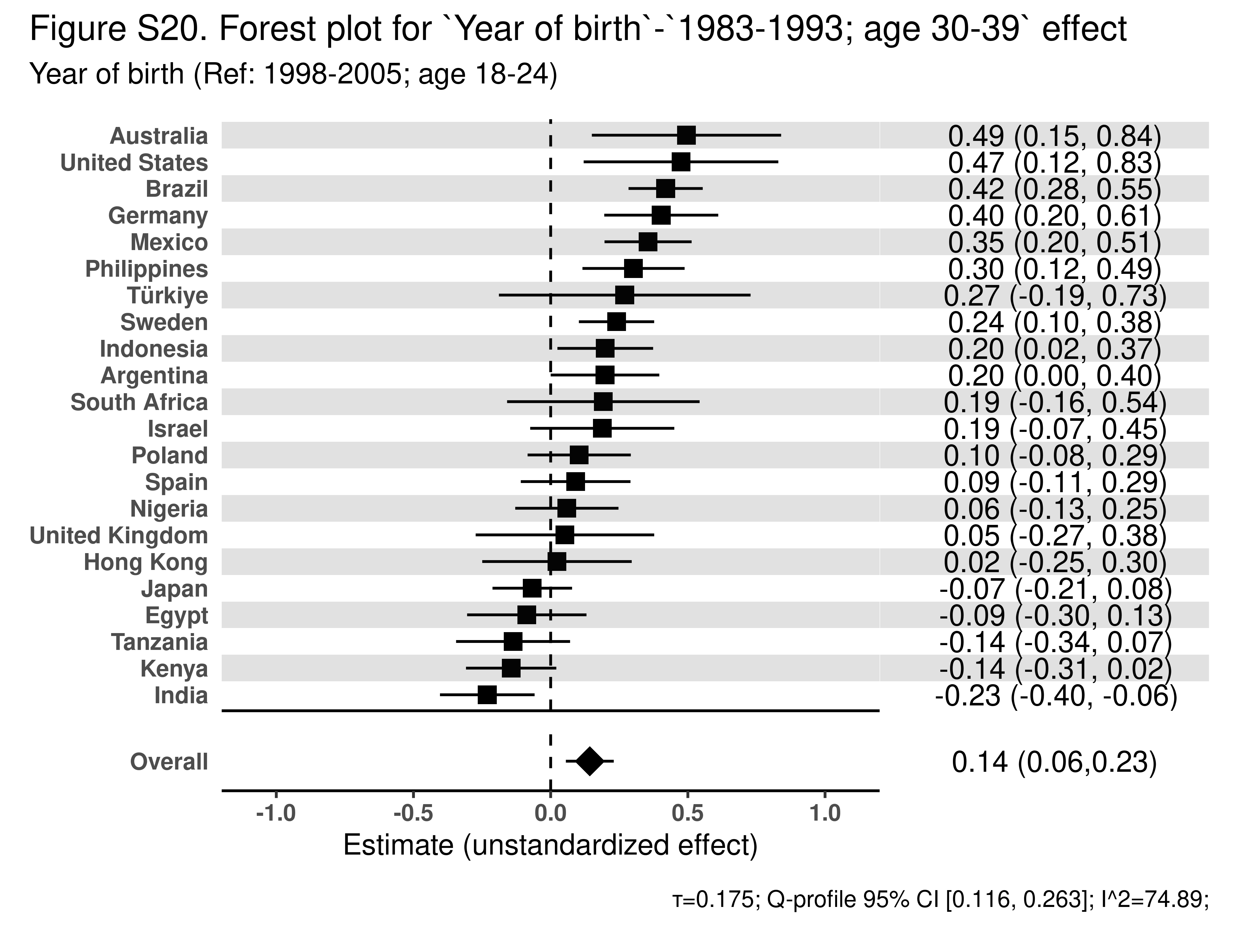

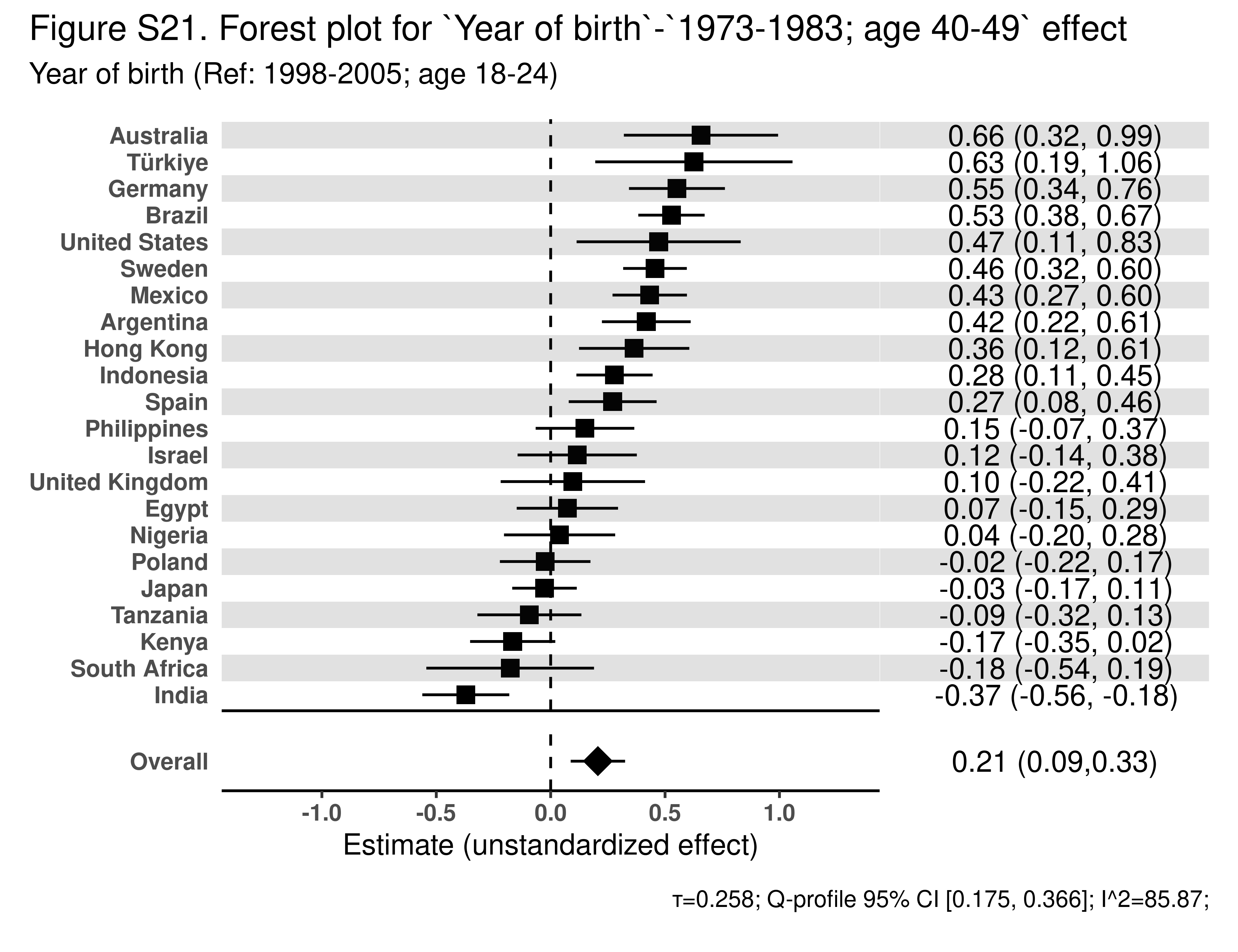

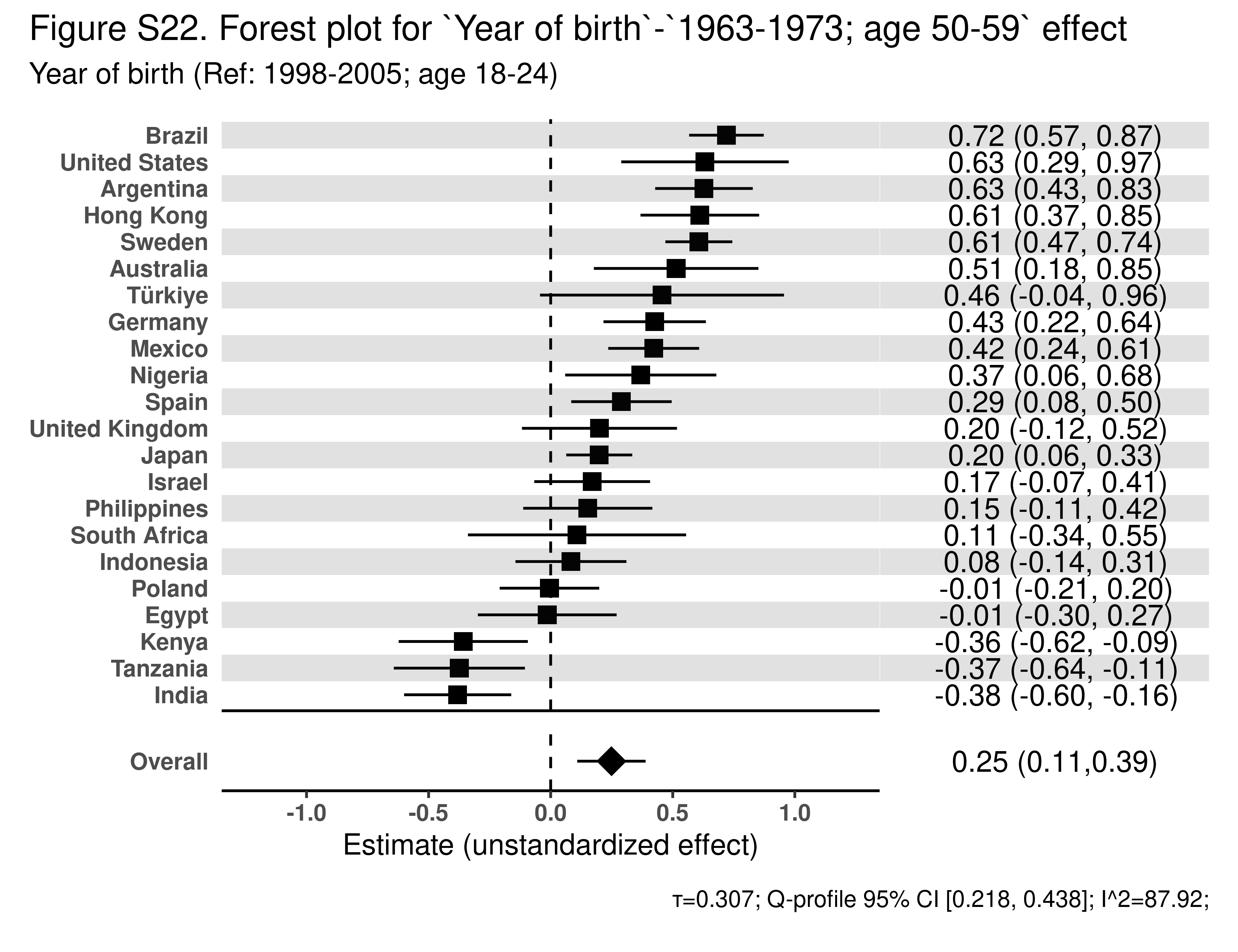

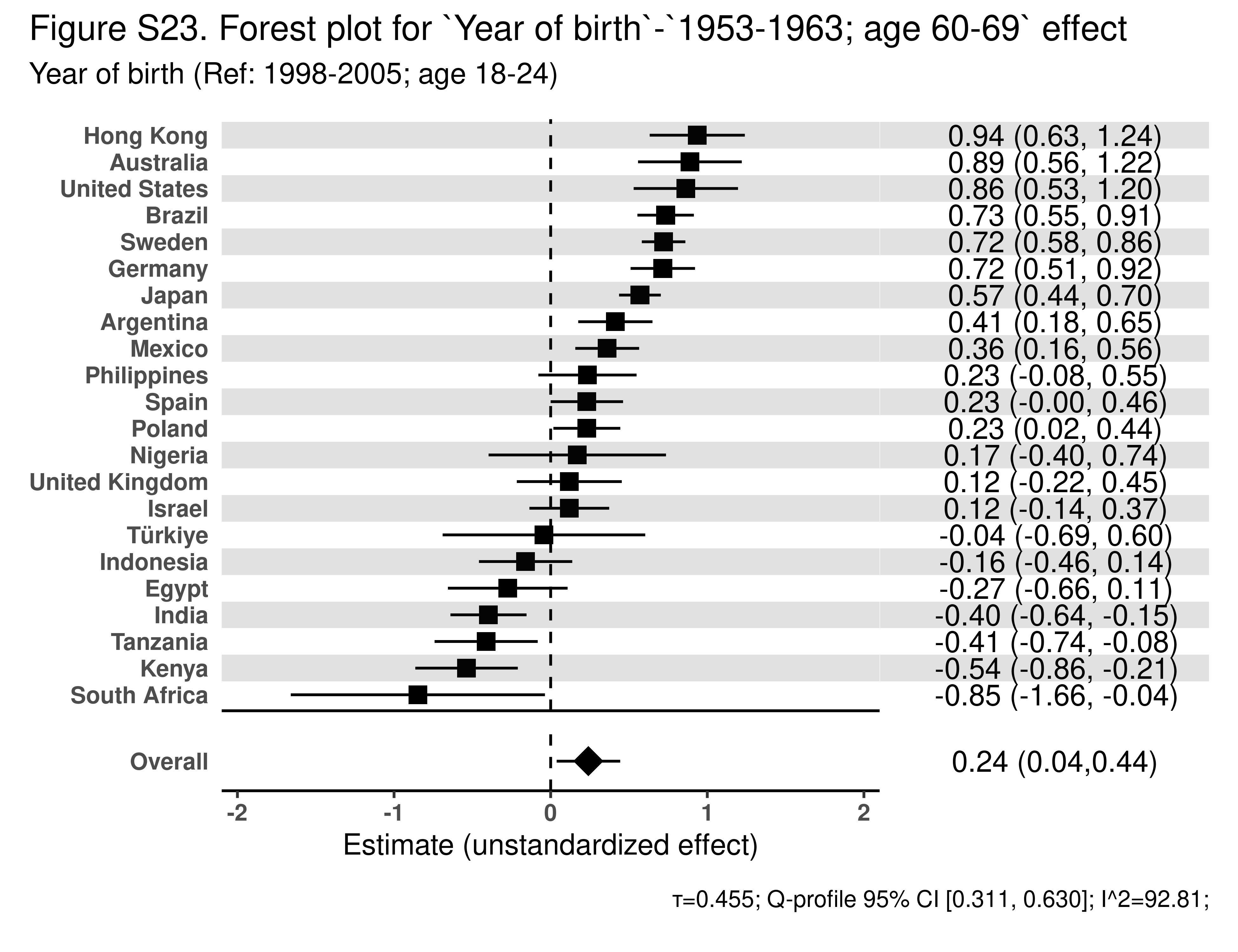

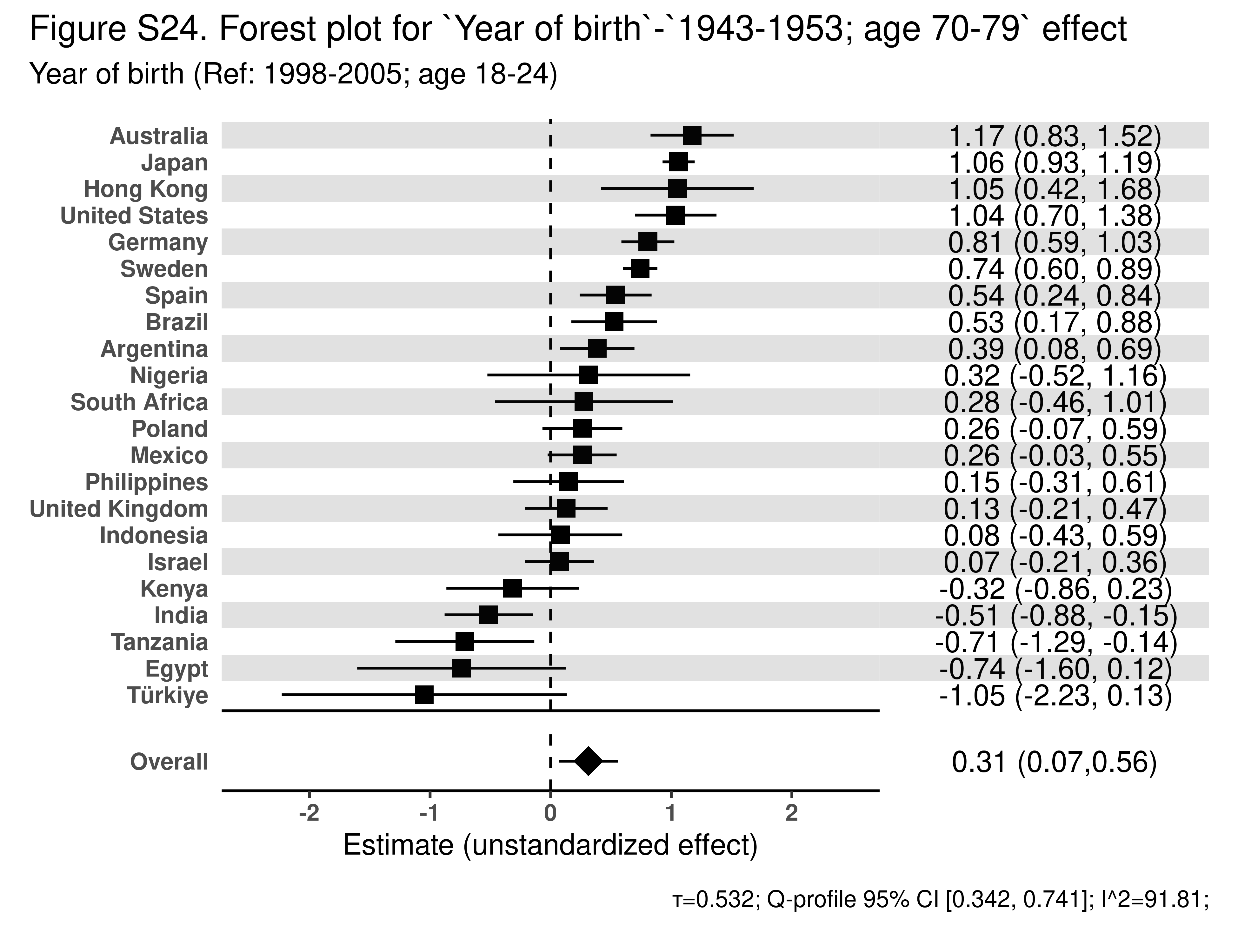

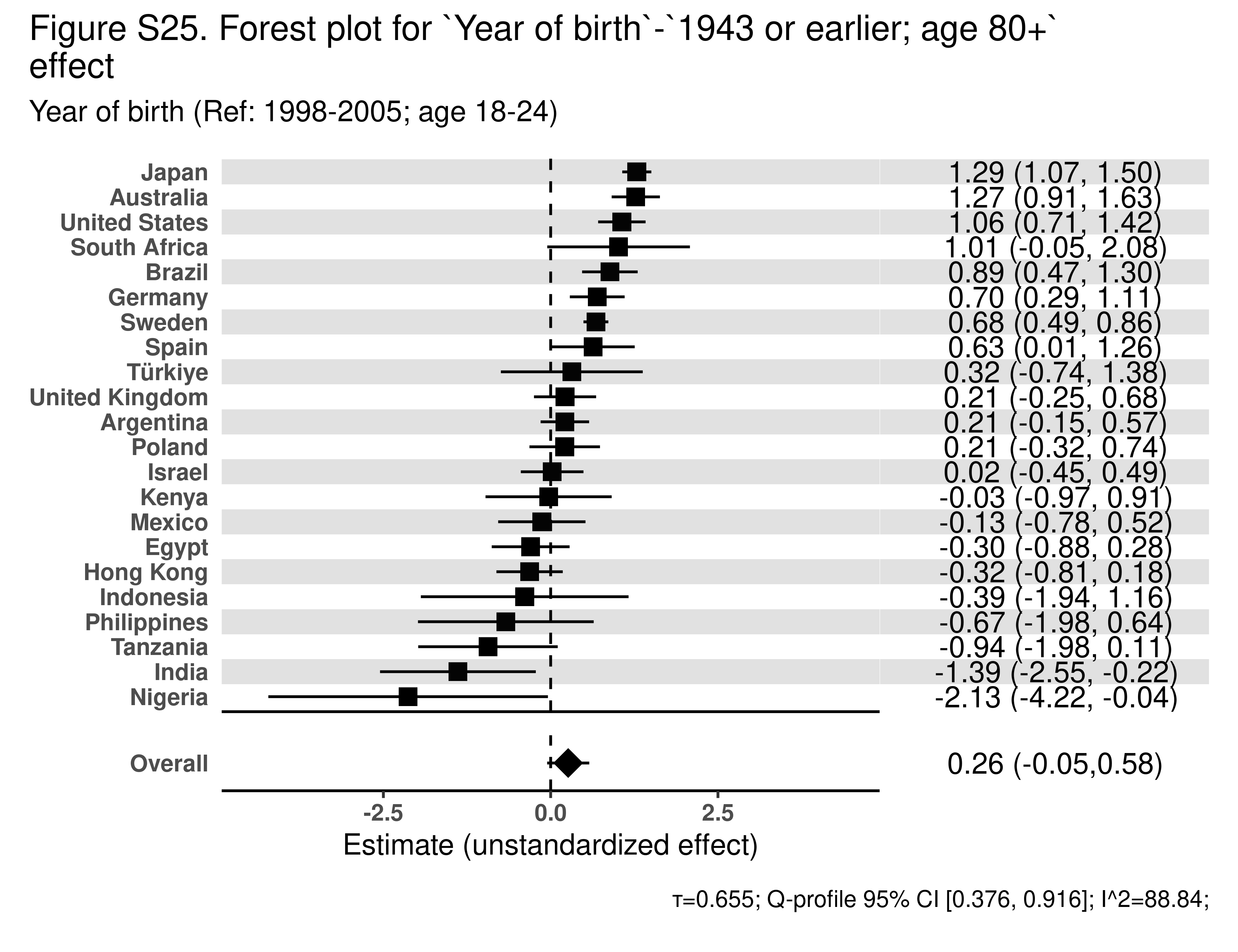

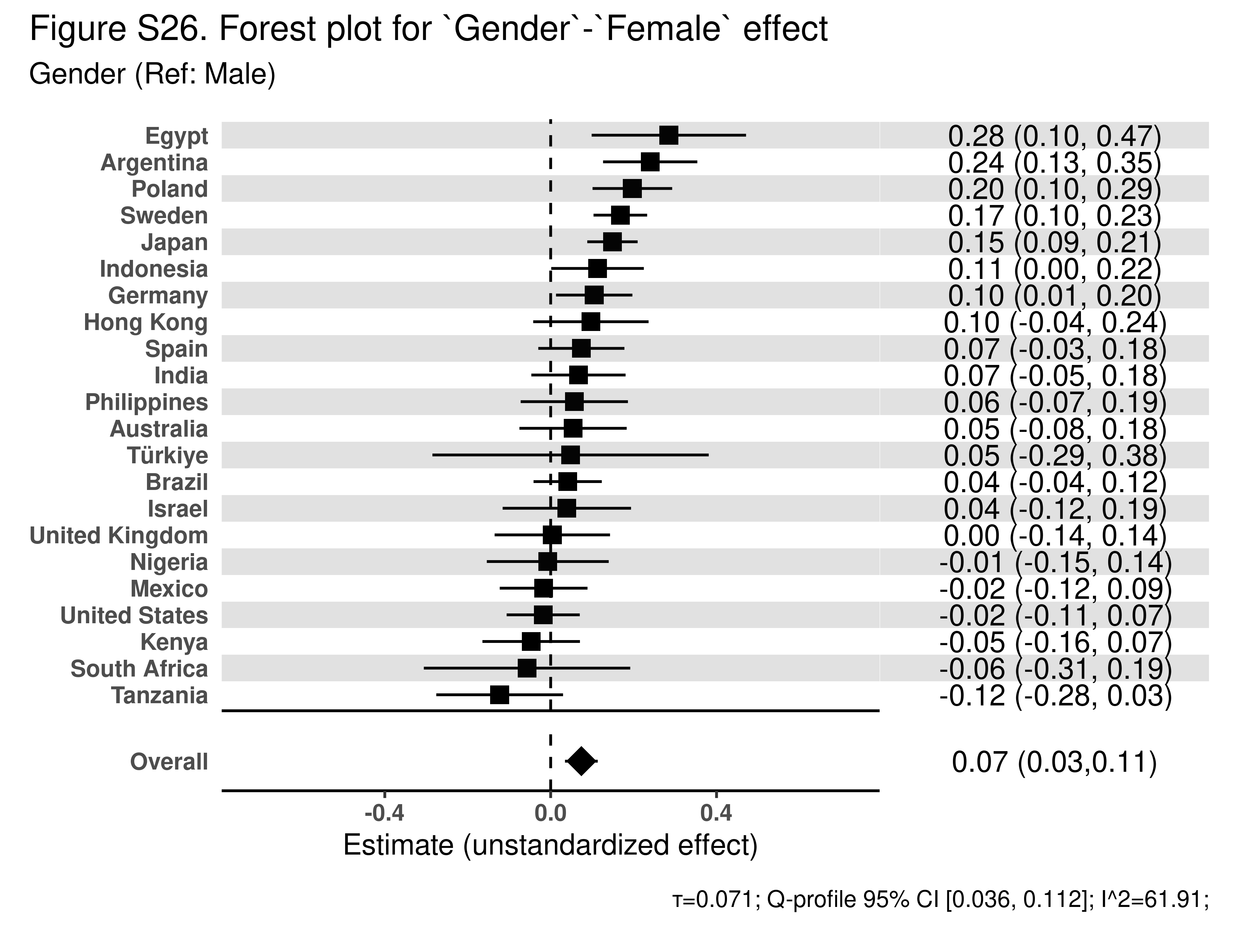

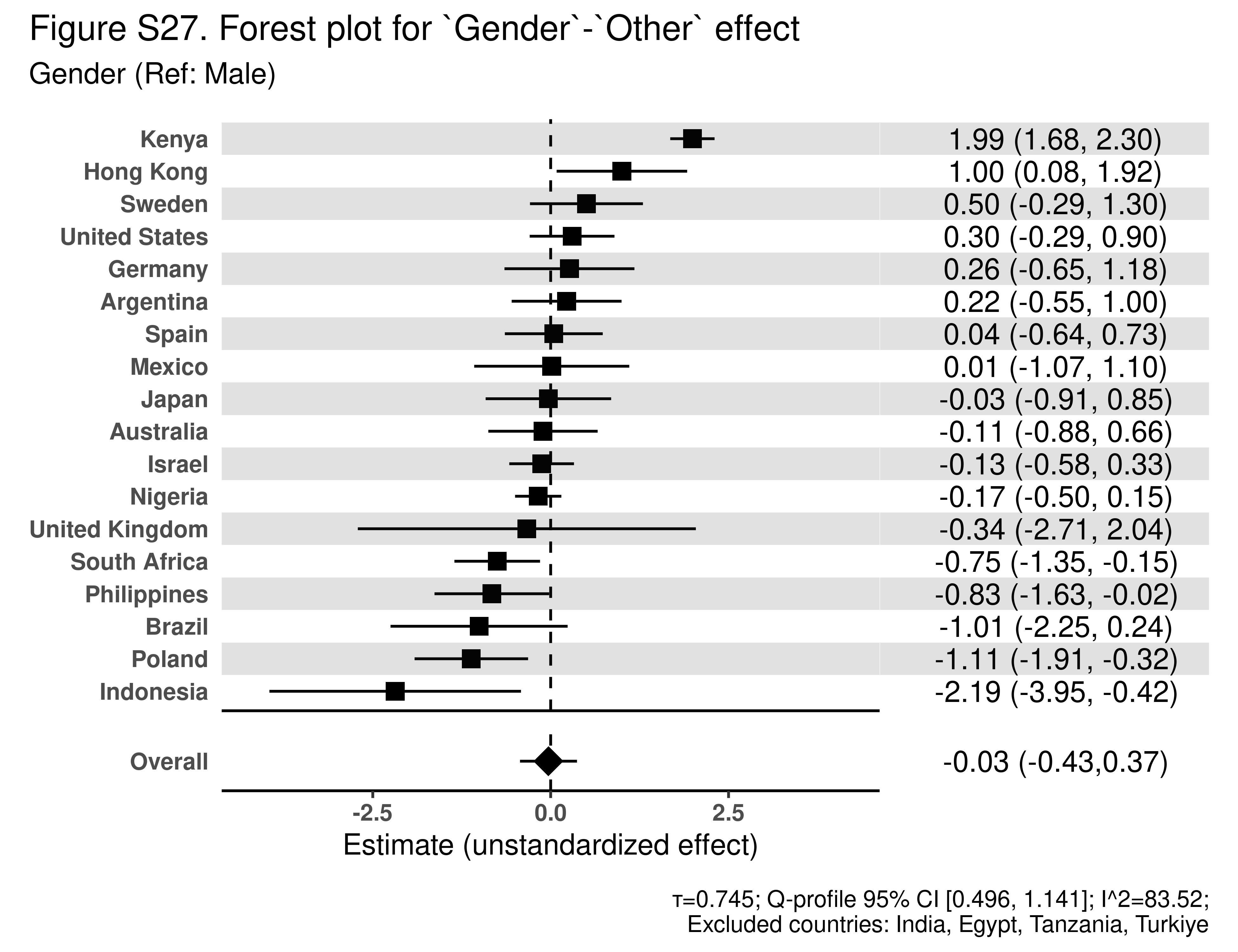
